# Nowcasting Reported COVID-19 Hospitalizations Using De-Identified, Aggregated Medical Insurance Claims Data

**DOI:** 10.1101/2023.12.22.23300471

**Authors:** Xueda Shen, Aaron Rumack, Bryan Wilder, Ryan J. Tibshirani

## Abstract

We propose, implement, and evaluate a method for nowcasting the daily number of new COVID-19 hospitalizations, at the level of individual US states, based on de-identified, aggregated medical insurance claims data. Our analysis proceeds under a hypothetical scenario in which, during the Delta wave, states only report data on the first day of each month, and on this day, report COVID-19 hospitalization counts for each day in the previous month. In this hypothetical scenario (just as in reality), medical insurance claims data continues to be available daily. At the beginning of each month, we train a regression model, using all data available thus far, to predict hospitalization counts from medical insurance claims. We then use this model to nowcast the (unseen) values of COVID-19 hospitalization counts from medical insurance claims, at each day in the following month. Our analysis uses properly-versioned data, which would have been available in real-time at the time predictions are produced (instead of using data that would have only been available in hindsight). In spite of the difficulties inherent to real-time estimation (e.g., latency and backfill) and the complex dynamics behind COVID-19 hospitalizations themselves, we find altogether that medical insurance claims can be an accurate predictor of hospitalization reports, with mean absolute errors typically around 0.4 hospitalizations per 100,000 people, i.e., proportion of variance explained around 75%. Perhaps more importantly, we find that nowcasts made using medical insurance claims are able to qualitatively capture the dynamics (upswings and downswings) of hospitalization waves, which are key features that inform public health decision-making.

## 1 Introduction

Timely access to public health data is critical to enable informed decision making during infectious disease outbreaks. However, setting up and maintaining public health reporting pipelines can be a burden on the health system itself. For example, beginning in May 2020, hospitals in the US have been required to report data on COVID-19 hospitalizations to the Department of Health and Human Services (HHS) (Department of Health and Human Services, 2023). This data has been critical for understanding the state of the pandemic and the current load on the health system. But the frequent reporting required for up-to-date situational awareness (daily, throughout most of the pandemic) has been quite difficult to implement and maintain. In an effort to achieve compliance, the Centers for Medicare and Medicaid Services (CMS) issued regulations in August 2020 that threatened to expel hospitals from the Medicare program, and apply monetary penalties, if they failed to comply with daily COVID-19 reporting requirements. This was strongly and openly opposed by the American Hospital Association (AHA) (Nickels, 2020), but the regulations remained in place for nearly three years, lasting until the conclusion of the COVID-19 Public Health Emergency in May 2023.

An intriguing alternative lies in data streams which already exist and are maintained for other purposes, yet are relevant for inferring disease activity. One example is medical insurance claims, which are filed by a healthcare provider to seek reimbursement from an insurance company for medical services performed. In this paper, we examine the use of *de-identified, aggregated* medical insurance claims data as a complement to public health reporting over the course of the pandemic. Specifically, we investigate the following: if hospital reporting on COVID-19 would have been reduced in frequency from daily to monthly during the Delta wave, could we use signals derived from medical insurance claims to accurately *nowcast* COVID-19 hospitalization counts during the interim periods?

Medical insurance claims have long been utilized in public health policy analysis, ranging from economic implications of healthcare (e.g., recent examples include Panczak et al. (2018); Li and Yang (2021); Sakai et al. (2019); Zheng and Peng (2021); Durizzo et al. (2022); Mori et al. (2022)), to examinations of treatment effectiveness and satisfaction (e.g., Nakayama et al. (2017); Jung et al. (2020); Geng et al. (2021); Yao et al. (2022); Song et al. (2023)). All of these works, however, are *retrospective* in nature: they seek to develop an understanding of a particular phenomenon using data that would not have been available in real-time, but only in hindsight. In contrast, our goal to carry out an analysis that reflects *real-time* estimation, so that we can understand (to the best extent we can) how our models would perform if they were to be operationalized in the future, for true prospective nowcasting. This requires us to use properly-versioned data at all times in our analysis, which would have been available in real-time, at the time nowcasts are produced. This is true of all data sources in question, but it is especially crucial for medical insurance claims signals: these signals are subject to heavy revisions, as claims can be filed long after a service was performed (we describe this more concretely later in the paper), altering previously-computed signal values. Therefore, using finalized (rather than properly-versioned) values of medical insurance claims signals for modeling and prediction can present a misleading picture of nowcasting performance.

Figure 1 displays an example of this, which plots reported COVID-19 hospitalizations in California, over a 1 year period spanning the winter wave of 2020 through the Delta wave of 2021. It also compares a signal derived from COVID-associated inpatient claims. This has its own separate units (as explained precisely in the methods section), and is thus given its own y-axis, on the right-hand side of the figure. Two versions of this inpatient signal are shown: a real-time signal, computed using claims that would have been available at each reference date on the x-axis, and a finalized signal, computed using claims that would have only been available 30 days after each reference date. All three series are smoothed over a trailing 7-day window. We can see that the finalized inpatient signal tracks reported hospitalizations quite well, however, the real-time signal is much more volatile, and its concordance with reported hospitalizations is much worse. (The real-time signal is also missing at some dates around the summer of 2021, which explains why no values are plotted there. We will discuss this shortly in the methods section.)

**Figure 1:**
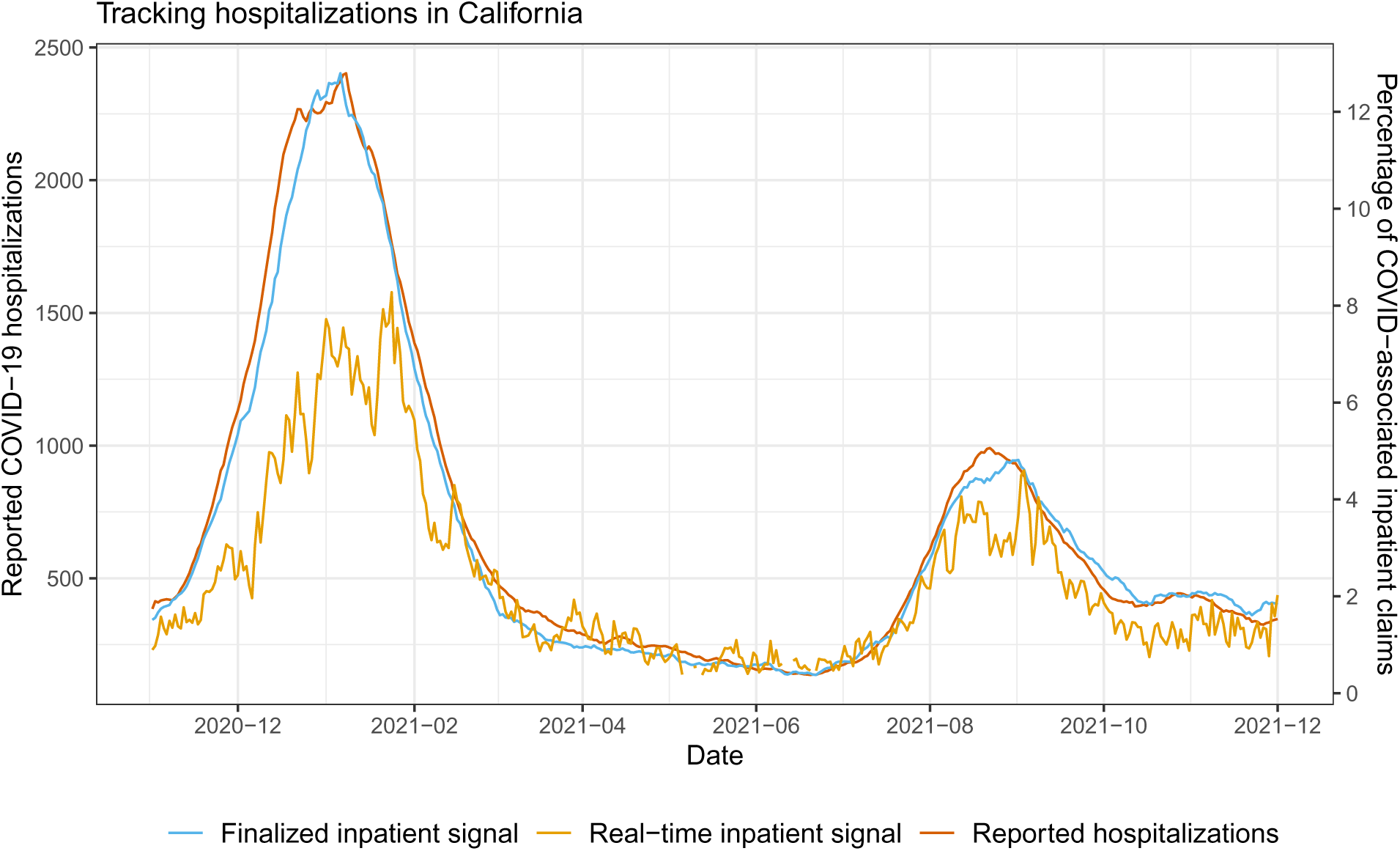
Reported daily COVID-19 hospitalizations, plotted alongside a signal derived from medical claims that measures daily COVID-associated inpatient admissions, for the state of California.

The importance of data versioning for epidemic tracking and prediction tasks is emphasized in Reinhart et al. (2021); McDonald et al. (2021), and the importance of leveraging existing healthcare data streams for epidemic surveillance, as a complement to traditional public health reporting, is motivated in Rosenfeld and Tibshirani (2021). Infectious disease nowcasting, using data from healthcare pipelines and also from a variety of other auxiliary data sources, has received increased attention over the last decade or so: see, e.g., Viboud et al. (2014); Smolinski et al. (2015); Yang et al. (2015); Farrow (2016); Santillana et al. (2016); Bastos et al. (2019); Jahja et al. (2019); Yang et al. (2019); Ackley et al. (2020); Brooks (2020); Leuba et al. (2020); Radin et al. (2020); Jahja et al. (2021); Li and White (2021); Menkir et al. (2021); Bergström et al. (2022); Miller et al. (2022); Seaman et al. (2022); Salazar et al. (2022); Wolffram et al. (2023); Lison et al. (2024). The goal in much of this work is to produce high-resolution, up-to-date estimates of disease activity for a fast-moving pathogen like influenza or COVID-19. The nowcasting problem is essentially to project finalized values from preliminary or partial measurements, making use of backfill or delay distributions, and possibly proxy signals which are exogenous to the main data reporting pipeline. The action of backfill or delay is naturally modeled via convolution, and Bayesian methods have been popular here, given their ability to streamline estimation and uncertainty quantification.

Our paper complements this line of work by considering the effectiveness of nowcasting based *purely* on proxy signals (for us, medical claims signals): we consider a hypothetical scenario in which hospital reporting on COVID-19 would have been set up at a coarser frequency, and under this hypothetical there is no partial or preliminary reporting data in the interm periods between reports (lasting a month, or longer) on which to base nowcasts. Finally, we examine intentionally simple statistical models based on regression, which would be relatively easy for a public health office to operationalize in practice.

## 2 Methods

This section describes the data that we use, the hypothetical scenarios (for hospital reporting cadence) that we consider, and the nowcast and backcast models that we build and evaluate. In describing these models, we also cover model training techniques which account for nonstationarity, time series cross-validation schemes for selecting tuning parameters, and variants of the basic (state-level) model which pool data across states. In the last subsection, we describe a nonparametric method for constructing prediction intervals.

### 2.1 Data

Throughout, we restrict our attention to nowcasting state-level hospitalizations. This is because the medical claims signals that we describe below are not generally available at each US county. That said, in principle, the same ideas we describe in what follows could be applied to nowcast hospital reports in large counties (subject to enough claims data being available in order to form robust signals). We also restrict our attention to nowcasting daily reported hospitalizations between April 1, 2021 and August 1, 2023, though we use data back until November 1, 2020 for training models.

State-level, daily COVID-19 hospitalization reports are obtained from the HHS, accessed via the Delphi Epidata API (Farrow et al., 2015; Reinhart et al., 2021). We use *Y_ℓ,s_* to denote the 7-day trailing average of *finalized* reported new COVID-19 hospitalization counts corresponding to location *ℓ* and time *s*. Averaging over 7-day trailing windows is mainly used as a smoother, and to account for weekday-weekend differences. Reported hospitalization counts are subject to revision (these are typically minor in comparison to revisions for claims signals), hence we also introduce notation to work with versioned data henceforth: we denote by *Y* ^(*t*)^ the 7-day trailing average of hospitalization counts for location *ℓ* and time *s*, but whose data version is *as of* time *t*. In this context, we often refer to *s* as the *reference date* and *t* the *issue date*. We use analogous notation and nomenclature for all versioned data in this paper.

De-identified, aggregated medical insurance claims data are provided to us by Change Healthcare, which covers around 25% of all commercial medical insurance claims in the US. Moreover, based on comparing total counts of COVID-19 hospitalizations from inpatient claims and those reported to the HHS (over the full period of our analysis), we find that there is a broad range: at the maximum end, for some US states, over 55% of hospitalizations are reflected in the claims data; at the minimum end, for other states, less than 20% appear in the claims data. (A more detailed analysis is given in Appendix A.) Nevertheless, we have enough claims data per state in order for us to be able to derive meaningful signals which reflect COVID-associated outpatient and inpatient activity. Specifically, in this work, we consider the following two signals:

- *O_ℓ,s_*: the *finalized* percentage of outpatient claims in a 7-day trailing window with a confirmed COVID diagnostic code, corresponding to location *ℓ* and time *s*.
- *I_ℓ,s_*: the *finalized* percentage of inpatient claims in a 7-day trailing window with a COVID-associated diagnostic code, corresponding to location *ℓ* and time *s*.

As before, we use *O_ℓ,s_*^(*t*)^ and *I_ℓ,s_*^(*t*)^ to denote the *versioned* outpatient and inpatient claims signals, respectively, corresponding to issue date *t*. The use of versioned data is extremely important when working with claims-based signals because medical insurance claims are often submitted and/or processed late, many days (and even months) after a given date of service. This process is generally referred to as *backfill*, and it has quite a pronounced effect on both outpatient and inpatient signals. We can look back at Figure 1 for an example of the inpatient signal in California. What is labeled as the “real-time inpatient signal” in the figure is *I*^(*s*)^ in our notation introduced here, for *s* ranging over the time values along the x-axis (and *ℓ* = California). What is labeled as the “finalized inpatient signal” is actually *I*^(*s*+30)^; note that 30 is just a large number chosen for simplicity, and it is not necessarily the case that all claims would have been filed after 30 days.

Sometimes backfill is so severe that no claims for reference date *s* are filed at all until a later issue date *t*, which we refer to as *latency*. If the latency is large enough, in particular, if no claims are available until a full 7 days after a given reference date, then the real-time claims signal value will be missing. This happens at a few dates in Figure 1 in May and June of 2021.

In defining the inpatient and outpatient claims signals, the use of a ratio of COVID-associated claims to all claims, instead of a count of COVID-associated claims, is important for two reasons. First, it adjusts for the unknown market share (unknown to us) of Change Healthcare in each given location. Second, this ratio tends to be more robust to backfill than a pure count, as both the numerator and denominator get updated as new claims are filed (whereas a count would only be revised upward, and as we will see later in Figure 2, especially for the inpatient signal, only a small fraction of the total volume of claims are available in the first few days after a given date of service).

**Figure 2:**
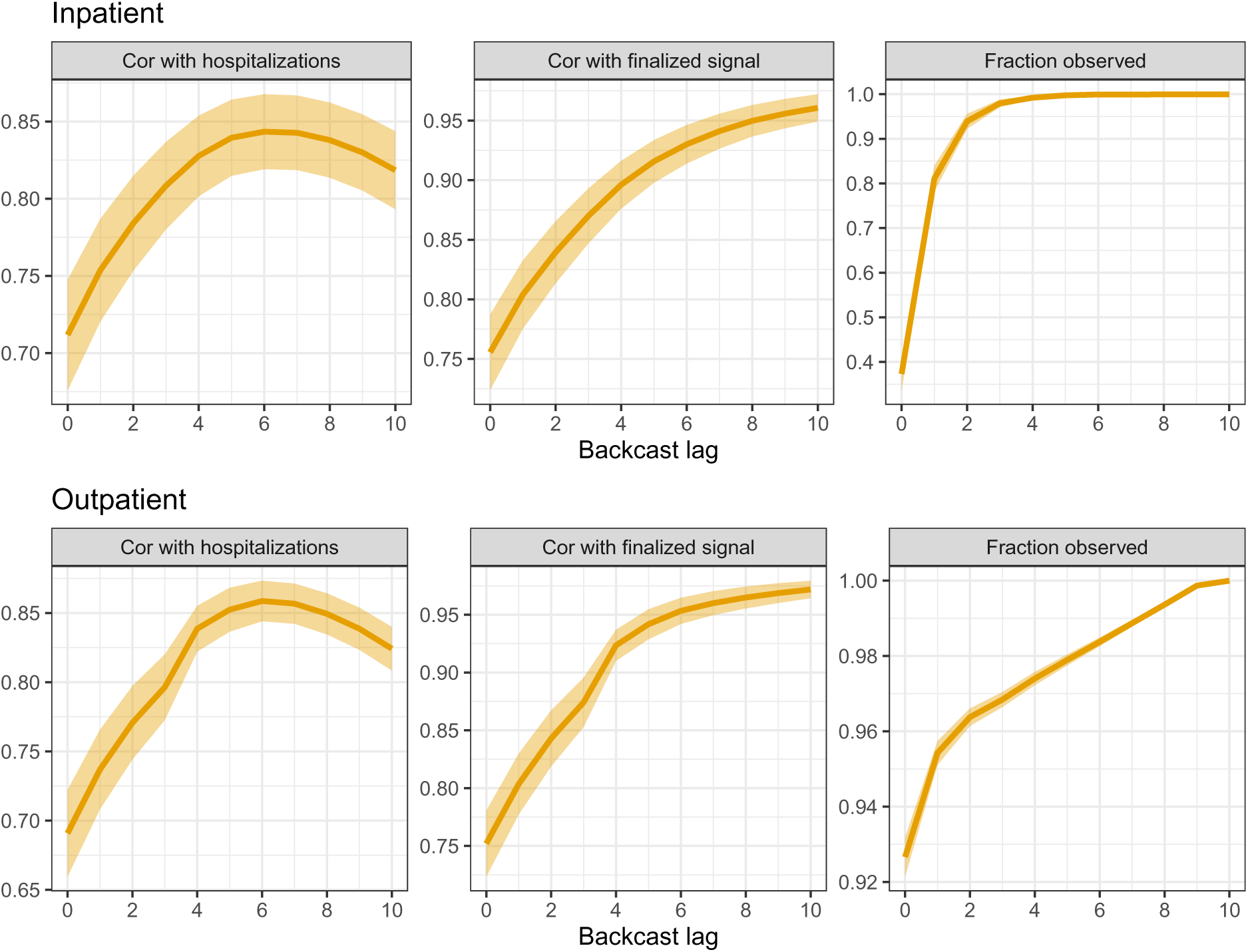
Analysis to help guide lag selection for inpatient and outpatient features in the working model (1). The rows correspond to different features, and the columns to different metrics, as explained precisely in the main text. The shaded regions show ±1 standard error bands for each metric, over the state averages.

Like the HHS hospitalization reports, various signals derived from medical insurance claims are available in the Delphi Epidata API. The precise diagnostic codes used in defining the outpatient and inpatient signals are given in the API documentation: https://cmu-delphi.github.io/delphi-epidata. For convenience, we have relayed these definitions in Appendix A; we have also made all data (including properly-versioned data) used in our analysis available for download at: https://github.com/cmu-delphi/hhs-nowcasting.

### 2.2 Hypothetical scenarios

We consider two hypothetical scenarios, described next. Recall that our nowcasting evaluation spans April 1, 2021 to August 1, 2023, and we have training data all the way back until November 1, 2020.

#### Scenario 1: monthly updates

Our first hypothetical scenario examines nowcasting from April 1, 2021 to November 30, 2021. In this scenario, on the first day of each month during this period, we receive daily hospitalization counts for the previous month, and we nowcast and backcast the (unobserved) hospitalization counts for each following day in the given month, using the outpatient and inpatient claims signals described above. Note that the period in this scenario covers the Delta wave in the US. The start of this hypothetical scenario is chosen to be April 1, 2021 so that we have a large enough “burn-in set” (initial training set), which extends back to November 1, 2020. (This includes the winter wave of 2020, which helps the initial models capture relationships during a time of dynamic change.)

To make this all more precise, let us introduce some notation. We drop reference to the location *ℓ* here and in what follows, whenever convenient (whenever it is not needed for the given explanation). Let *t*_0_ be a date that marks the start of a month during the period of April 1, 2021 to November 30, 2021. Then on each day *t* = *t*_0_*, t*_0_ + 1*, …, t*_1_ − 1, where *t*_1_ marks the first day of the next month, we have access to:

- {*Y* ^(*t*0)^}*_s_*_≤*t*−1_, hospitalization counts through day *t*_0_ − 1, with versions as of day *t*_0_; and
- {(*I*^(*t*)^*, O*^(*t*)^)}, outpatient and inpatient signals through day *t*, with versions as of day *t*.

On each such day *t*, we use the data we have to train a regression model, call it *f_t_*, to predict hospitalizations from claims signals. We use this to make nowcasts:

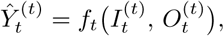

and lag-*k* backcasts:

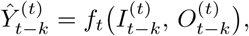

for each *k* = 1, …, 10. (Clearly, a nowcast is equivalent to a lag-0 backcast.) This is repeated for each of the 9 months in the period spanned by this monthly-update scenario. The details of how regression models are trained and evaluated will be given in the next subsection.

#### Scenario 2: no updates

Our second hypothetical scenario spans nowcasting dates from December 1, 2021 to August 1, 2023. In this scenario, we receive reported hospitalizations up through November 30, 2021 with versions as of December 1, 2021, and we receive *no further* hospitalization counts after that. As before, we nowcast and backcast the (unobserved) hospitalization counts in the remaining period using the outpatient and inpatient claims signals. Note that the period in this scenario covers the Omicron wave in the US, and that this is second scenario is generally far more challenging than the first scenario.

The data received and nowcasts and backcasts made in the no-update scenario can be written in precise notation, in fact, exactly as introduced in the description for the monthly-update scenario above, but now we fix *t*_0_ at December 1, 2021, and make nowcasts and backcasts at each *t* from *t*_0_ through the end of the period, August 31, 2023. The details of how regression models are trained and evaluated in this scenario will again be covered in the next subsection.

### 2.3 Regression model

As a basic working model, we predict hospitalizations using a linear combination of a set *L^I^* of lags of the inpatient signal and a set *L^O^* of lags of the outpatient signal,

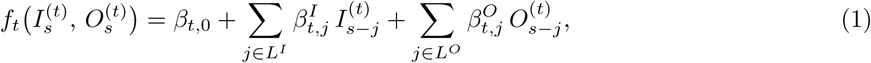

where the coefficients *β_t,_*_0_, *β^I^*, and *β^O^* are estimated, i.e., the model *f_t_* is fit, by training on historical data available at time *t*, either separately for each location *ℓ* (recall, the notational dependence on the location has been dropped for now), or in a way that pools data across locations. Details will be given below. First, we describe how we select the lag sets *L^O^*, *L^I^* for the inpatient and outpatient features.

#### 2.3.1 Selecting feature lags

To select the lag sets *L^O^*, *L^I^* for the working model (1), we restrict our attention to the burn-in set, before the first nowcast date of April 1, 2021 (so as to avoid overfitting to the data in our nowcasting period and reserve this for proper evaluation of our models in the two scenarios outlined above).

To guide lag selection, we consider three metrics, which measure predictive power, feature stability, and data availability. Let *X*^(*t*)^ denote a generic feature in our usual notation for versioned data, i.e., *X*^(*t*)^ = *I*^(*t*)^ for the inpatient feature or *X*^(*t*)^ = *O*^(*t*)^ for outpatient feature, we consider, as a function of lag *j*:

- correlation of *X*^(*t*)^ with finalized hospitalizations *Y_t_*, which is a measure of predictive power;
- correlation of *X*^(*t*)^ with its own finalized value *X_t_*_−*j*_, which is a measure of stability;
- the fraction of total claims used to compute the finalized signal *X_t_*_−*j*_ that are observed by time *t*.

(Note: the finalized signal values would not have been available at the end of the burn-in period. However, this does not pose any additional risk to overfitting the data in the nowcasting period, which was the reason to separate out the burn-in set in the first place.) Each metric is computed over time: between November 1, 2020 and March 31, 2021, for each location. The results are displayed in Figure 2, averaged over all locations (all states). The shaded regions display ±1 standard error bands, computed over the state averages.

These metrics exhibit a tradeoff as a function of the lag *j*. The first is nonmonotone: is increases at first because the claims signal at a larger lag *j* is less volatile (more total claims observed), but then decreases eventually because the signal reference date *t* − *j* is less related to hospitalizations at *t*. The second and third metrics are monotone in *j*. Altogether, we can see that lag 6, for each of the inpatient and outpatient signals, offers a nice balance across the three metrics: maximum predictive power (achieving the highest correlation between hospitalization for both groups of signals), high stability and availability. We therefore include lag 6 in each of the sets *L^I^*and *L^O^*. To improve robustness and capture longer-range dependencies, we also include lags 13 and 20 in each of *L^I^* and *L^O^*, which is roughly consistent with choices of feature lags in other basic epidemic prediction models (e.g., McDonald et al. (2021)). The choice of 7-day spacing here is also motivated by the desire to limit correlations between features in (1); recall that the inpatient and outpatient signals are computed using a trailing 7-day window of claims data.

#### 2.3.2 Training the regression model

At each nowcast date *t*, we fit the coefficients *β_t,_*_0_, *β^I^* in (1) by solving the following (weighted) least squares optimization problem:

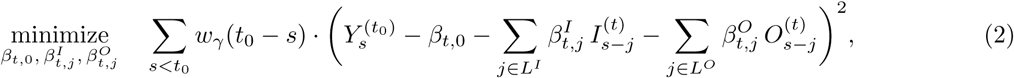

where we use exponentially decaying observation weights:

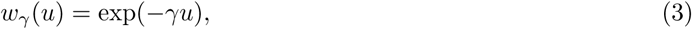

for a tuning parameter *γ* ≥ 0. The index *t*_0_ in (2) marks the latest observation boundary for hospitalization reports before time *t*: this is either the start of the month in scenario 1 (where we receive monthly updates), or December 1, 2021 in scenario 2 (where we receive no further updates). Hence, in other words, we fit the coefficients in (2) by minimizing the weighted mean squared error of our working regression model, over all dates at which response values (reported hospitalization counts) are available. Note carefully that in (2) we only use properly-versioned data that would have been available at *t*. If any such data, response or feature values, are missing at time *t* then the corresponding summand is simply omitted from (2).

As a default, we fit the model by solving (2) separately for each location *ℓ* (recall, we have suppressed the dependence on *ℓ* in the notation for simplicity). We call this the *state-level* model. Later, we discuss schemes for pooling training data across locations. Next, we focus on the decay parameter *γ* in (2), (3).

#### 2.3.3 Selecting the decay parameter by cross-validation

Before describing how we select *γ* in the exponential weights (3) that are used in the weighted least squares problem (2), we pause to discuss the question: why is it useful to use decaying observation weights in the first place? The reason is because the the features—lagged versions of the inpatient and outpatient medical insurance claims signals, and response—reported hospitalizations, need not be jointly stationary, i.e., their relationship may be changing over time.

Looking back at Figure 1, note that we observe evidence of nonstationarity in the relationship between the real-time inpatient signal and reported hospitalizations. If we were to regress reported hospitalizations on the inpatient signal alone, then the regression coefficient that would be appropriate for the period April–July 2021 would be too small for the first hospitalization wave starting in December 2020 (and to a lesser extent, also too small for the second wave starting in August 2021).

By allowing *γ* itself change over time, we can adapt to the degree of nonstationarity at any point in time, i.e., we can adapt to the amount of past training data that is relevant for the current prediction. Indeed, we will select *γ* in a dynamic, time-varying fashion using cross-validation (CV). Given the sequential nature of our prediction problem, we use a version of CV that is purely forward-looking, and is sometimes referred to as *time series cross-validation* in the literature (Hyndman and Athanasopoulos, 2021). This works as follows. Let *t*_0_ denote the index that marks the most recent observation boundary for hospitalization reports, and let *t*_−1_*, t*_−2_ denote the indices that mark the previous two observation boundaries before *t*_0_. For each *γ* in a grid Γ of tuning parameter values, we carry out the following procedure:

- for each *t* ∈ [*t*_−2_*, t*_−1_):

**–** fit a regression model by solving:

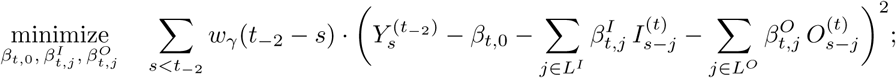

**–** produce backcasts *Y*^^(*t*)^, *s* ∈ [*t*_−2_*, t*];

- for each *t* ∈ [*t*_−1_*, t*_0_):

**–** fit a regression model by solving:

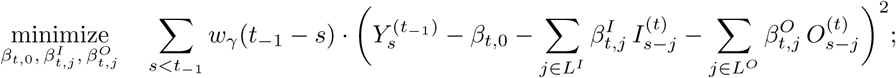
**–** produce backcasts *Y*^^(*t*)^, *s* ∈ [*t*_−1_*, t*];
- compute the mean absolute error (MAE) of all backcasts made at all times *t* ∈ [*t*_−2_*, t*_0_), against reported hospitalization counts *Y* ^(*t*0)^, *t* ∈ [*t*_−2_*, t*_0_).

In other words, this procedure uses the last 2 months of data as a validation set for tuning *γ*. Ultimately, we choose *γ* ∈ Γ that minimizes the MAE computed in the last step above. To be clear, after choosing *γ* in this way, we then fit the model by solving (2) and use this to make backcasts at each time *t* ∈ [*t*_0_*, t*_1_), where *t*_1_ is the observation boundary after *t*_0_.

Toward defining the tuning parameter set Γ, we first compute *γ*_max_, the value of *γ* such that the effective sample size of the weight sequence equals 30, i.e., it solves

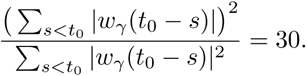

This is chosen as a heuristic upper bound on the “reasonable” range of *γ* values, motivated by the idea that restricting to at least (effectively) 1 month of training data helps to avoid fitted models which are too volatile. We then set Γ to contain 25 evenly-spaced values between 0 and *γ*_max_.

#### 2.3.4 Stabilizing predictions by geo-pooling

To borrow strength across locations, we consider fitting the regression model at each time *t* by pooling data across locations, which we refer to as the *geo-pooled* model. To be precise, instead of solving (2) per location *ℓ*, here we instead solve:

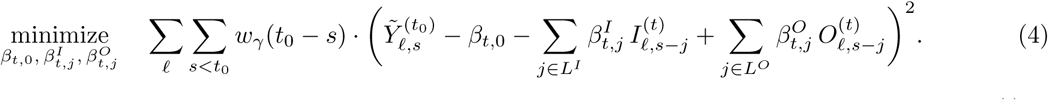

Note that in (4), the training set includes data from all locations *ℓ*. Importantly, in (4), the response *Y*^^^(*^t^*^)^ is the *hospitalization rate* in location *ℓ* at reference date *s*, as of time *t*, which is defined as the hospitalization count per 100,000 people, i.e.,

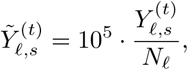

where *N_ℓ_*is the population of location *ℓ*. The use of rates rather than counts in the pooled regression (4) is critical, because otherwise the coefficients would have different meanings in different locations, and pooling across locations would not make sense. After solving (4), in order to use the fitted model to make predictions of hospitalization counts at a given location *ℓ*, we would then need to rescale the predictions by *N_ℓ_/*10^5^.

Finally, we also consider a *mixed* model, which linearly combines the state-level *Y*^^(*t*),state^ and geo-pooled *Y*^^(*t*),pooled^ predictions, via

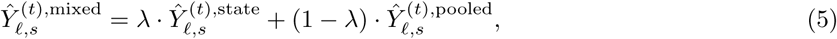

for a mixing parameter *λ* ∈ [0, 1]. We select *λ*, separately per state, using the same cross-validation strategy described previously, where we tune *λ* over a grid of 50 evenly-spaced values between 0 and 1. (We tune over *γ* separately for each of the state-level and geo-pooled models, and then tune over *λ*.)

### 2.4 Prediction intervals

In addition to considering point predictions, from the models described above, we use residuals from these models and *quantile tracking* (Angelopoulos et al., 2023) to generate prediction intervals. Quantile tracking is a method from the online conformal literature, and produces nonparametric intervals which are guaranteed to attain long-run coverage over an arbitrary bounded data sequence (including nonstationary time series). Abstractly, given a predicted value *Y*^^^*_t_* of some unobserved target value *Y_t_* at time *t*, we begin by specifiying a score function *φ* (assumed to be negatively-oriented, so that lower values correspond to better accuracy) and we construct a prediction set C*_t_* at time *t* via

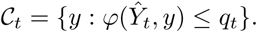

Here *q_t_* denotes a parameter that is output by the quantile tracking algorithm. Given a nominal coverage level 1 − *α*, the parameter *q_t_* is updated using an online gradient descent step with respect to an optimization problem defined by summing the level 1 − *α* quantile losses of *φ*(*Y*^^^*_t_, y*) − *q_t_* over the sequence *t* = 1, 2, 3, ….

In our problem, we do not receive new target values after we make each prediction, unlike the standard online learning setup used in Angelopoulos et al. (2023), so we adapt their method so that *q_t_* is adjusted only at the observation boundaries in scenario 1. Further, to allow our intervals to exhibit varying-width between observation boundaries, we use a scaled score that divides the residual by the value of the prediction. Finally, in order to allow our intervals to be asymmetric around the predicted value, we define a separate *lower* and *upper* score, *φ^l^*and *φ^u^*, respectively:

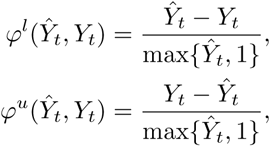

and effectively run two instances of quantile tracking, with parameters *q^l^* and *q^u^*, respectively, each at the level 1 − *α/*2. Combining the results from these two procedures then gives us the lower and upper endpoints of the prediction intervals.

We are now ready to describe our adaptation of quantile tracking (under batched updates). In the same notation as that used to describe fitting the regression models above, let *t*_0_ denote the the latest observation boundary, and *t*_1_ denote the next observation boundary. Suppose we are making backcasts at lag *k* (with *k* = 0 representing nowcasts). For each *t* ∈ [*t*_0_*, t*_1_), we:

- produce a point prediction *Y*^^(^*^t^*) using one of the regression models described in previous subsections;
- produce a prediction interval via

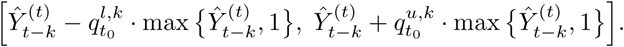

Then, at *t* = *t*_1_, we:

- look back and compute one-sided coverage errors,

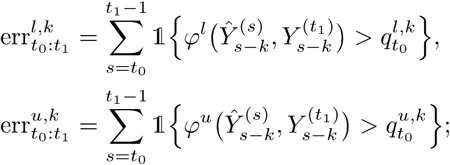
- update each of *q^l,k^*, *q^u,k^* by taking appropriate gradient descent steps,

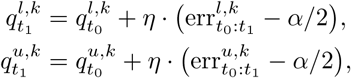

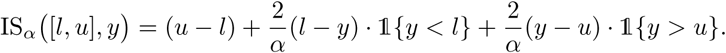

where *η >* 0 is a small fixed learning rate.

To prevent crossing of quantiles, we enforce *q^l,k^_t1_* ≥ 0 and *q^u,k^_t1_* ≥ 0 (by clipping them at zero if a gradient descent update were to make them negative).

## 3 Results

This section examines the nowcasting and backcasting results in scenario 1 (monthly-update period), and scenario 2 (no-update period), through quantitative backcast error analysis and qualitative inspection of the nowcast dynamics over time. We then perform an ablation study, to tease apart how individual parts of the model relate to overall accuracy, and finish by analyzing the prediction intervals.

### 3.1 Scenario 1: backcast error analysis

Figure 3 shows the MAE of all backcasts, as a function of lag *k* = 0, …, 10, from the state-level, geo-pooled, and mixed models over the monthly-update period (which, recall spans April 1, 2021 to November 30, 2021). To be clear, for each model, this is computed by averaging the absolute error of backcasts made to *finalized* hospitalization counts, per state. The left panel displays the average of these state MAE values, and as well as standard error bands. The right panel is similar but normalizes the state MAE values by state population times 10^5^, thus considering MAE on the scale of hospitalization rates (rather than counts).

**Figure 3:**
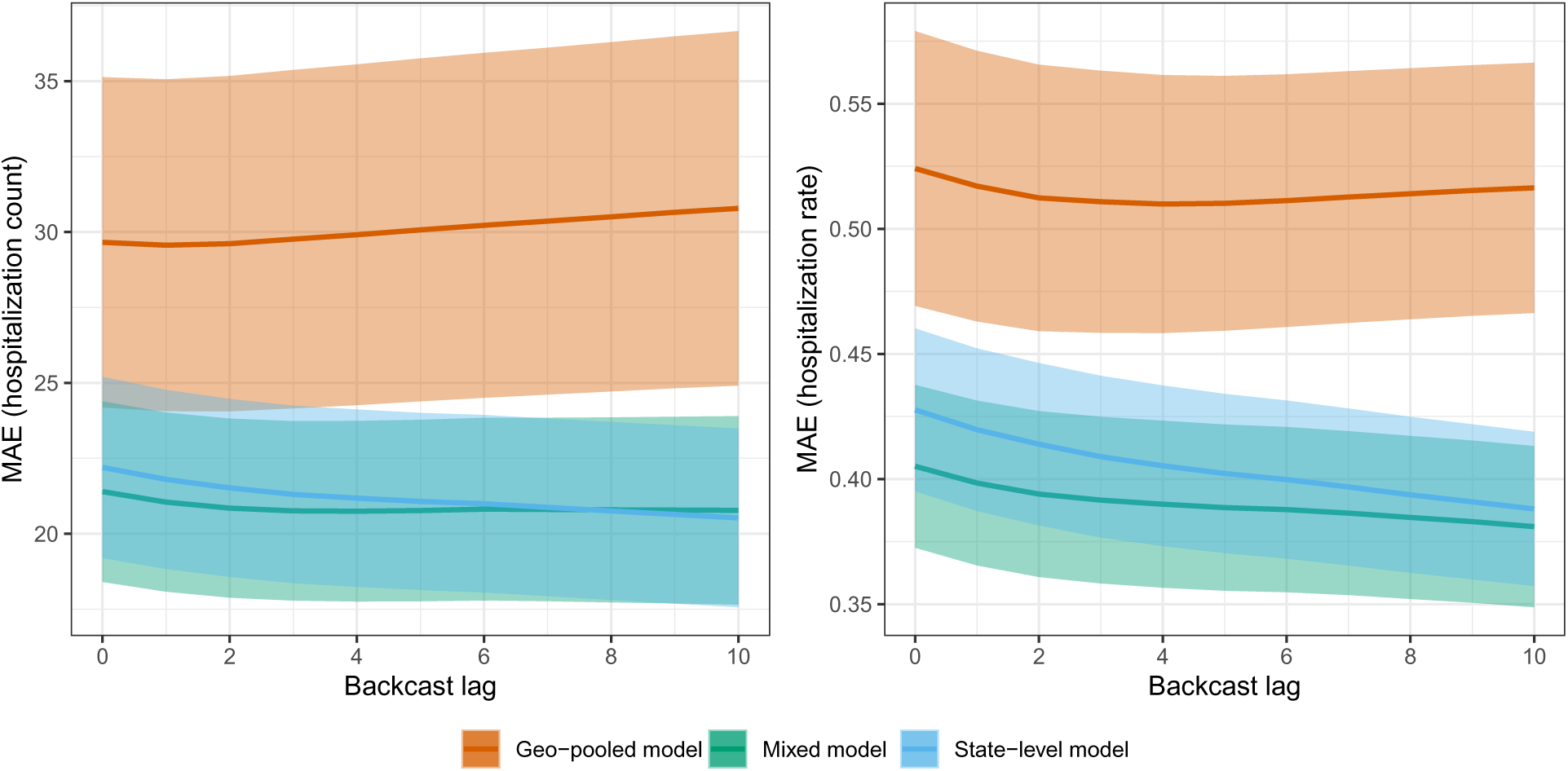
MAE as a function of backcast lag for the state-level, geo-pooled, and mixed models in scenario 1, the monthly-update period. The shaded regions show ±1 standard error bands, over the state MAE values.

The geo-pooled model is clearly worse in terms of MAE than the mixed and state-level models. On the counts scale (left panel), the state-level model has slightly better MAE than the mixed model; on the rates scale (right panel), the opposite is true. This is because the mixed model generally performs better for smaller states, where predictions tend to be more volatile and shrinking toward the geo-pooled model helps to reduce variance. This is not reflected in the MAE plot on the left panel, since poor backcast performance on small states contributes little when measured in terms of counts.

In absolute terms, the backcasts made by the state-level and mixed models are generally quite accurate. For example, the nowcasts (backcasts at lag 0) made by the state-level and mixed models have MAEs of 0.428 and 0.405, respectively, on the scale of hospitalization rates. These correspond to proportions of variance explained (PVEs) of 71.6% and 75.4%, respectively. Even the geo-pooled model, which is relatively quite a bit worse, produces nowcasts with an MAE of 0.524 on the scale of hospitalization rates, which corresponds to a still respectable PVE of 61.1%.

### 3.2 Scenario 1: illustrative nowcast examples

We examine nowcasts made by the state-level and mixed models during the monthly-update period, in four states: California (CA), Kentucky (KY), Vermont (VT), and New York (NY). The first three are chosen to demonstrate the qualitative behavior of nowcasts in states of different sizes, with CA having the largest population, KY having roughly median population, and VT having the smallest. NY is chosen because it represents somewhat of failure case for the robustness of nowcasts from the state-level model.

Figure 4 displays state-level and mixed model nowcasts for CA, KY, and VT. To be clear, in each panel of the figure, the nowcasts use *real-time* inpatient and claims signals as predictive features, and the models behind these nowcasts are trained using hospitalization data up to the latest observation boundary (marked by dotted vertical lines). The shaded bars in the figure display *finalized* reported hospitalizations, the target of ultimate interest. In CA and KY, where the Delta wave is prominent (roughly July to November), we can see that both the state-level and mixed model nowcasts track the dynamics of the Delta wave. For example, looking at July 1, 2021 in CA, the hospitalization reports from the previous months (which is all that would have been available at that time) show no indication of an upswing to come. Still, the nowcasts during July present a clear upward trend, which means that policy-makers would know from these nowcasts that a wave is underway. As we see it, evaluating nowcasts for their qualitative shape (in comparison to hospitalization waves) is important—just as much as (if not more so than) numerical error analysis.

**Figure 4:**
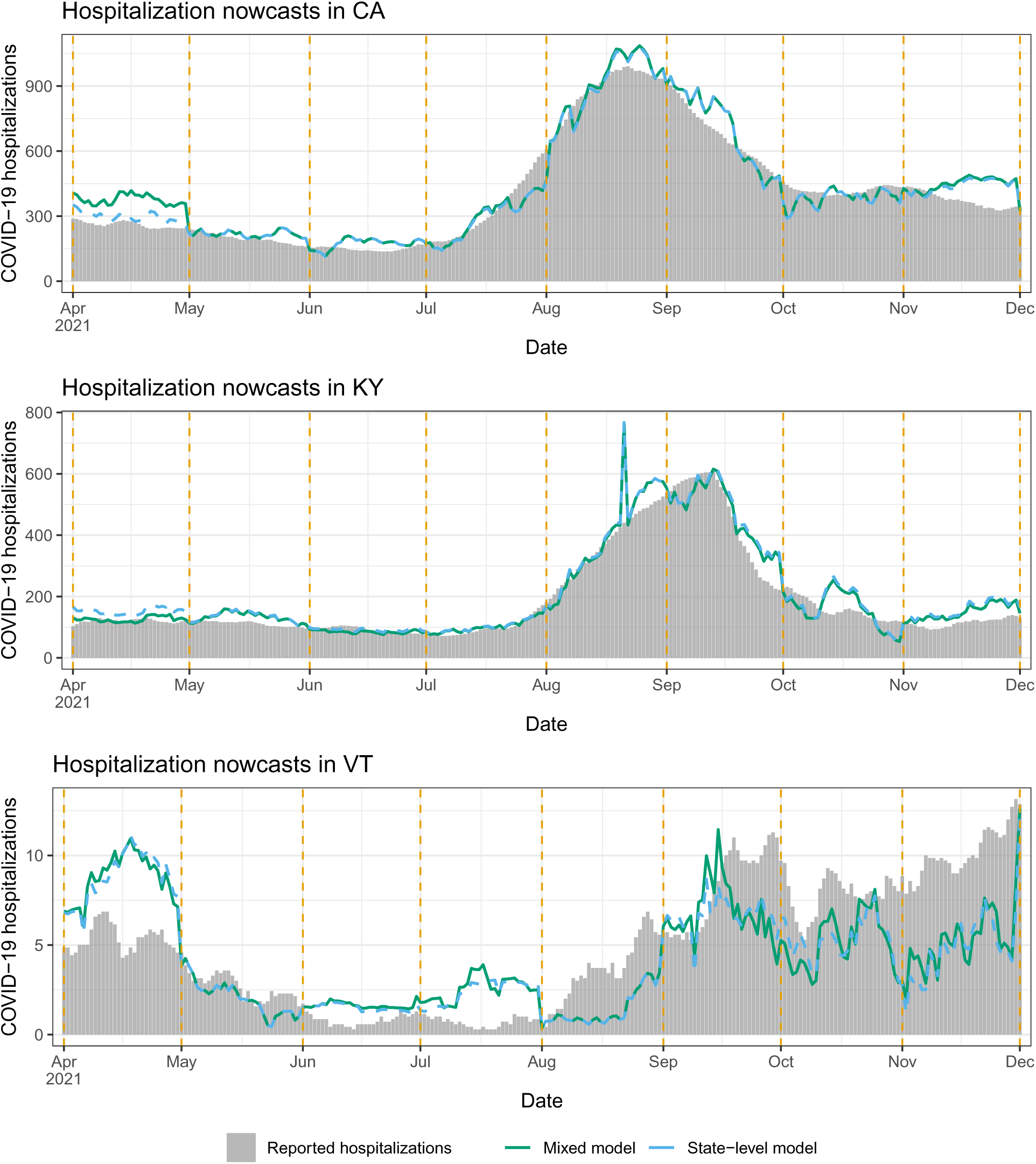
Nowcasts from the state-level and mixed models in scenario 1, the monthly-update period, for CA, KY, and VT. The dotted vertical lines mark the observation boundaries—when hospitalization reports for the previous month are received, and models are retrained.

Being much smaller, VT has on the order of ten reported COVID-19 hospitalizations daily, not several hundreds, as in KY or CA. Accordingly, we can see in Figure 4 that the (finalized) reported hospitalizations curve in VT is much noisier—we do not really see a clear Delta wave (instead, an increasing but quite noisy trend from August through December). The claims signals are also more noisy in a small state like VT, since they are ratios of small counts. Therefore, both the response and features are more volatile in the prediction problem for VT, and not surprisingly, the nowcasts here appear qualitatively worse. That said, the nowcasts still roughly track the gross trends in reported hospitalizations: higher in April and May, lower in June and July, and growing again in August onward.

Generally, the discussion above translates to the rest of the US: nowcasts tend to be in good qualitative agreement with hospitalization waves in larger states, and less so in smaller states. Appendix C provides a full set of nowcast and backcast plots in all 50 states, from the mixed model, under scenario 1. Occasionally, we find that the trend in the predictions disagrees with that in hospitalization reports in a given month, but this gets corrected at the next observation boundary, once new training data is available and the regression model is refit. We also find that backcasts at lag 5 or 10 tend to be smoother than nowcasts.

Figure 5 shows state-level and mixed model nowcasts for NY. This is a notable example of a failure in robustness of the state-level model: its nowcasts for a good part of the month of June are actually *negative* (and we truncate them at zero for visualization purposes). This is due to a large, systematic revision which occurred on June 8 in the outpatient signal, where its values for reference dates in May were revised upward. Moreover, when the state-level model is fit to data through the end of May, the regression coefficient on the largest outpatient feature lag ends up being negative, and consequently the upward revision of past feature values on June 8 introduces a strong downward bias in the subsequent predictions. Supporting analysis for this explanation is given in Appendix B.

**Figure 5:**
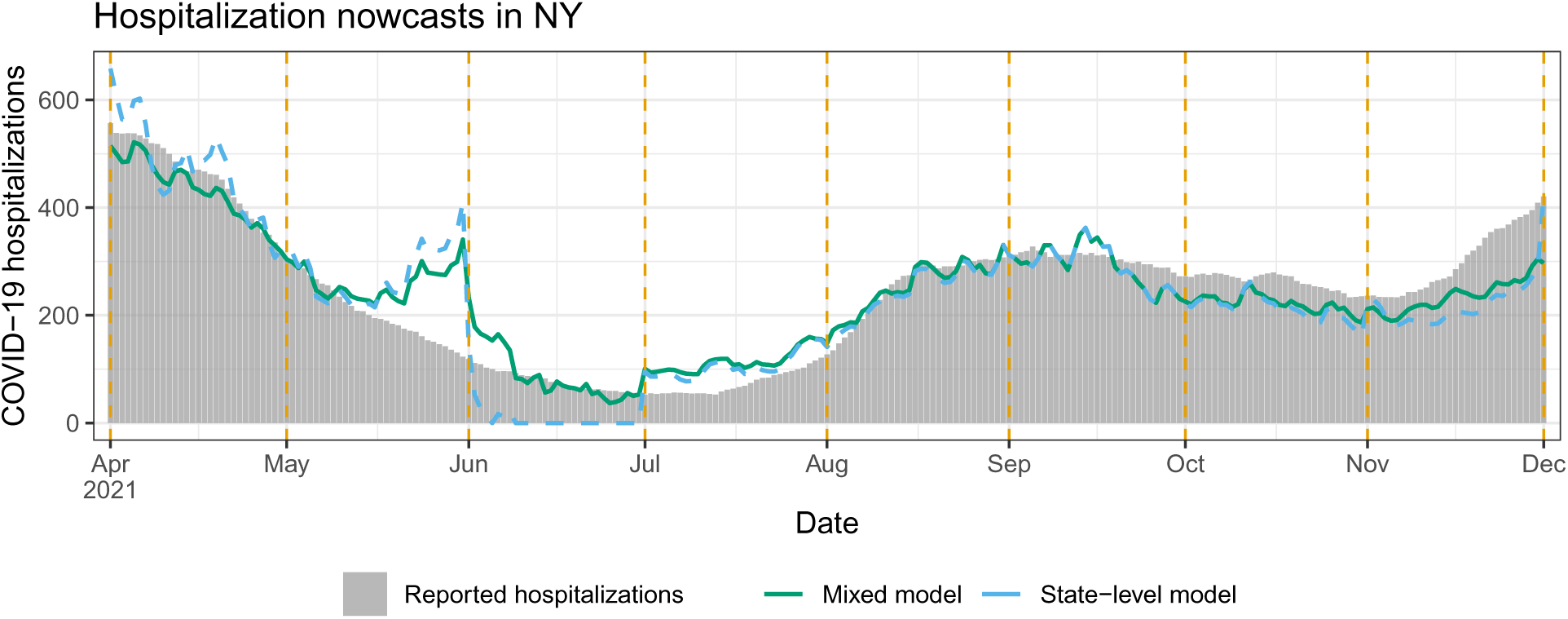
Nowcasts from the state-level and mixed models in scenario 1, the monthly-update period, for NY. The dotted vertical lines mark the observation boundaries, as in Figure 4.

As we can see in the figure, the mixed model produces nowcasts that are much more stable in the month of June. However, the mixed model and even the geo-pooled model are themselves not immune to large and erratic revisions in the input features at prediction time. These revisions can result in erratic nowcasts and backcasts. To mitigate this, we could turn to models that fuse information across a wider variety of auxiliary signals (where ideally, some of these signals would have less severe backfill than claims-based signals), an idea that we return to in the discussion section.

### 3.3 Scenario 2: backcast error analysis

Figure 6 shows the MAE of all backcasts, as a function of lag *k* = 0, …, 10, from the state-level, geo-pooled, and mixed models over the no-update period (spanning December 1, 2021 to August 31, 2023). The format is as in Figure 3. We see two notable differences compared to the results in the monthly-update period. First, as expected, each model performs worse than it did in Figure 6. On the scale of hospitalization rates (right panel), the range of MAEs has jumped from (roughly) 0.4–0.53 in the monthly-update period to 0.62–0.72 in the no-update period, and correspondingly, the PVEs have dropped from (roughly) 61–75% to 10–37%.

**Figure 6:**
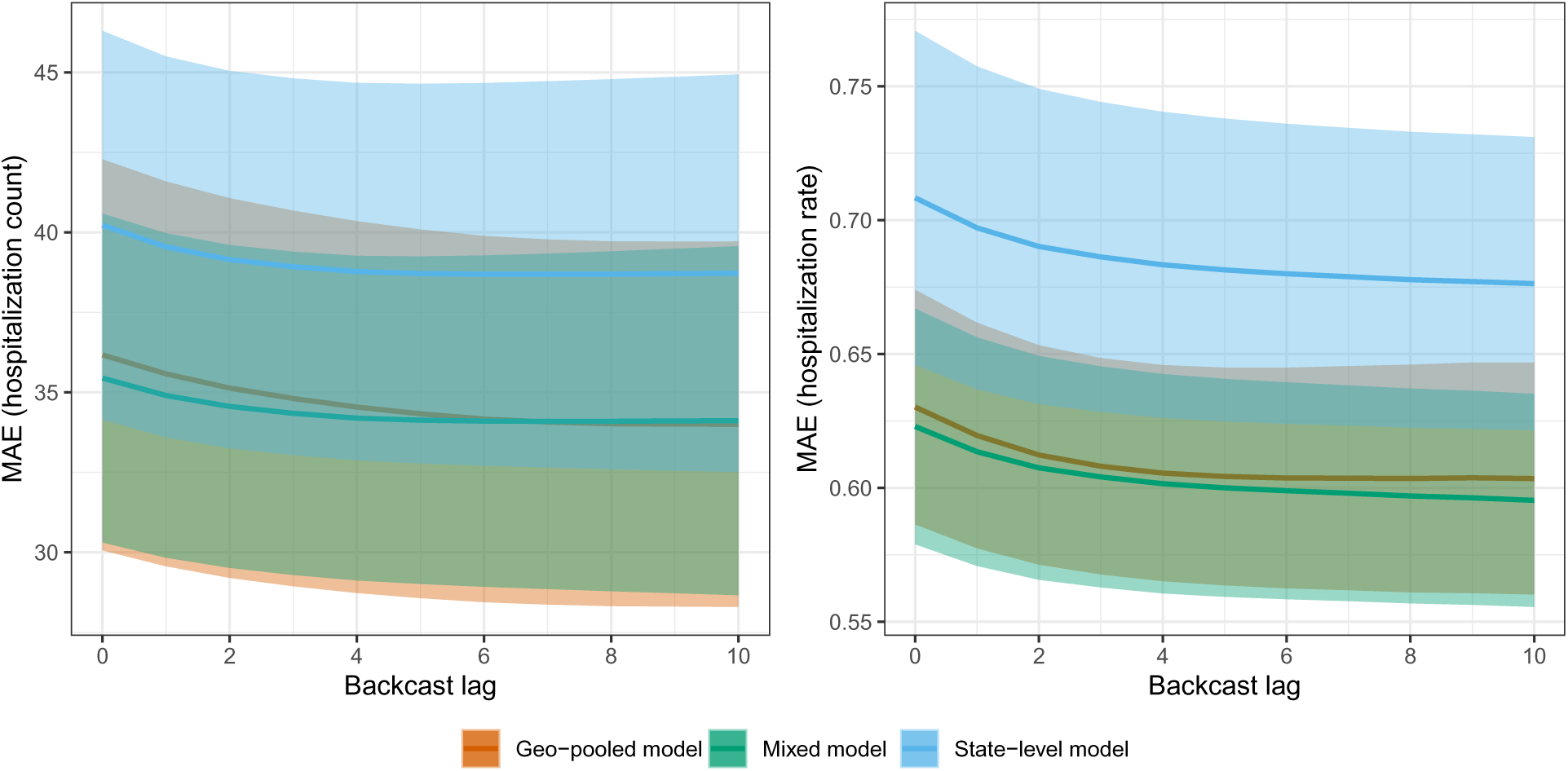
MAE as a function of backcast lag for the state-level, geo-pooled, and mixed models in scenario 2, the no-update period. The shaded regions show ±1 standard error bands, over the state MAE values.

Second (and perhaps a bit more surprising), we see in the no-update period that the state-level model performs clearly worse than the geo-pooled and mixed models when MAE is measured either on the counts or rates scale. The mixed model MAE is somewhat better than the geo-pooled model MAE with respect to counts, while the two are basically the same with respect to rates. It is encouraging to see that the mixed model performs competitively across both monthly-update and no-update scenarios: this model—equipped with the CV procedure for tuning *λ*—is able to effectively adapt the strengths of local and global training approaches (underlying the state-level and geo-pooled models) to the task at hand, in order to yield accurate predictions in an average-case sense.

### 3.4 Scenario 2: illustrative nowcast examples

We examine nowcasts made by the geo-pooled and mixed models for CA, KY, and VT. NY, which served as failure case in the monthly-update period, is not shown here, because KY itself provides such an example: as we will see, the mixed model lacks robustness over a part of the Omicron wave (meanwhile, the mixed model performs fine for NY throughout the no-update period—as shown in the appendix).

Figure 7 displays the nowcasts for CA, KY, and VT, with the same general format as in Figure 4. CA is a clear success case: both geo-pooled and mixed models capture the dynamics of the Omicron wave faithfully, and also track the summer 2023 and winter 2023 waves, despite (recall) receiving no reported hospitalizations past December 1, 2022. VT, as in the monthly-update period, is a challenging case because it corresponds to much noisier prediction problem, and the nowcasts here look qualitatively worse overall. However, the mixed model nowcasts still pick up the Omicron wave. KY represents a failure case: the mixed model nowcasts for all of February are *negative* and truncated at zero for the visualization. What this is actually demonstrating is a failure of the state-level model, and simultaneously, a failure of tuning of the mixing parameter *λ*. The mixed model here ends up placing a large weight on state-level predictions (not shown), which are themselves volatile for reasons similar to what happens in NY during the monthly-update period—large revisions to the input features at prediction time.

**Figure 7:**
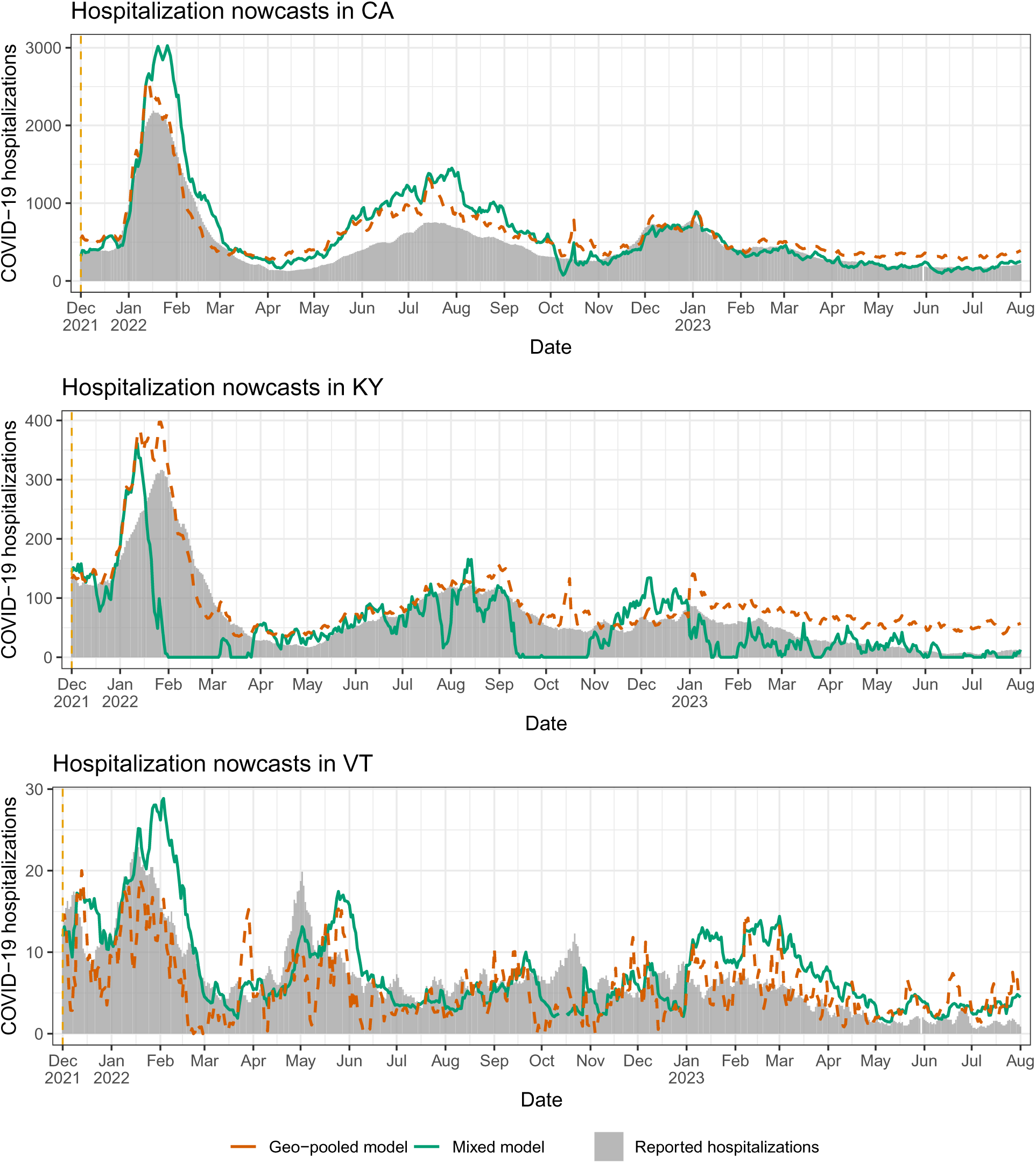
Nowcasts from the geo-pooled and mixed models in scenario 2, the no-update period, for CA, KY, and VT. The dotted vertical line (on the left side of the plot) marks the sole observation boundary in this period—hospitalization reports for all dates prior to this are available, but no reports are received after that.

The volatility of the mixed model nowcasts in KY during Omicron provides an important perspective: in the MAE sense, we found in Figure 6 that the mixed model (with its CV tuning for *λ*) successfully navigates the strengths and weaknesses of the geo-pooled and state-level models; and yet for specific states and periods of time, the mixed model can still lack robustness. To be clear, such fragility is not limited to the no-update period, and it could have happened in NY in the monthly-update period, had the CV tuning procedure for *λ* not downweighted the state-level predictions so heavily. Improving robustness is a direction for future work, and we revisit this topic in the discussion section.

Lastly, in Appendix D, we again provide a full set of nowcast and backcast plots in all 50 states, from the mixed model, under scenario 2. For larger states, we find that the nowcasts generally trace out the Omicron wave, but for smaller states, this happens less consistently and the nowcasts look more noisy. Backcasts at lag 5 or 10 tend to smooth out the nowcasts, though not dramatically.

### 3.5 Ablation study

To examine the importance of some of our modeling choices (as described in the methods section), we carry out an ablation study in which we remove a particular component of the model, use a simpler alternative in its place, and evaluate the result in terms of MAE. In particular, we consider four ablated models:

1. Unweighted, all past: we fit the regression model (2) without observation weights, on all past reported hospitalization data available (considering summands *s < t* in (2)).
2. Unweighted, two months: we fit the regression model (2) without observation weights, on the reported hospitalization data available for the latest two months (considering summands *s* ∈ [*t*_−2_*, t*_0_) in (2)).
3. Weighted, inpatient only: we fit the regression model (2) using only the lags of the inpatient signal.
4. Weighted, outpatient only: we fit the regression model (2) using only the lags of the outpatient signal.

Tables 1 and 2 compare the performance of the state-level model to these ablated models, for scenarios 1 and 2, respectively. In each scenario, for each model, we compute its MAE over the range of time values in the scenario, per backcast lag *k* = 0, …, 10 and state, then we report the average and standard error of these MAE values across all backcast lags and states. The state-level and inpatient-only models perform the best overall, and make up the two best models in either scenario. Meanwhile, the all-past model is slightly worse and the two-month model is significantly worse, which emphasizes the importance of decaying weights (and CV tuning) as part of the original model design. The outpatient-only model is competitive in scenario 1 but not in scenario 2. This could be a reflection of distribution-drift in the relationship between the outpatient signal and hospitalization reports post Omicron, which in scenario 2 is compounded by our inability to retrain the regression model in order to account for such drift.

**Table 1:** MAE results from the state-level and ablated models, averaged over all backcast lags and all states, in scenario 1.

| MAE (hospitalization count) |  | MAE (hospitalization rate) |  |
| --- | --- | --- | --- |
| Model | MAE (Standard error) | Model | MAE (Standard error) |
| All-past | 23.74 (1.03) | All-past | 0.44 (0.01) |
| Two-month | 51.79 (2.83) | Two-month | 0.86 (0.02) |
| State-level | 21.18 (0.88) | State-level | 0.41 (0.01) |
| Inpat-only | 22.11 (0.88) | Inpat-only | 0.42 (0.01) |
| Outpat-only | 23.17 (1.14) | Outpat-only | 0.43 (0.01) |

**Table 2:**
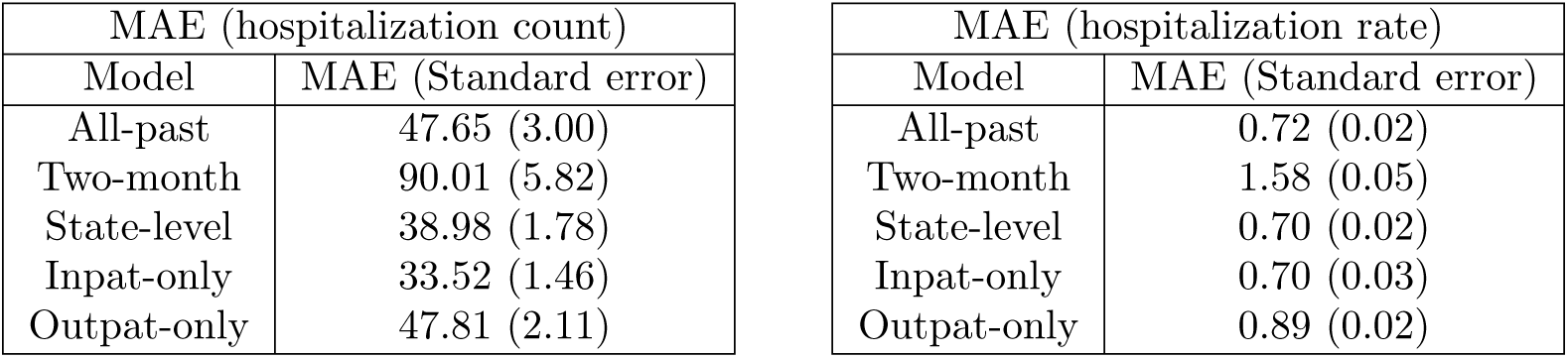
MAE results from the state-level and ablated models, averaged over all backcast lags and all states, in scenario 2.

### 3.6 Prediction interval analysis

We now examine the performance of the quantile tracker, applied to the state-level model in scenario 1. To gain an understanding of how quantile tracking performs against off-the-shelf alternatives, we consider:

- parametric intervals: we apply the standard method for obtaining prediction intervals associated with a well-specified linear regression model with independent Gaussian errors;
- sample quantiles: we obtain intervals by simply computing *q^l,k^* and *q^u,k^* as sample quantiles of all past scores (in lieu of quantile tracking), where the empirical distribution of past scores is weighted with the same decaying weights that are used to train the regression model.

We and evaluate all methods in terms of both coverage of their prediction intervals, and interval score (a metric that combines coverage and sharpness), which for a predicted interval [*l, u*] at the nominal level 1 − *α*, and target value *y*, is defined as

Figure 8 reports interval score and coverage averaged over all dates in scenario 1, at the nominal coverage levels 0.6 and 0.8. (The setup is the same as that in Figure 3, except with either interval score or coverage replacing absolute error.) Quantile tracking clearly dominates the other methods, and it is the only method achieving anywhere close to the nominal coverage level.

**Figure 8:**
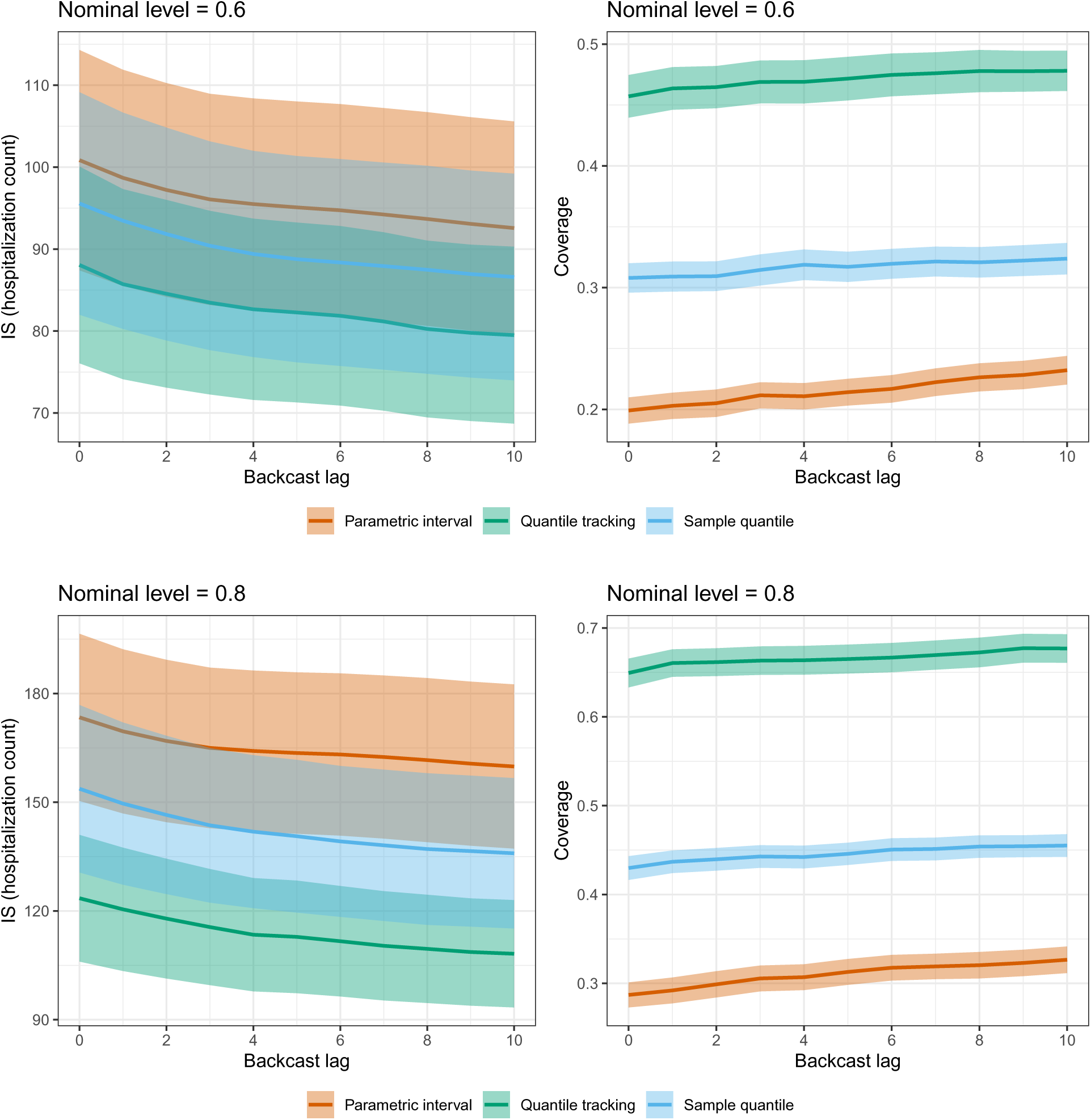
Interval score and coverage as a function of backcast lag for quantile tracking, parametric (linear regression) modeling, and sample quantiles in scenario 1, when the nominal coverage level is 0.6 (first row) and 0.8 (second row). The shaded regions show ±1 standard error bands, over the state values.

Figure 9 visualizes the intervals formed by quantile tracking and the two alternative methods in CA, KY, VT. The nominal coverage level is 0.8. We can see that quantile tracking generally accounts for uncertainty much better than the other methods, avoiding exceedingly narrow intervals.

**Figure 9:**
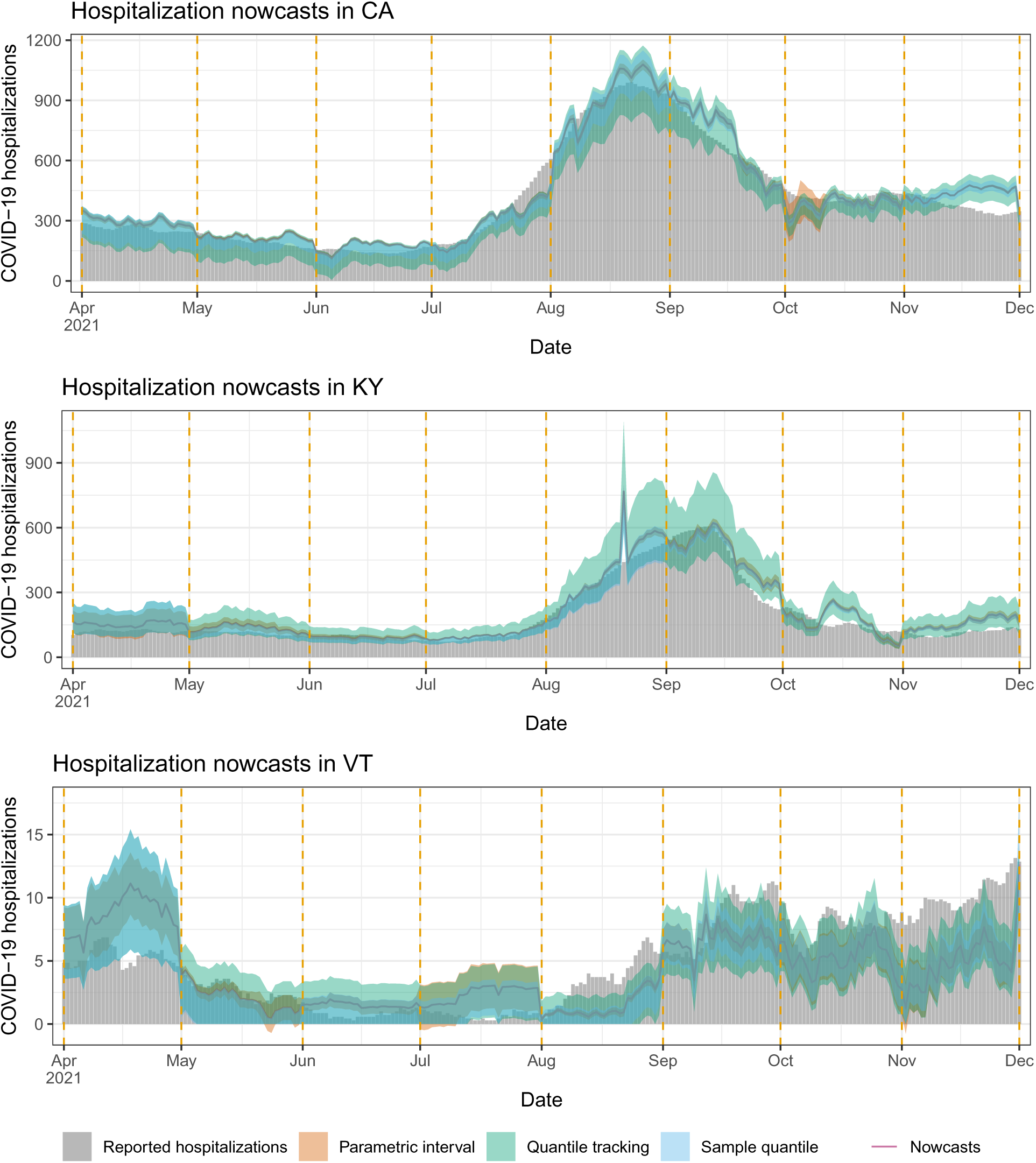
Nowcasts and prediction intervals from quantile tracking, parametric (linear regression) modeling, and sample quantiles in scenario 1, for CA, KY, and VT.

## 4 Discussion

This paper demonstrates that relatively simple regression models and medical insurance claims data can be used to provide accurate real-time estimates of reported COVID-19 hospitalizations, in different hypothetical scenarios in which hospitalization reporting is dramatically reduced in frequency, or shut down entirely. Of course, we are not advocating for nowcasts from claims data to replace traditional public health surveillance. However, leveraging statistical models which use auxiliary data for nowcasting, so that we may then reduce reporting frequency and thus reduce the burden that reporting entails, may be a favorable tradeoff for public health agencies to consider.

We now reflect on some aspects surrounding modeling in this paper. First, the working linear models in the methods section could have been expressed equivalently using an appropriate probabilistic framing. For example, if we ignore versioning for simplicity, we can of course recast the regression in (2) used to make predictions at time *t* as the maximum likelihood estimator in the model:

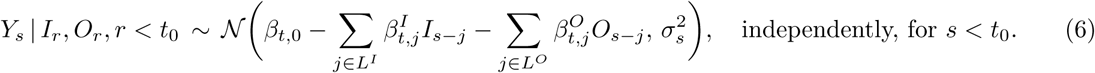

where N (*a, b*^2^) denotes the normal distribution with mean *a* and variance *b*^2^, and where *σ*^2^ = *w_γ_*(*t*_0_ − *s*)^−1^. Specifying distributional assumptions explicitly, as done in (6), becomes particularly important if one relies on the model for problems of inference. Our focus in this paper was primarily on prediction, evaluated via pure prospective predictive accuracy. However, recall, we did touch on inference by investigating prediction intervals, and for this, we found that the parametric prediction intervals generated by the Gaussian linear model (6) performed significantly worse according to coverage or WIS (see Figures 8 and 9) compared to the nonparametric intervals obtained using quantile tracking. For this reason, and because of our focus on prediction more generally, we chose to present our models in the methods section from the perspective of optimization rather than from a probabilistic framing.

The observation weights in (3), used to deal with nonstationarity in the relationship between *Y_s_*and the signals *I_s_*_−*j*_ and *O_s_*_−*j*_ as *s* varies, cause the variance in the equivalent probabilistic model (6) to diverge as *s* moves back into the past. Traditional time series models handle nonstationarity in a different manner. For example, in ARIMA, we would model the differences (of a given order) of the series as a stationary process. This accommodates certain forms of nonstationarity, depending on the order *d* of the difference, i.e., *d* = 1 accommodates a linear drift, *d* = 2 a quadratic drift, and so on. Meanwhile, the model (2) or equivalently (6) allows for a more general form of nonstationarity, albeit one that varies smoothly in time (the exponential training weights can be viewed as a softer version of using a trailing training window in (2), or limiting the number of samples in (6)). The ablation studies, recall, exposed the importance of these observation weights (with CV tuning for the decay parameter). It would be interesting to connect the use of these weights to a more traditional time series method. At present, the precise connection is unclear to us, and this may be a useful direction for future work.

In terms of smoothing, the geo-pooled (4) and mixed models (5) were simple modifications of the basic regression model (2), based on pooling the training data across different locations. The literature on spatial modeling offers various alternatives for richer models which leverage spatial dependence more intelligently (note that pooling does not leverage this dependence at all). Unfortunately, the resolution (and data) we have available in our study is too coarse to support this type of model. Epidemics are driven by much more local dynamics, and at the local level, further issues like political dynamics (Jones and Kiley, 2020) and especially social determinants of health (Khan et al., 2022) likely become important factors as well in terms of modeling and leveraging dependence.

Transitioning to discussing data sources, the medical claims signals used in our work have been available, in real-time, for essentially the entire COVID-19 pandemic. In general, medical insurance claims cover a wide range of health conditions and COVID-19 is not the only worth target for nowcasting systems built on top of claims data. As an example, nowcasts of influenza hospitalizations would also be useful to public health decision-makers, and could be generated via analogous regression models using proxy claims signals. Whether an approach like ours could be used operationally in a future pandemic is a complex issue, which depends on many factors (including data availability). That said, one could argue that methods which tend to be the most reliable in times of emergency are the ones which have been developed and tested ahead of time. This motivates us to continue working on proof-of-concept systems like the one in this paper.

In a different vein, medical insurance claims are certainly not the only relevant auxiliary data stream for tracking COVID-19 hospitalizations, and this analysis may be repeated with any number of other auxiliary signals. In operational systems in public health, robustness is (arguably) more important than average-case performance, and failure examples of the proposed nowcasting methods, as seen in the results section (NY in scenario 1, and KY in scenario 2) would likely be concerning to public health decision-makers. These failure examples were driven by large revisions to the claims signals, occurring at certain points in time. Any model which makes predictions based on a linear combination of a set of features can suffer from erratic behavior if these features are subject to large fluctuations at prediction time. Combining multiple nowcasts built from different auxiliary signals is a way to improve robustness, especially when some of the signals are more stable and less subject to heavy revisions. Signals derived from electronic medical records (EMR), medical device data, and internet search queries all typically have less backfill compared to insurance claims signals.

Model combination methods—which also go by names such as: aggregation, ensembling, or fusion—have been quite successful in influenza nowcasting systems in pre-pandemic years (e.g., Farrow (2016); Jahja et al. (2019)). Ensembles of COVID-19 forecasts have likewise demonstrated considerable robustness compared to the constituent forecasters (e.g., Cramer et al. (2022); Ray et al. (2023)). An important direction for future work is to incorporate similar ideas into the nowcasting settings considered in this paper, in an effort to move toward more reliable and robust systems.

## Data Availability

All data and code produced are available online at https://github.com/cmu-delphi/hhs-nowcasting

## Acknowledgements

We would like to thank members of the Delphi research group for valuable feedback, and Change Healthcare and Optum/United Health Group for their invaluable data partnership and their collaboration. This work was supported by Centers for Disease Control and Prevention (CDC) grant no. 75D30123C15907.

## A More details on claims signals

### A.1 Coverage of claims signals

We investigate the coverage of our claims data by comparing the counts of hospitalizations from inpatient claims to the HHS reported counts, over our entire analysis period (November 1, 2020 to August 31, 2023). Figure 10 plots the sum of COVID-19 hospitalizations from these sources, broken down to individual states. Figure 11 plots the percentage of COVID-19 hospitalizations recorded by inpatient claims over the same time period.

**Figure 10:**
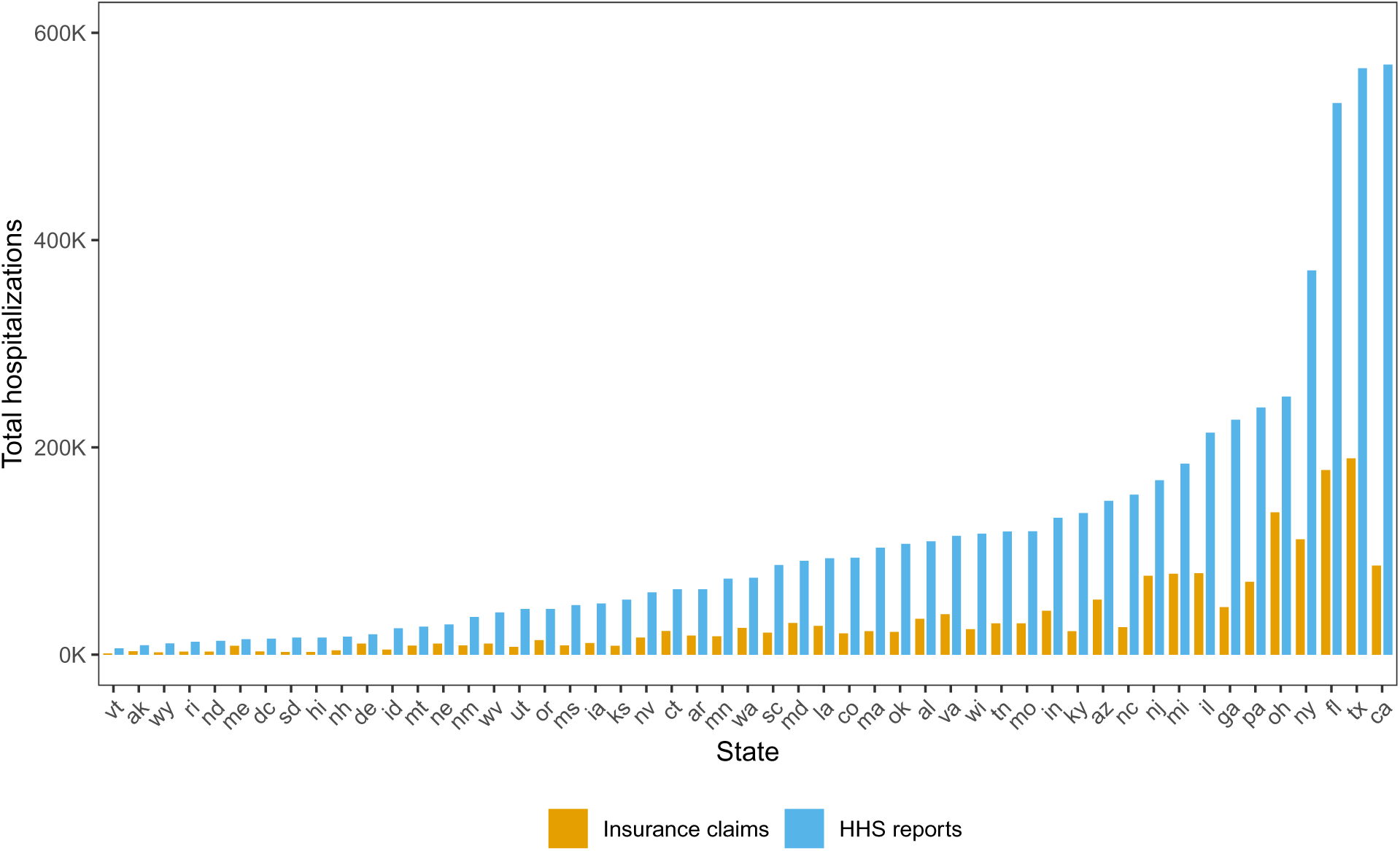
Total number of COVID-19 hospitalizations from official reports and inpatient claims.

**Figure 11:**
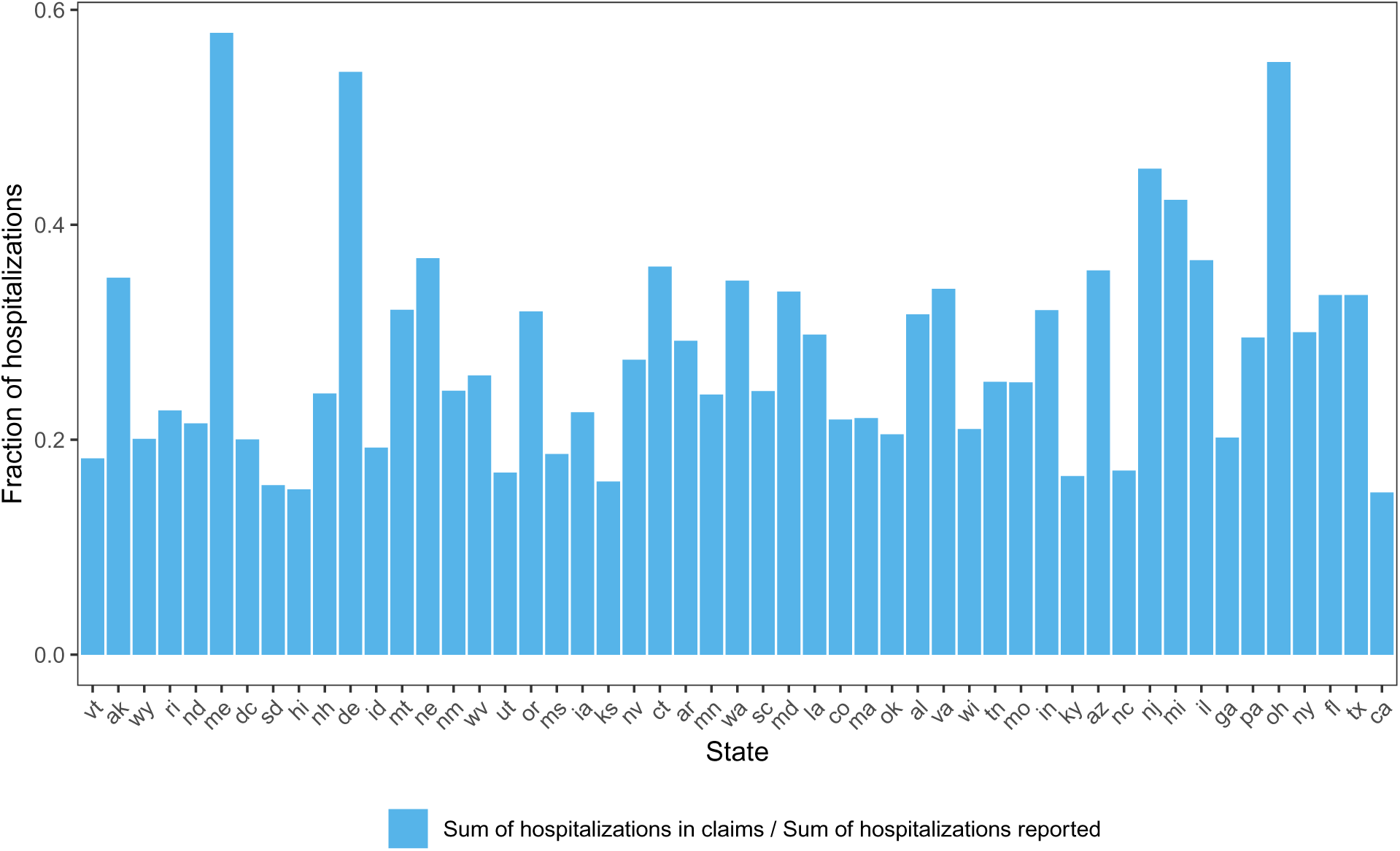
Fraction of reported hospitalizations captured by inpatient claims.

### A.2 ICD-10 codes for claims signals

For the outpatient signal, any claim that has a primary ICD-10 code of U07.1, B97.21, or B97.29 is counted as confirmed COVID. For the inpatient signal, any claim with a primary ICD-10 code of U07.1, U07.2, B97.29, J12.81, Z03.818, B34.2, or J12.89 is counted as COVID-associated.

These definitions are consistent with the definitions of analogous outpatient and inpatient signals in the Delphi Epidata API. However, the signals that we use in our analysis differ from those in the Delphi Epidata API, at the time that this paper was written, in a way that relates to smoothing: the signals in the API are smoothed with a more sophisticated smoothing approach (which also explicitly adjusts for weekday/weekend differences), whereas the signals in this paper are smoothed via 7-day pooling (which implicitly accounts for weekday/weekend differences).

## B Scenario 1: further investigation for NY

We present further analysis as to why state-level model for NY produces negative nowcasts over a portion of the month of June 2021, in scenario 1 (recall Figure 5 in the main paper). Figure 12 displays two versions of the outpatient signal over the month of May 2021, corresponding to issue dates of June 7 and June 8. We can see that a systematic upward revision of all signal values occurs on June 8. The nowcasting model, fit by training on data through the end of May (which sees the revision on June 8), places a sizeable negative weight on the largest lag of the outpatient feature; once the upward revision occurs on June 8, the nowcasts on that and subsequent days display a significant downward trend. This is confirmed by Figure 13, where we decompose the nowcasts made in June into the contributions from each feature (simply the coefficient times the feature value). After June 7, we see that out_20, the largest lag of the outpatient figure, is responsible for driving the downward trend.

**Figure 12:**
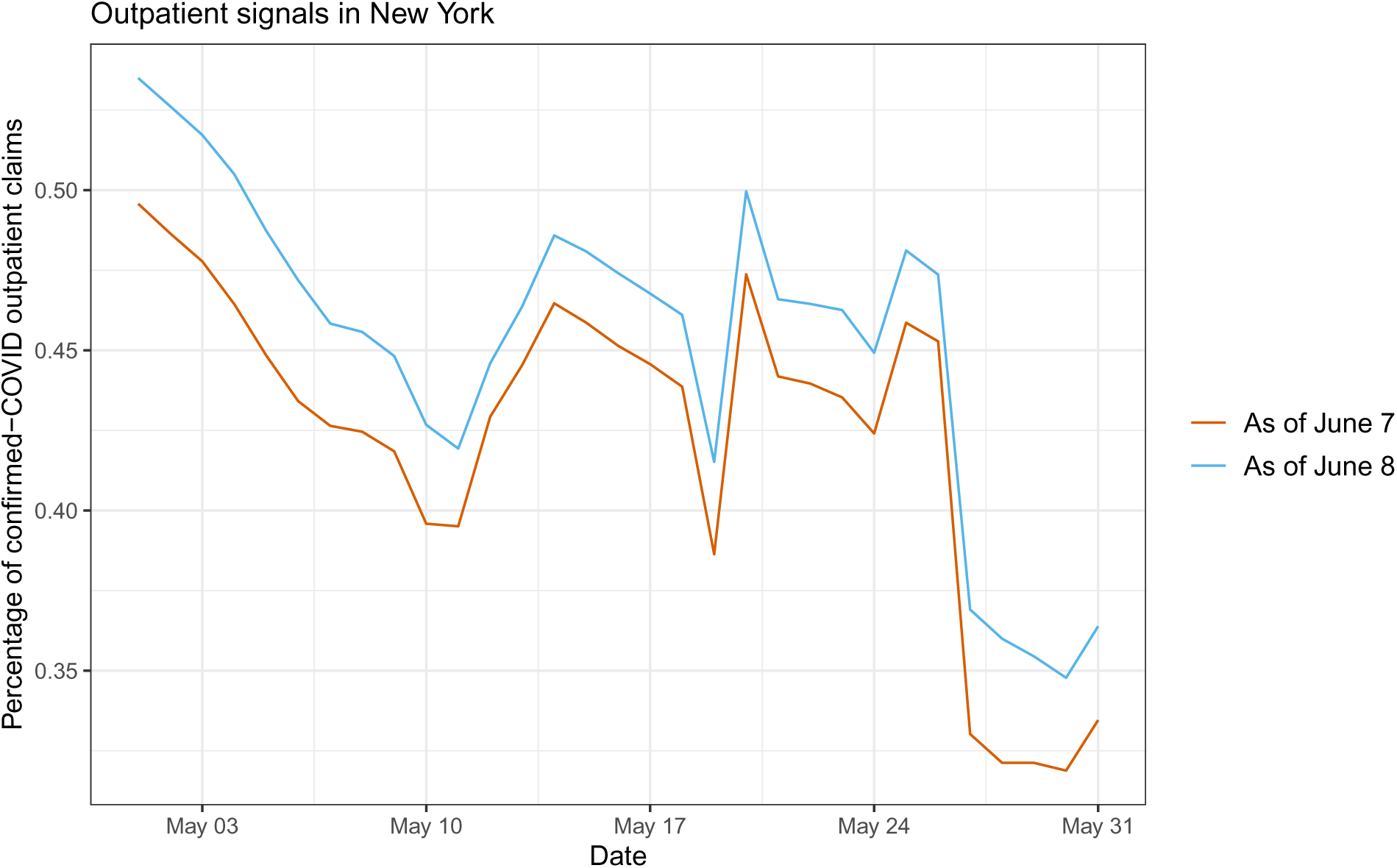
Two versions of the outpatient signal during May 2021, as of June 7 and June 8.

**Figure 13:**
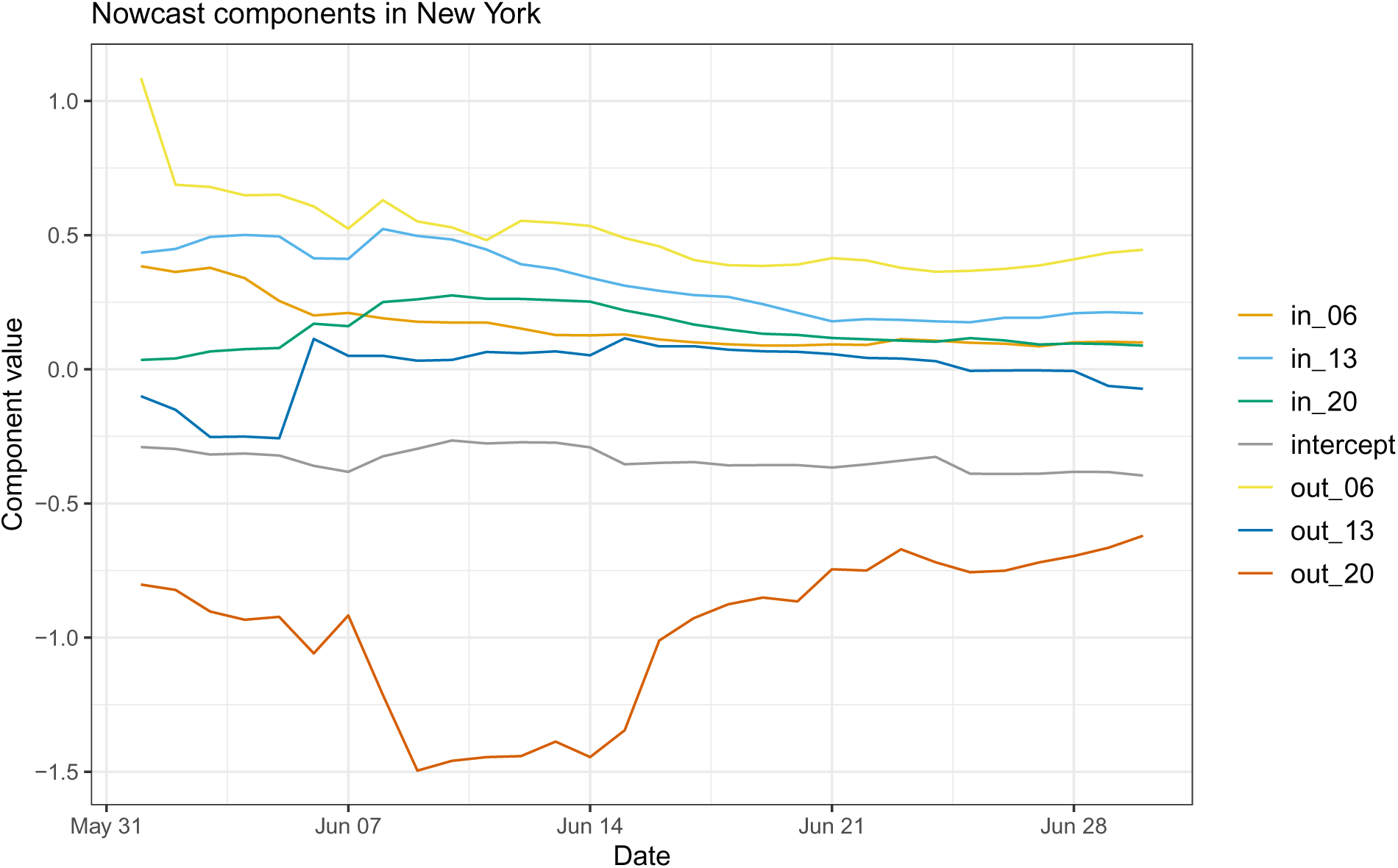
Contributions of each feature to the nowcasts during June 2021.

## C Scenario 1: full set of backcasts

Figures 14–30 display backcasts at lag 0 (i.e., nowcasts), 5, and 10, for all 50 US states and DC, made by the mixed model in scenario 1, the monthly-update period. The format follows Figure 4 in the main paper.

**Figure 14:**
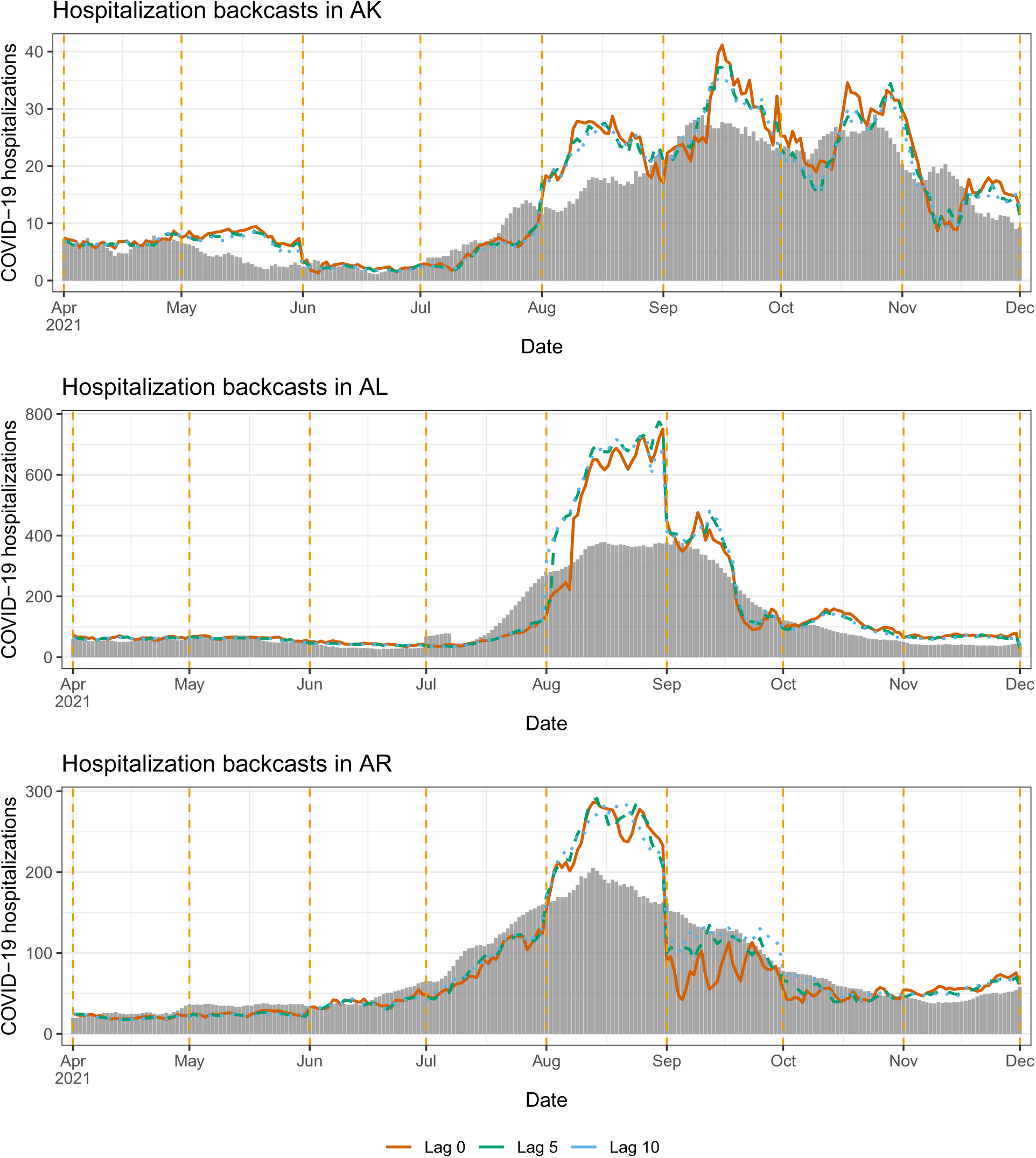
Backcasts from the mixed model in scenario 1, for AL, AK, AZ.

**Figure 15:**
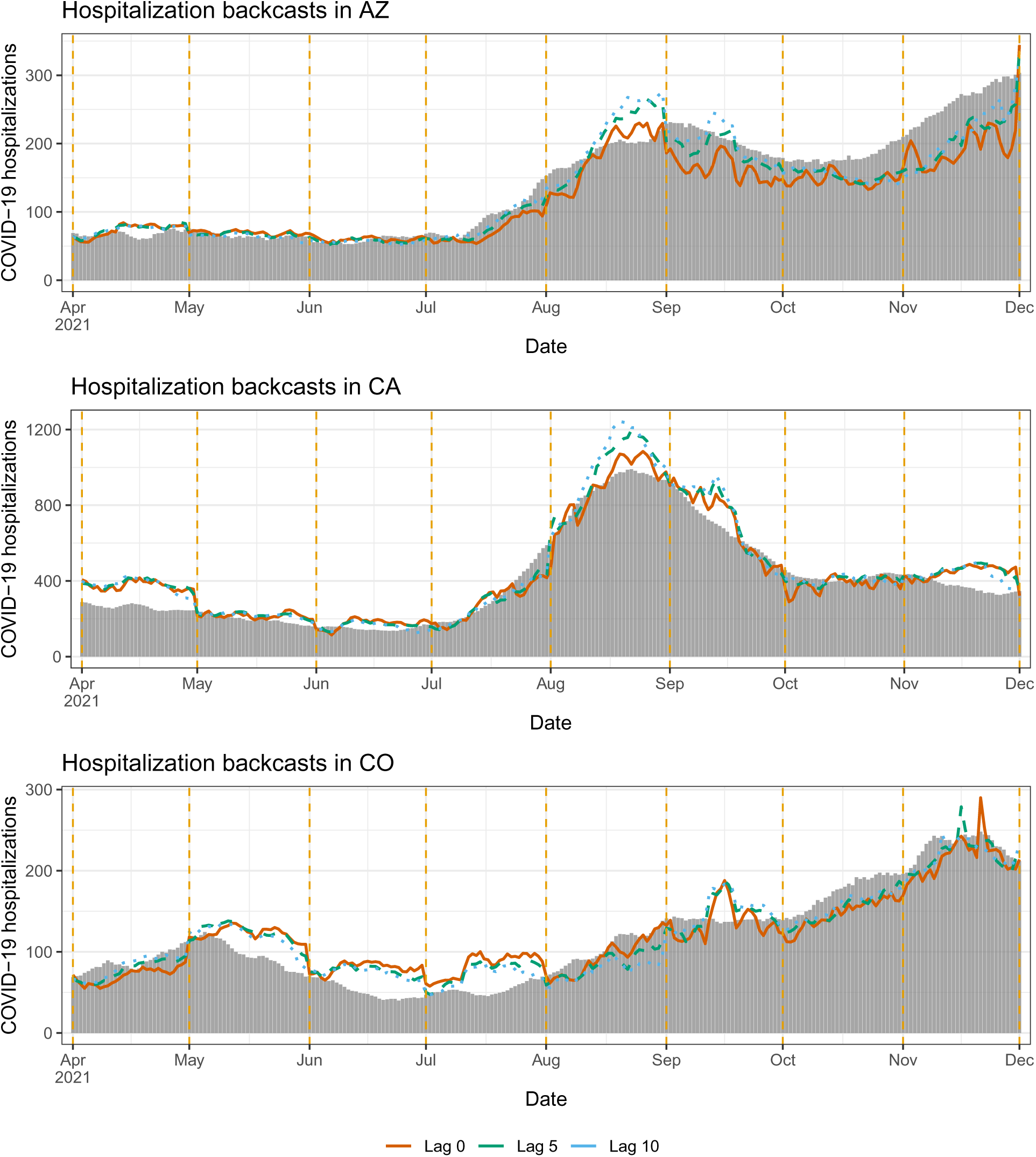
Backcasts from the mixed model in scenario 1, for AR, CA, CO.

**Figure 16:**
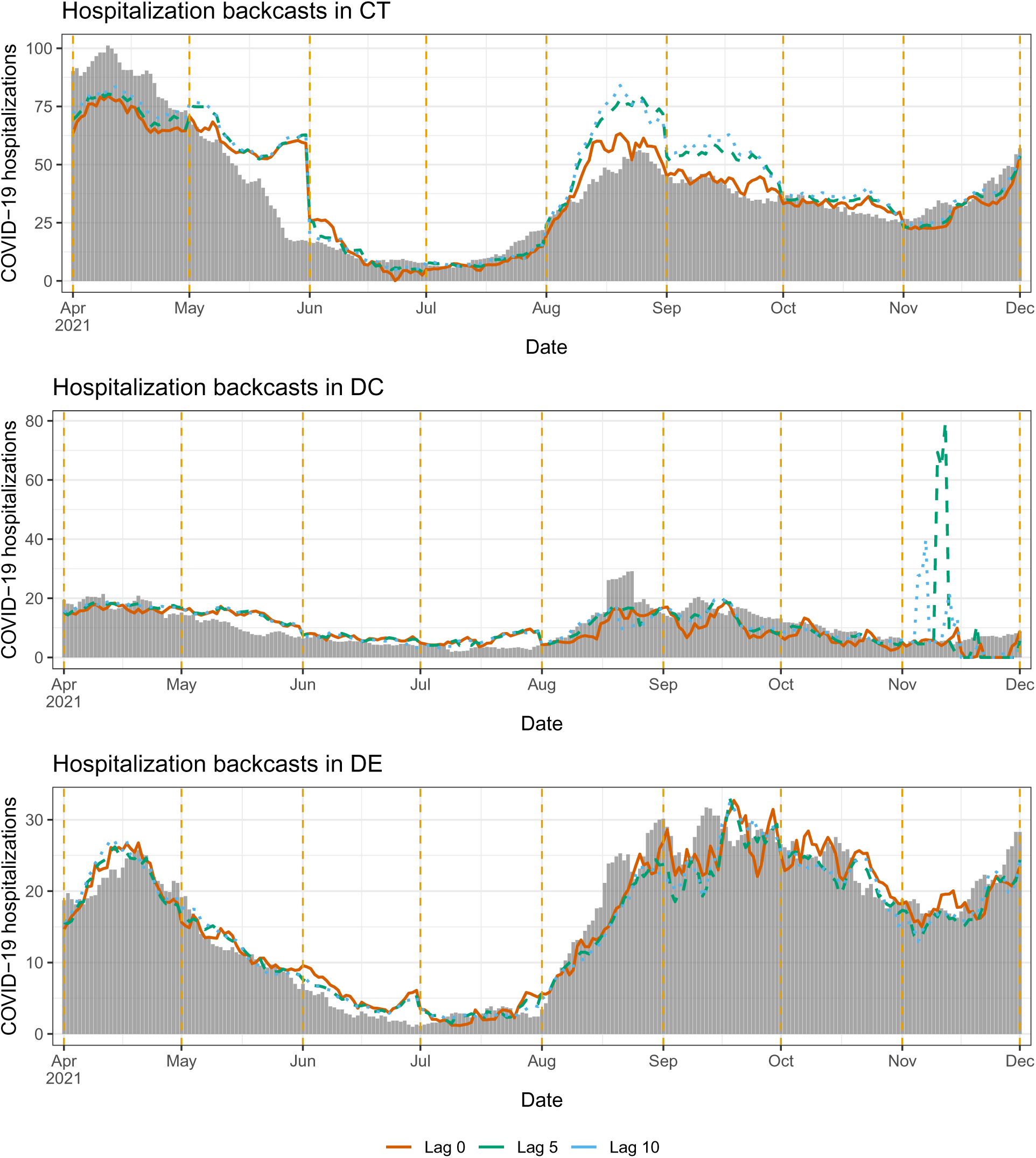
Backcasts from the mixed model in scenario 1, for CT, DC, DE.

**Figure 17:**
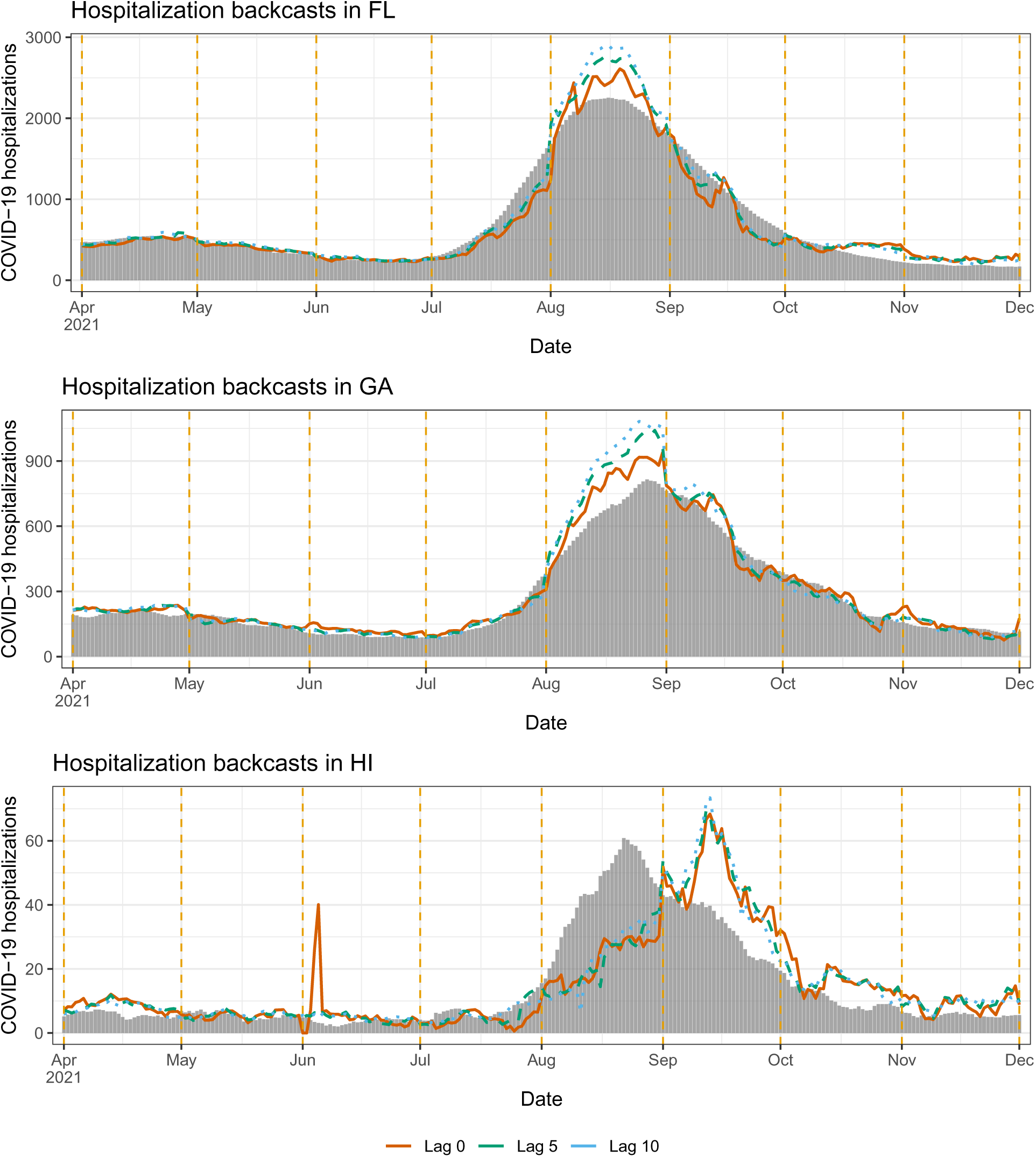
Backcasts from the mixed model in scenario 1, for FL, GA, HI.

**Figure 18:**
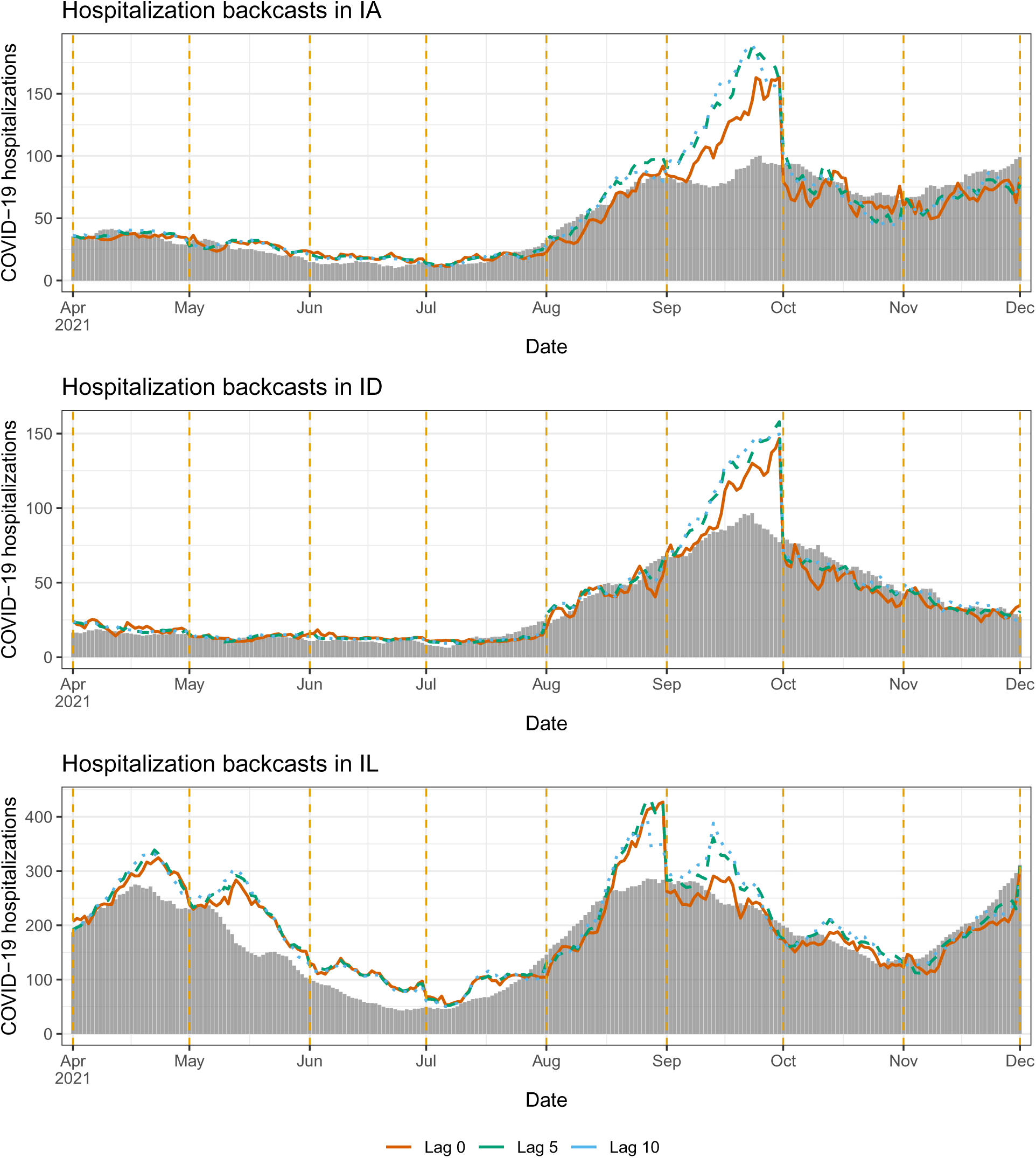
Backcasts from the mixed model in scenario 1, for IA, ID, IL.

**Figure 19:**
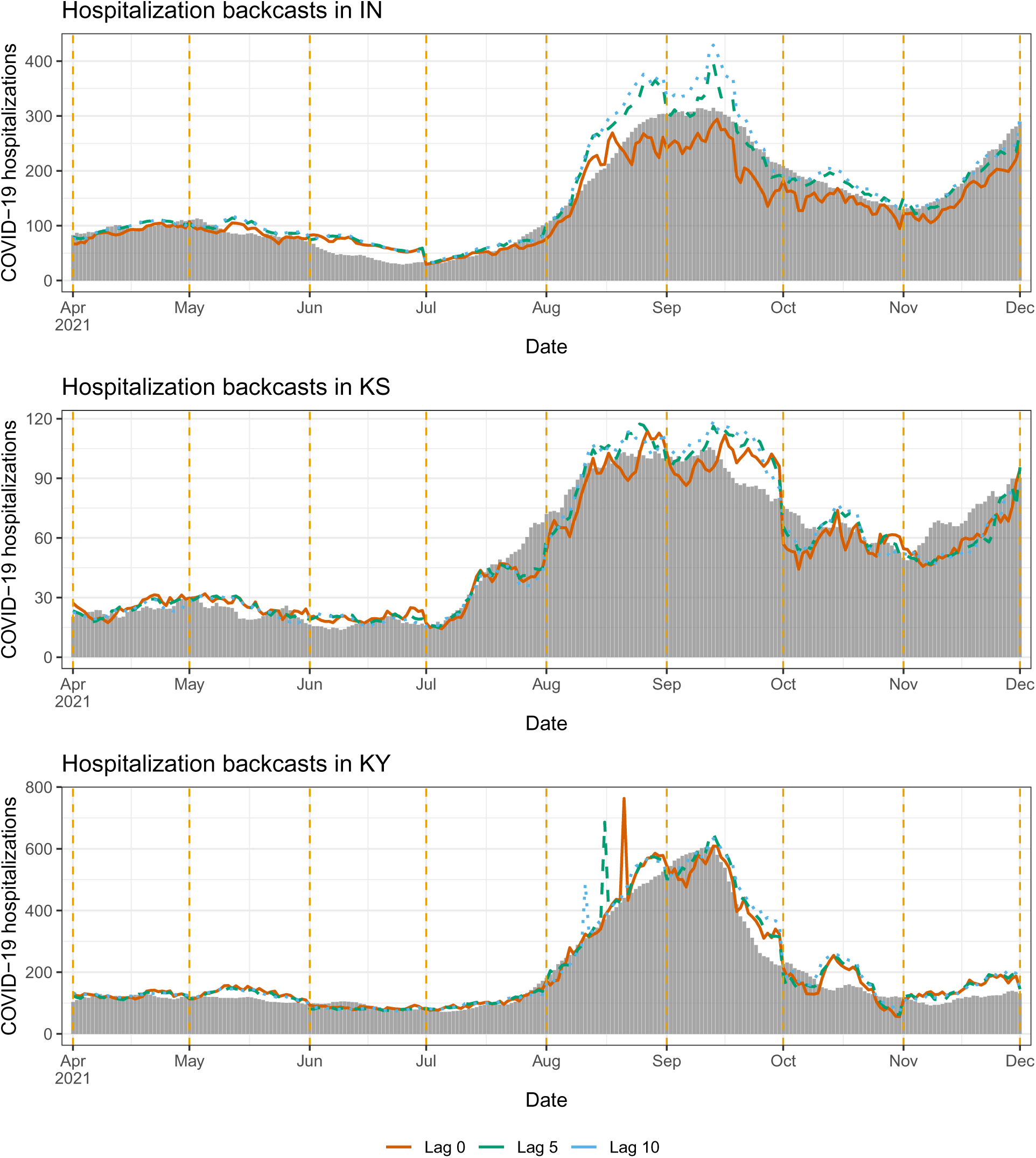
Backcasts from the mixed model in scenario 1, for IN, KS, KY.

**Figure 20:**
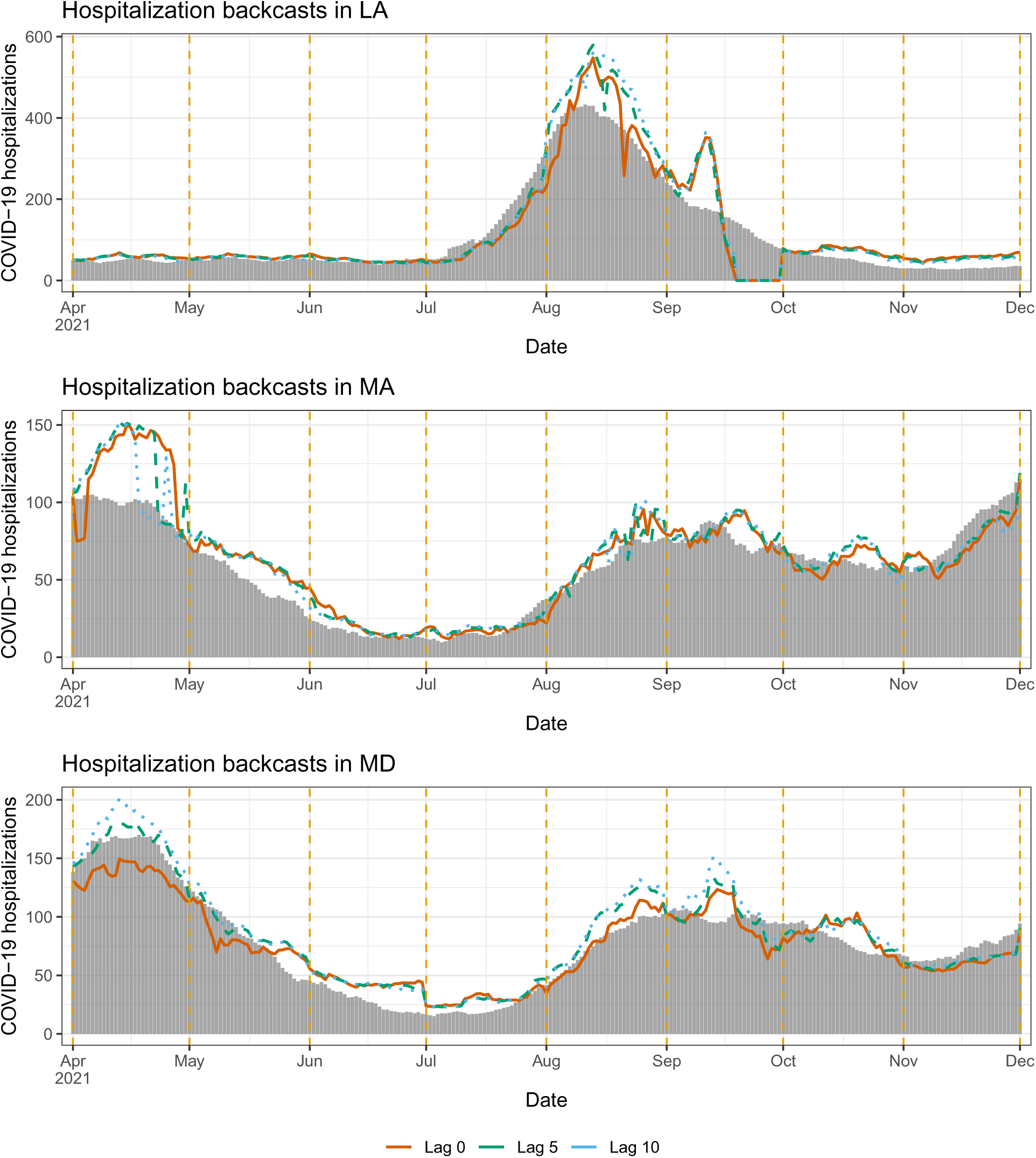
Backcasts from the mixed model in scenario 1, for LA, MA, MD.

**Figure 21:**
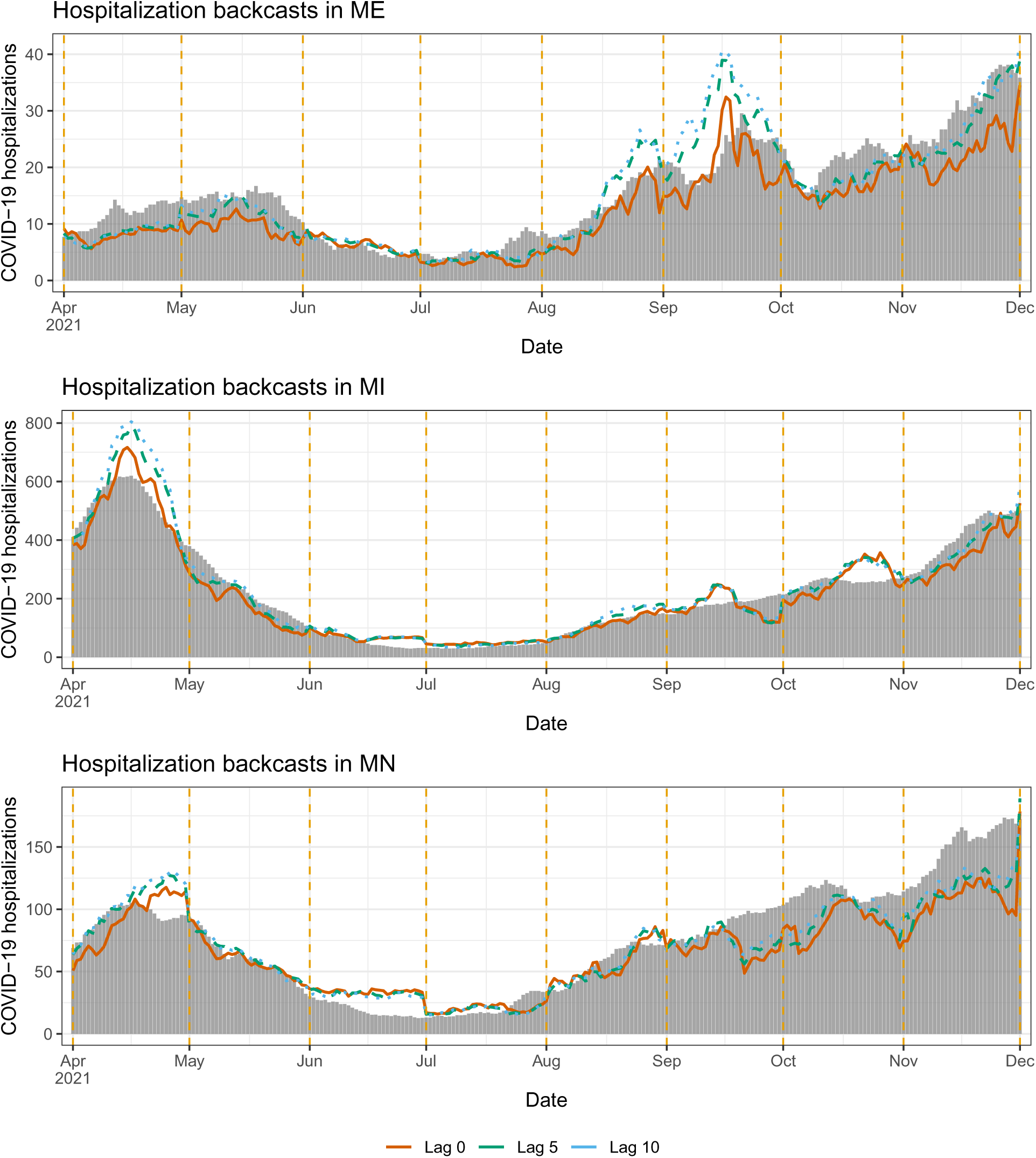
Backcasts from the mixed model in scenario 1, for ME, MI, MN.

**Figure 22:**
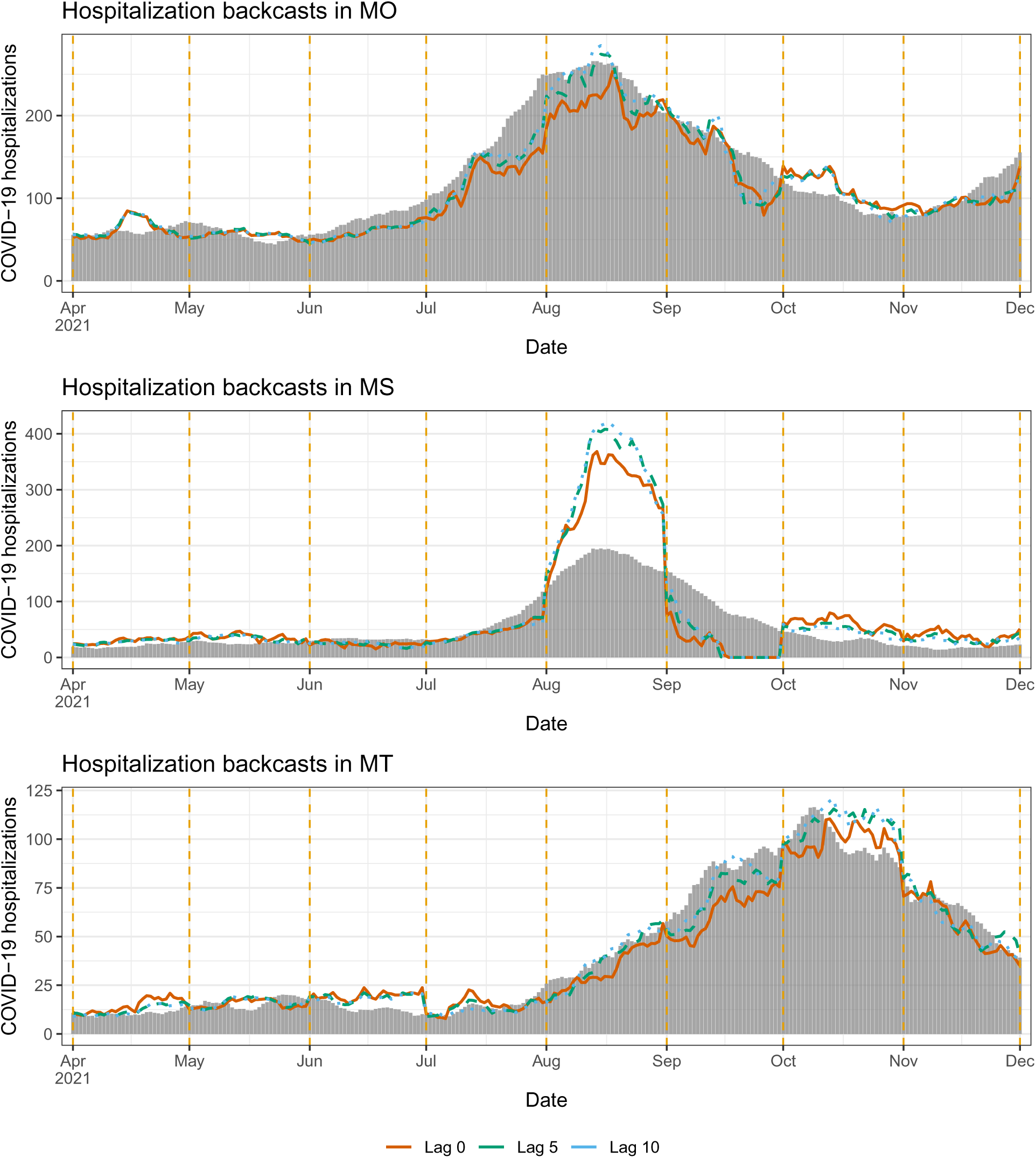
Backcasts from the mixed model in scenario 1, for MO, MS, MT.

**Figure 23:**
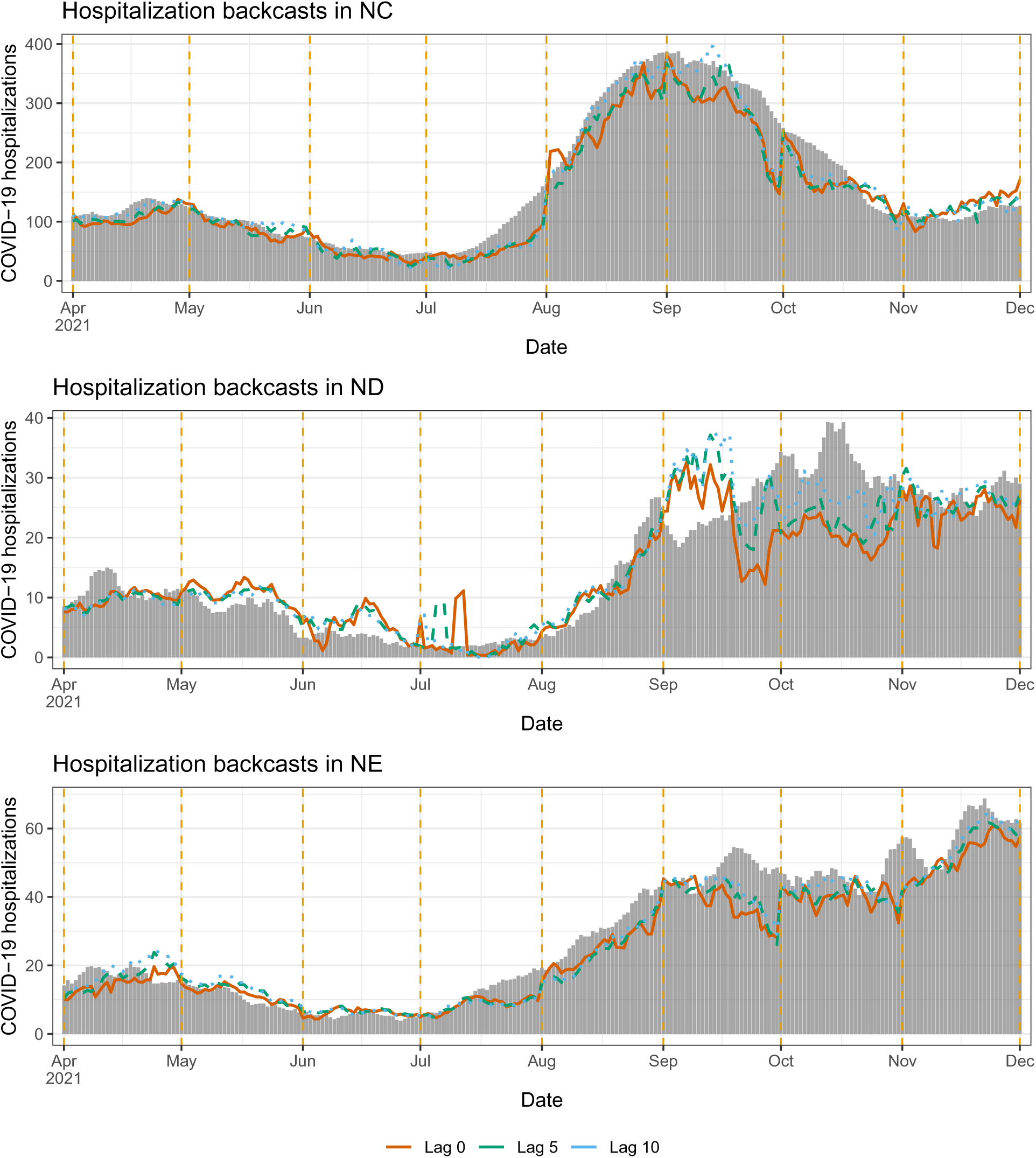
Backcasts from the mixed model in scenario 1, for NC, ND, NE.

**Figure 24:**
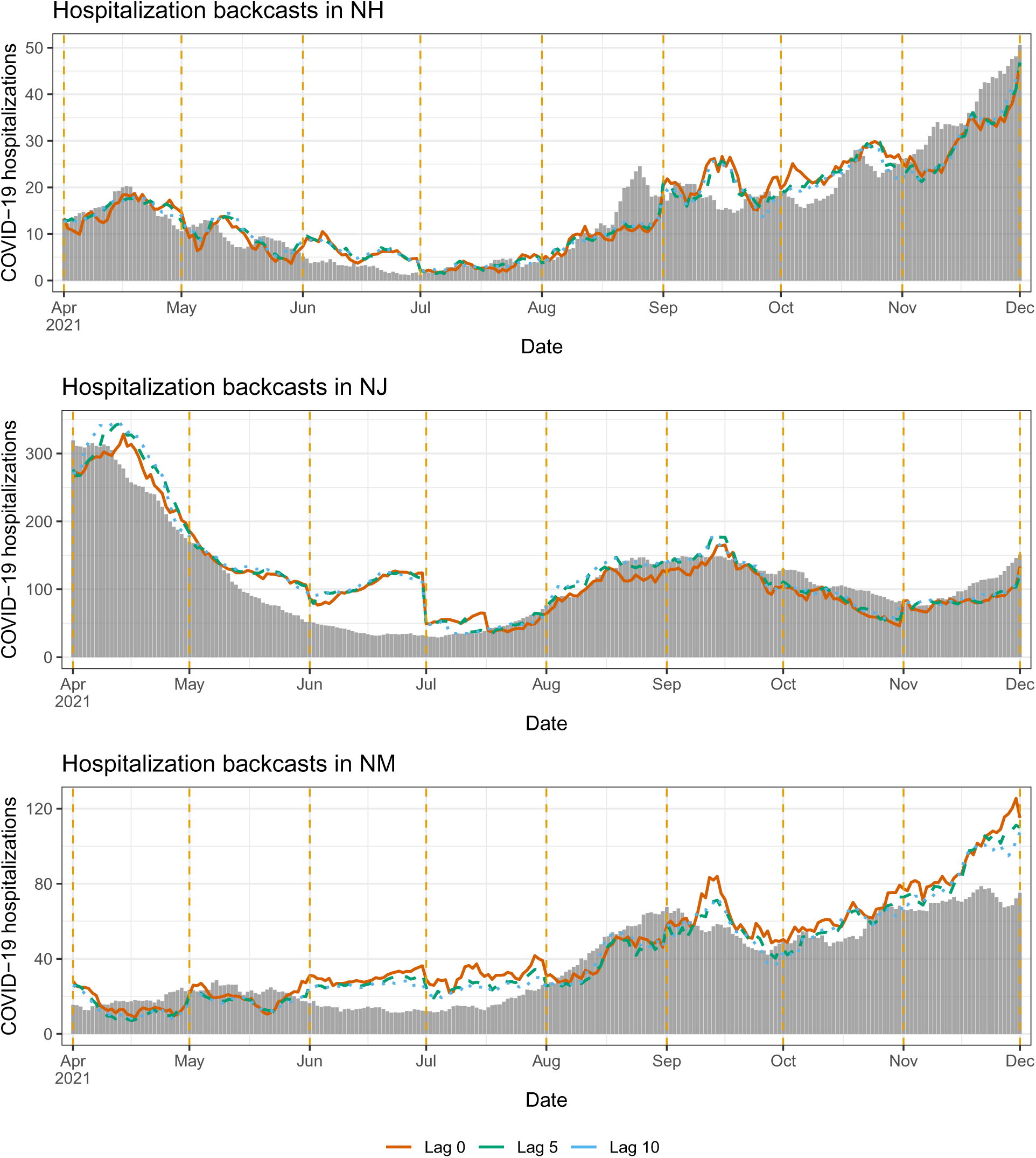
Backcasts from the mixed model in scenario 1, for NH, NJ, NM.

**Figure 25:**
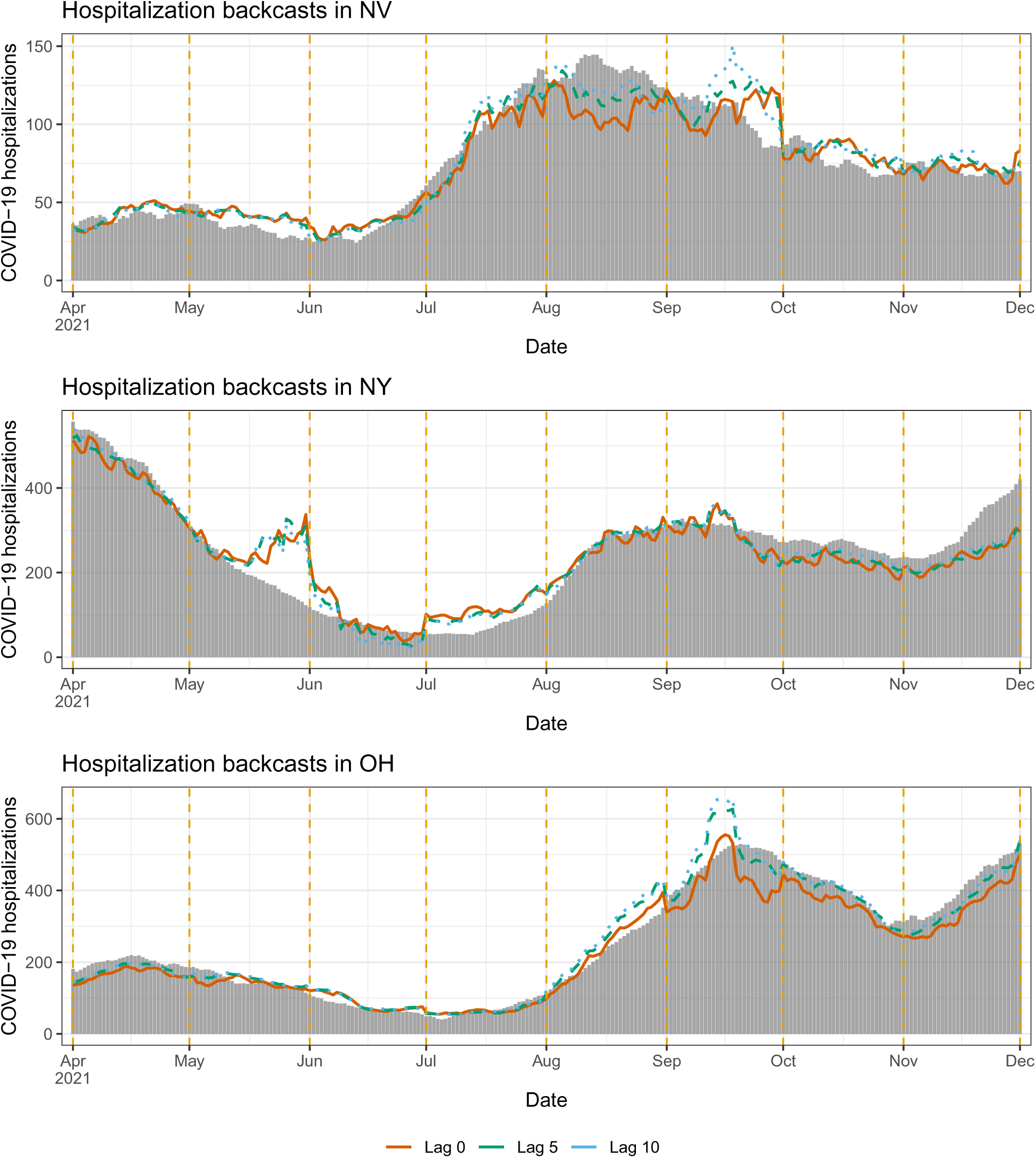
Backcasts from the mixed model in scenario 1, for NV, NY, OH.

**Figure 26:**
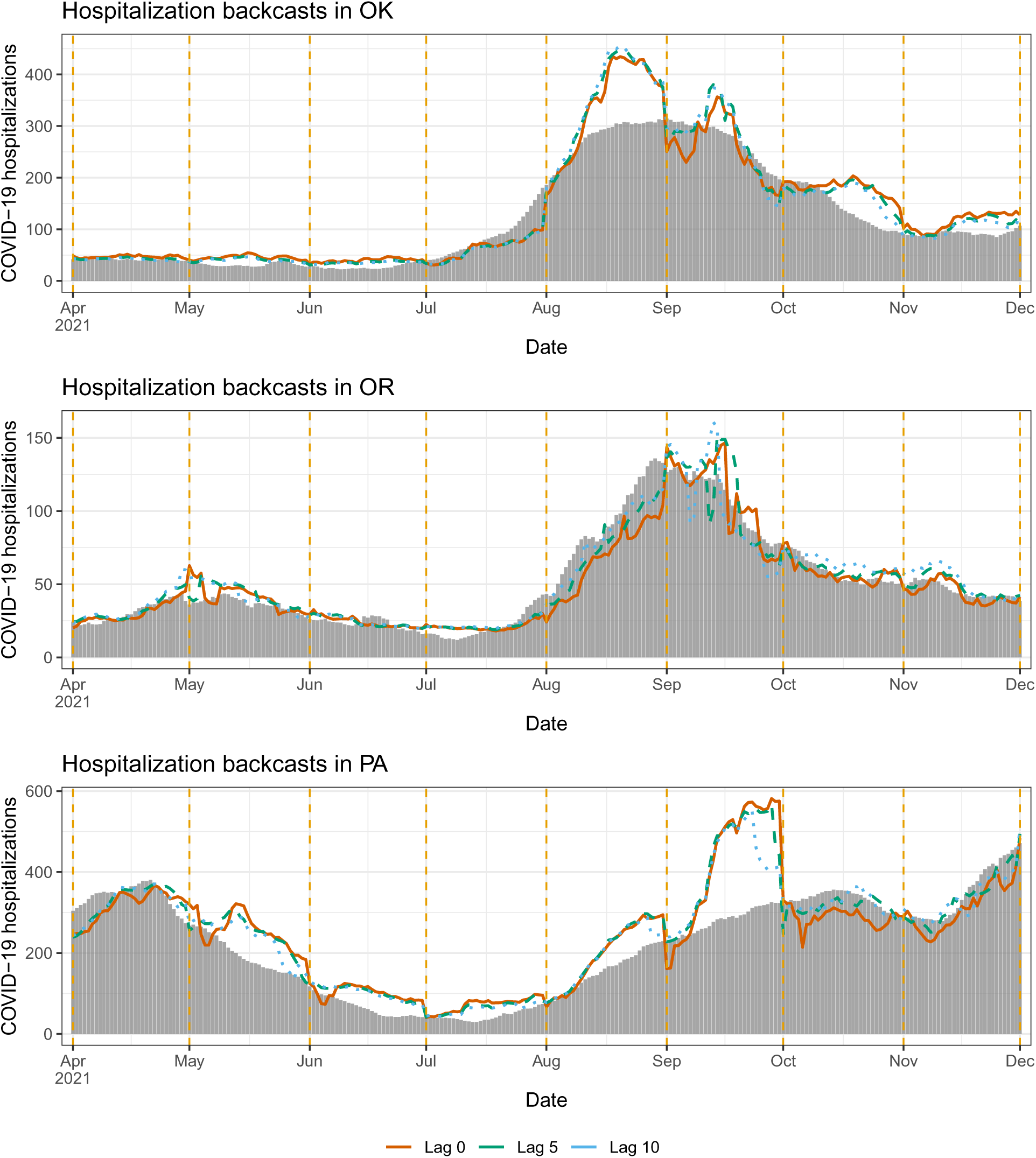
Backcasts from the mixed model in scenario 1, for OK, OR, PA.

**Figure 27:**
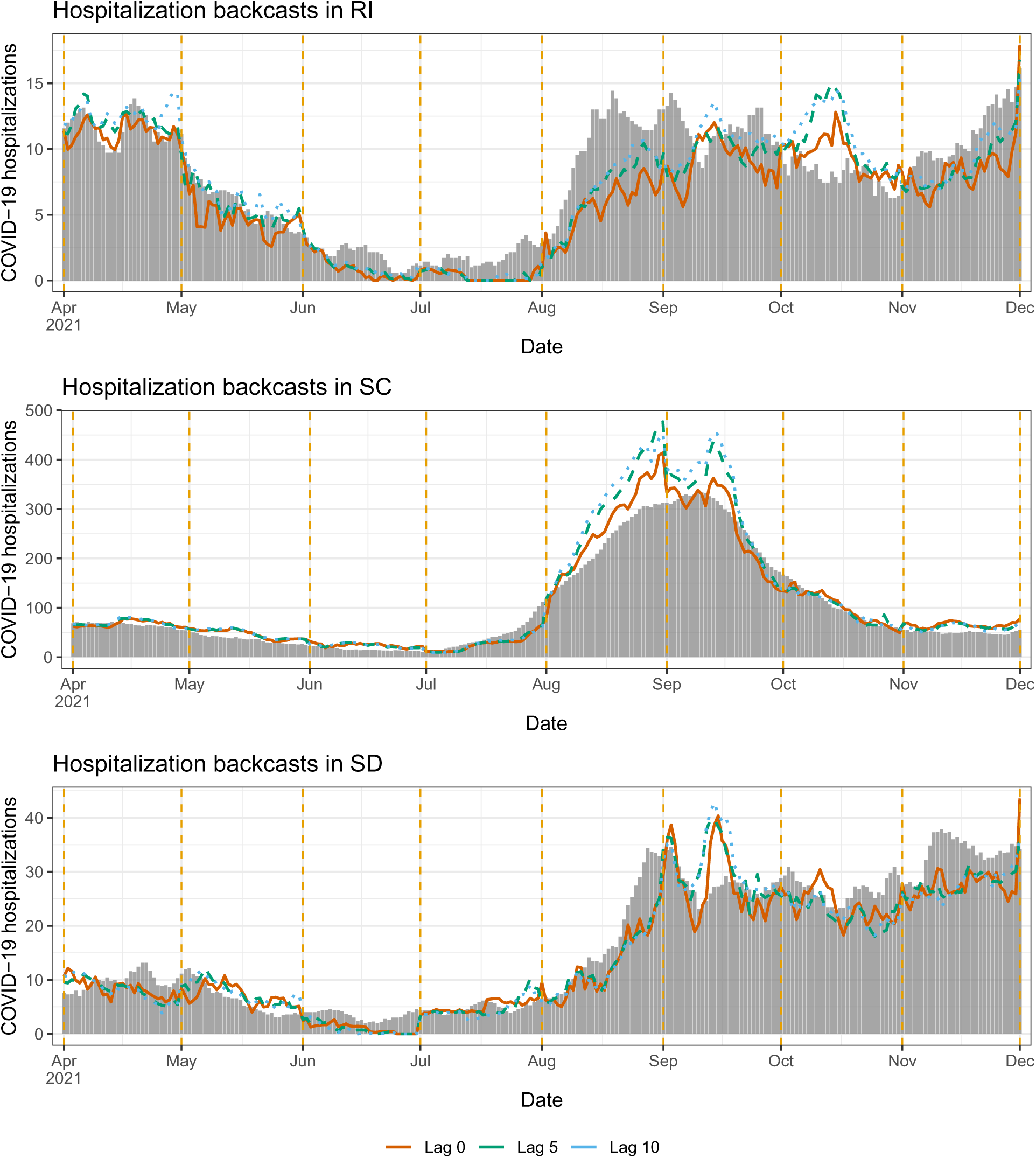
Backcasts from the mixed model in scenario 1, for RI, SC, SD.

**Figure 28:**
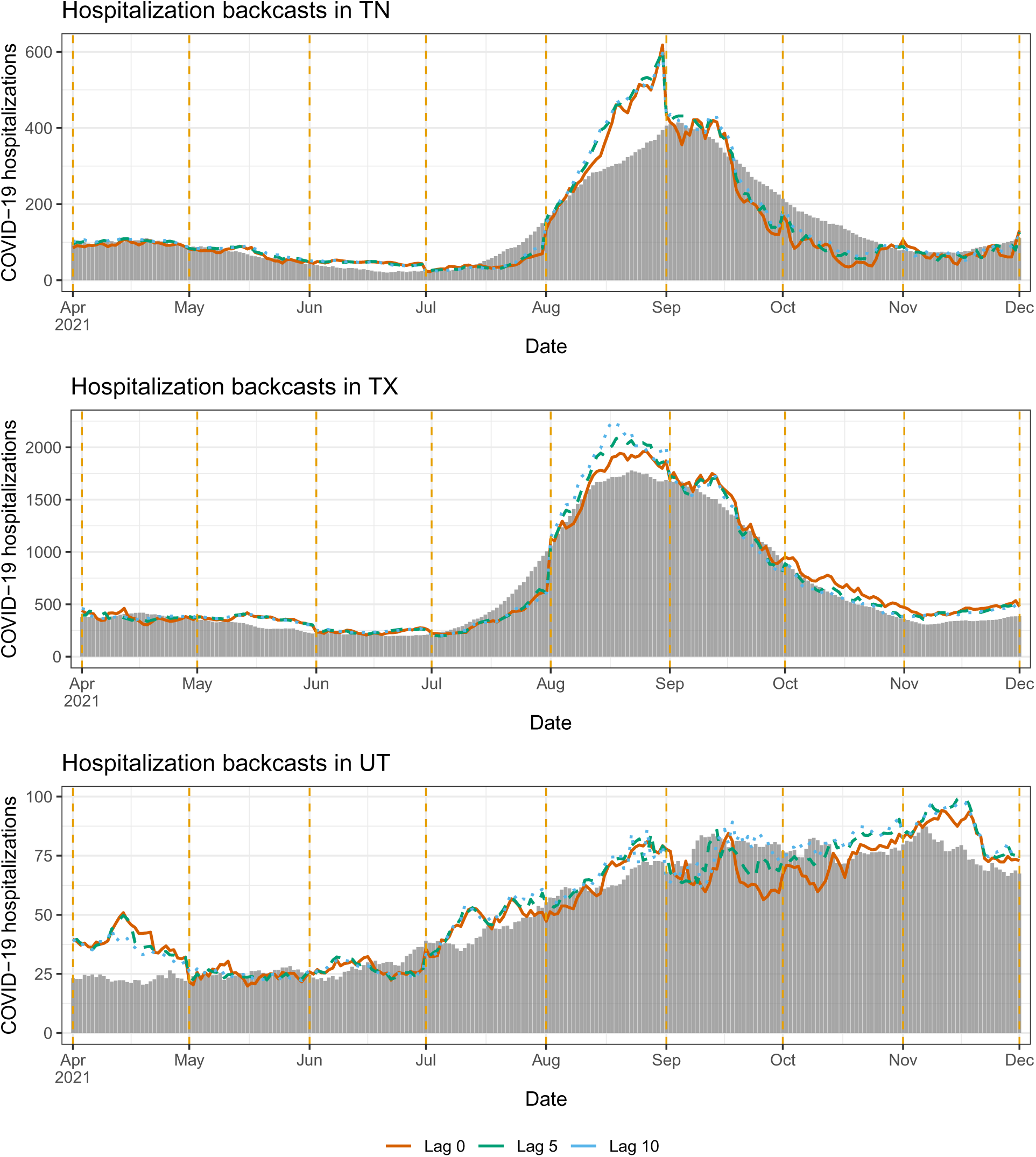
Backcasts from the mixed model in scenario 1, for TN, TX, UT.

**Figure 29:**
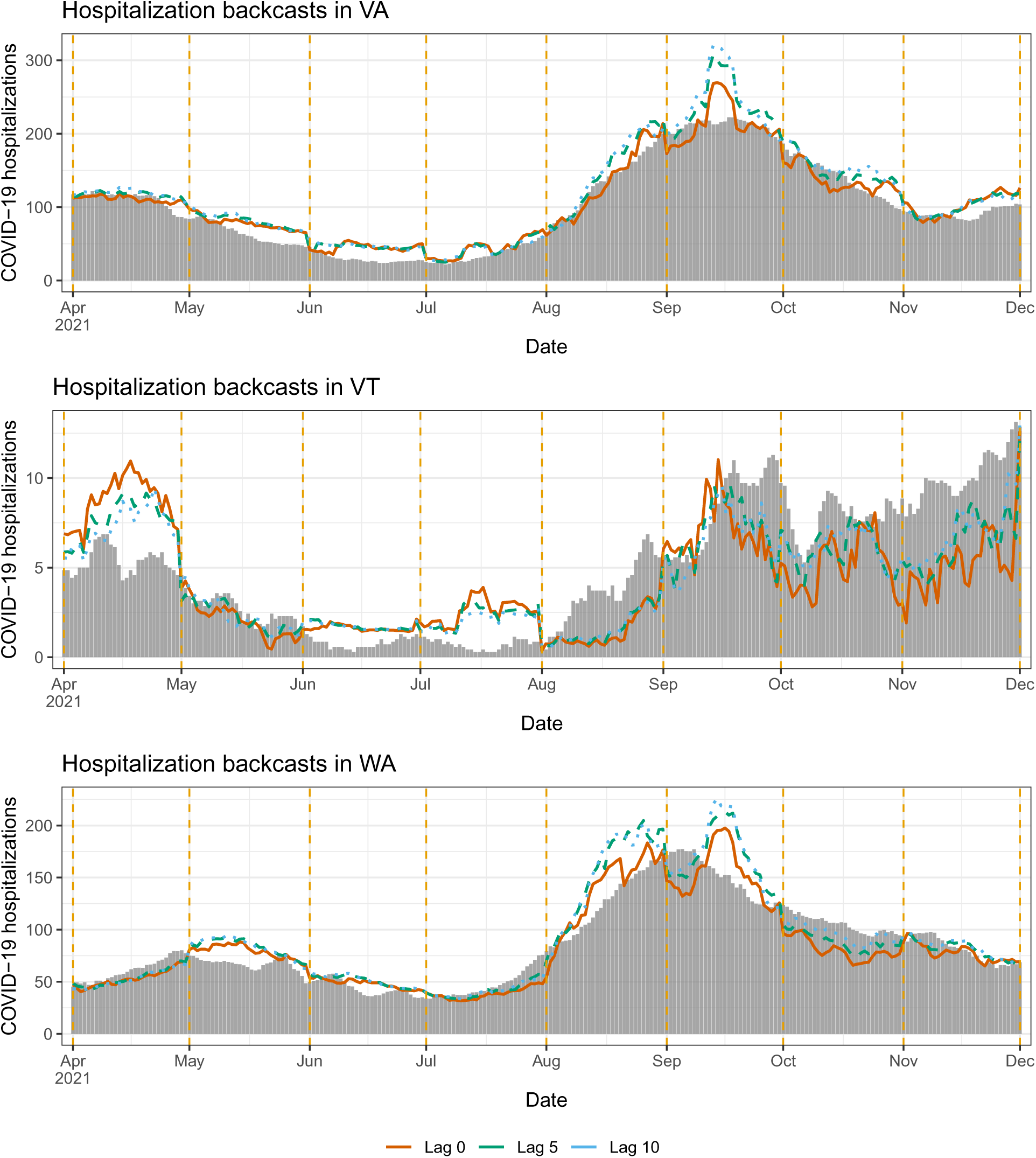
Backcasts from the mixed model in scenario 1, for VA, VT, WA.

**Figure 30:**
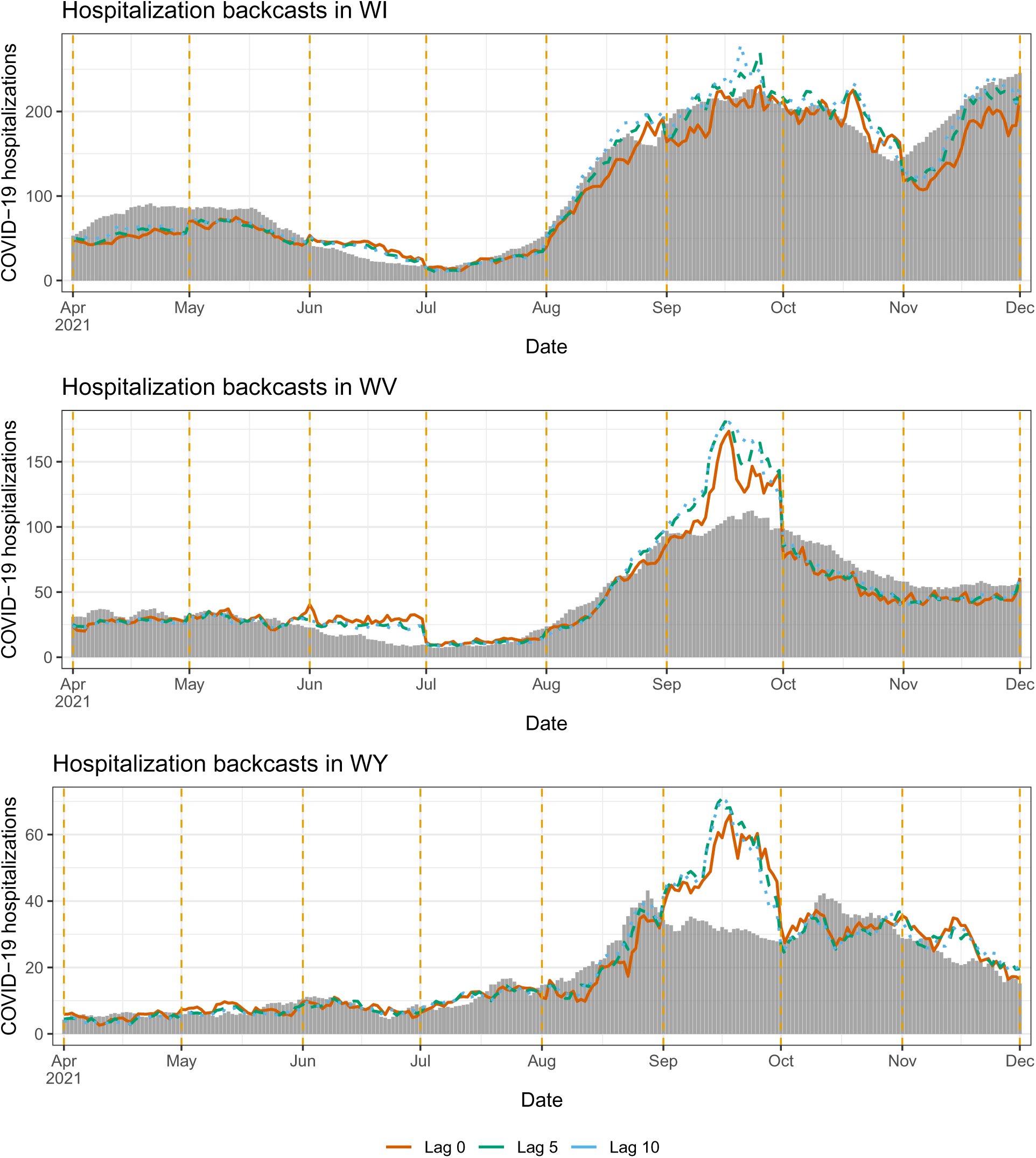
Backcasts from the mixed model in scenario 1, for WI, WV, WY.

## D Scenario 2: full set of backcasts

Figures 31–47 display backcasts at lag 0 (i.e., nowcasts), 5, and 10, for all 50 US states and DC, made by the mixed model in scenario 2, the no-update period. The format follows Figure 7 in the main paper.

**Figure 31:**
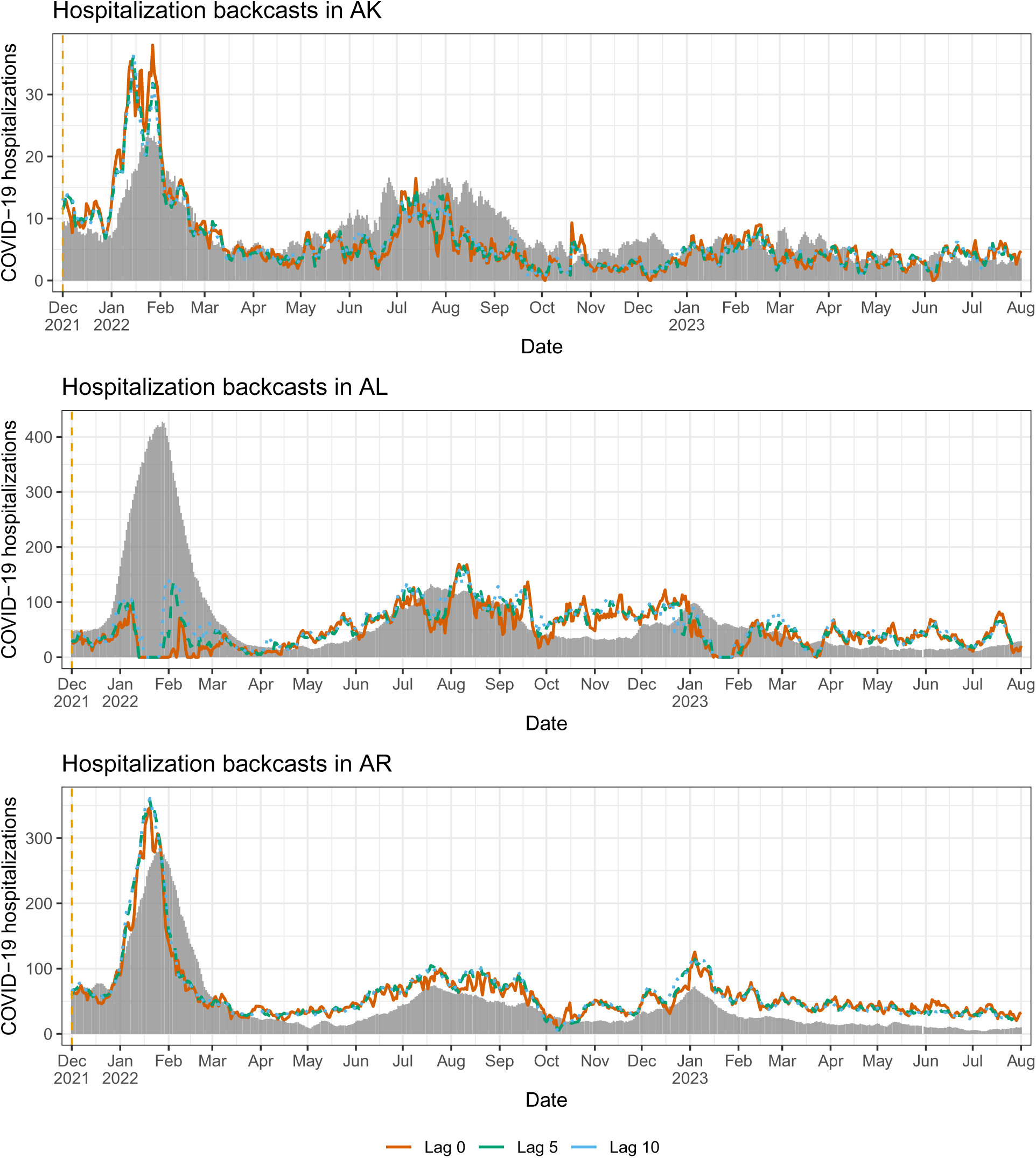
Backcasts from the mixed model in scenario 2, for AL, AK, AZ.

**Figure 32:**
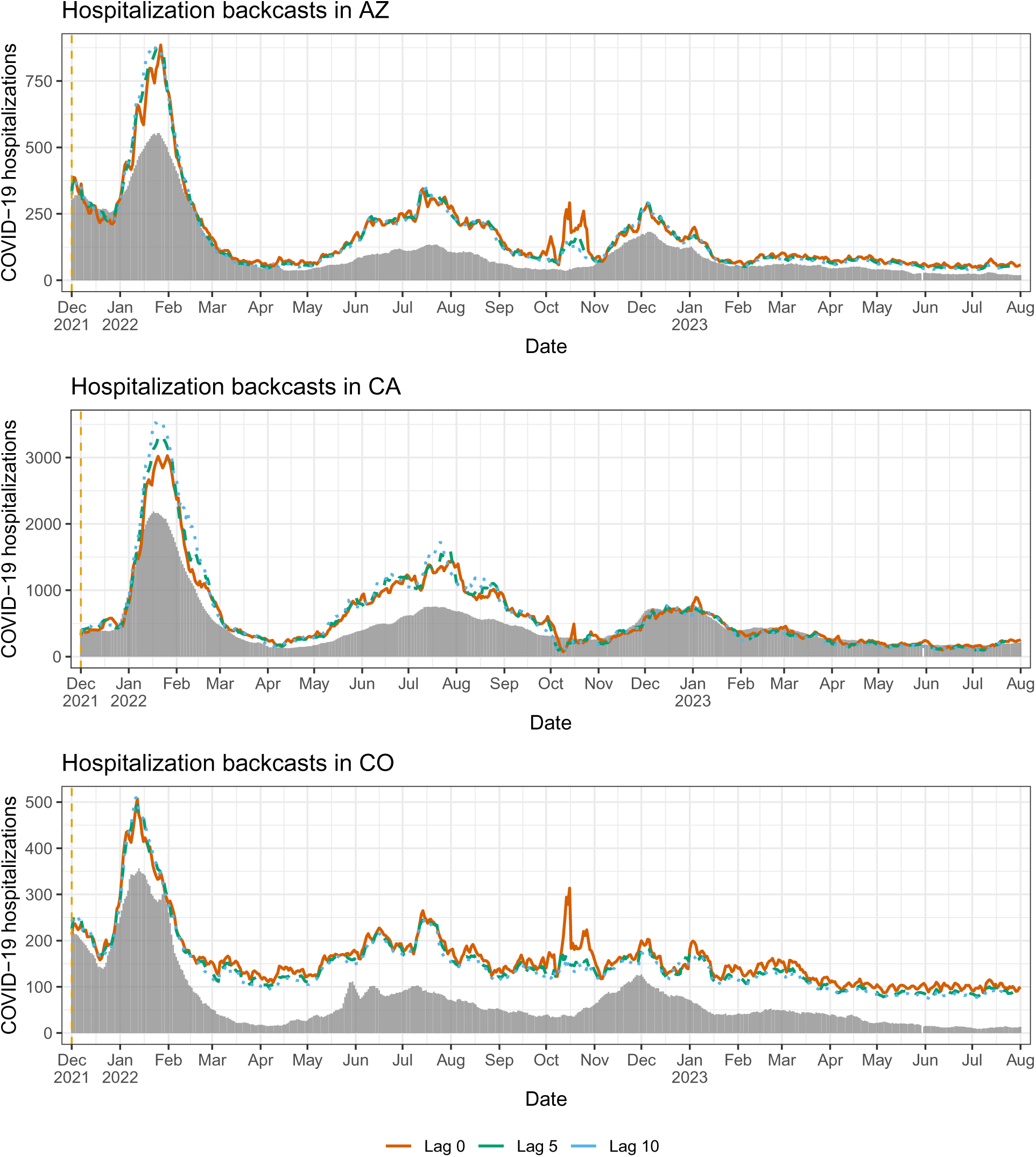
Backcasts from the mixed model in scenario 2, for AR, CA, CO.

**Figure 33:**
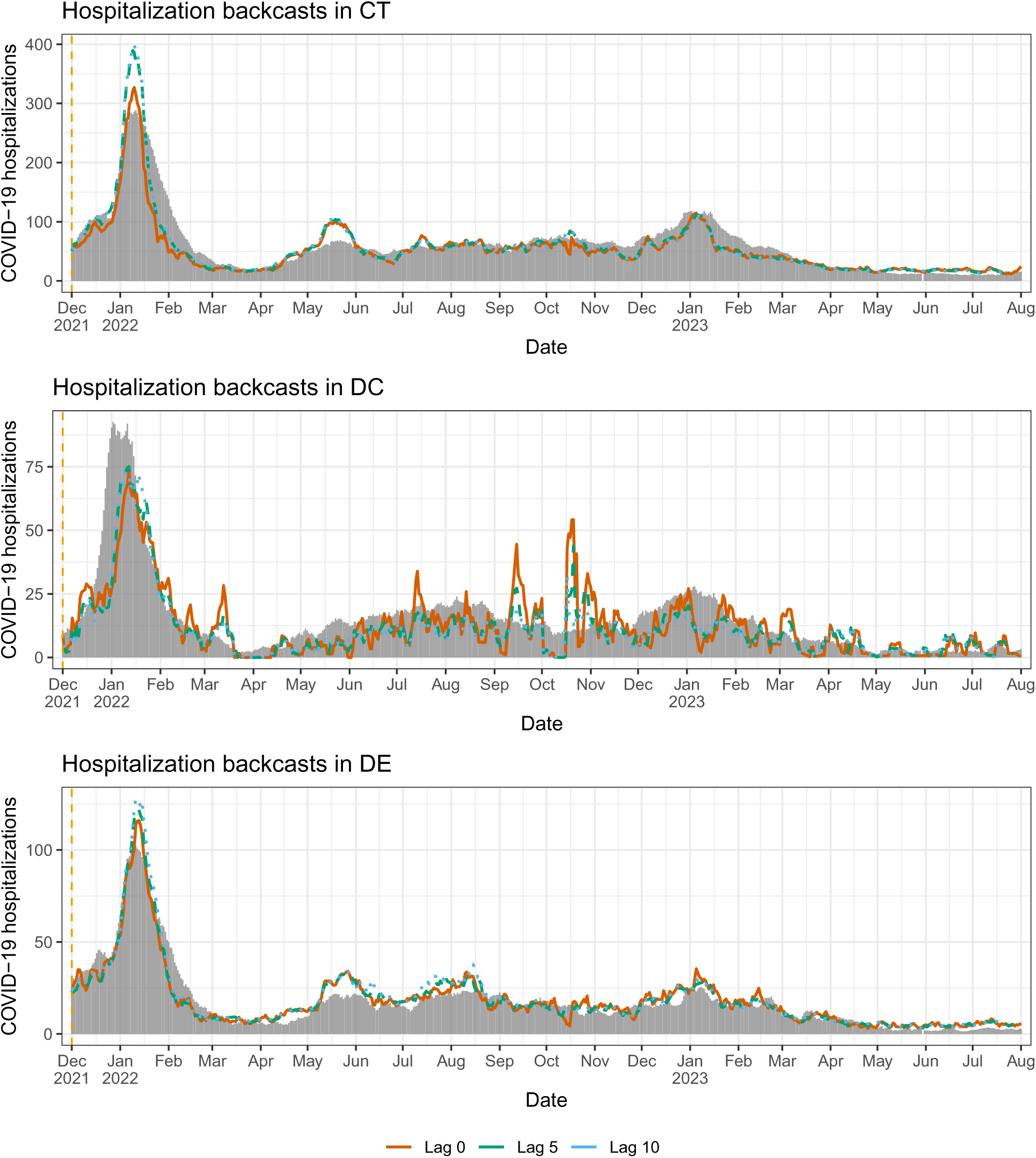
Backcasts from the mixed model in scenario 2, for CT, DC, DE.

**Figure 34:**
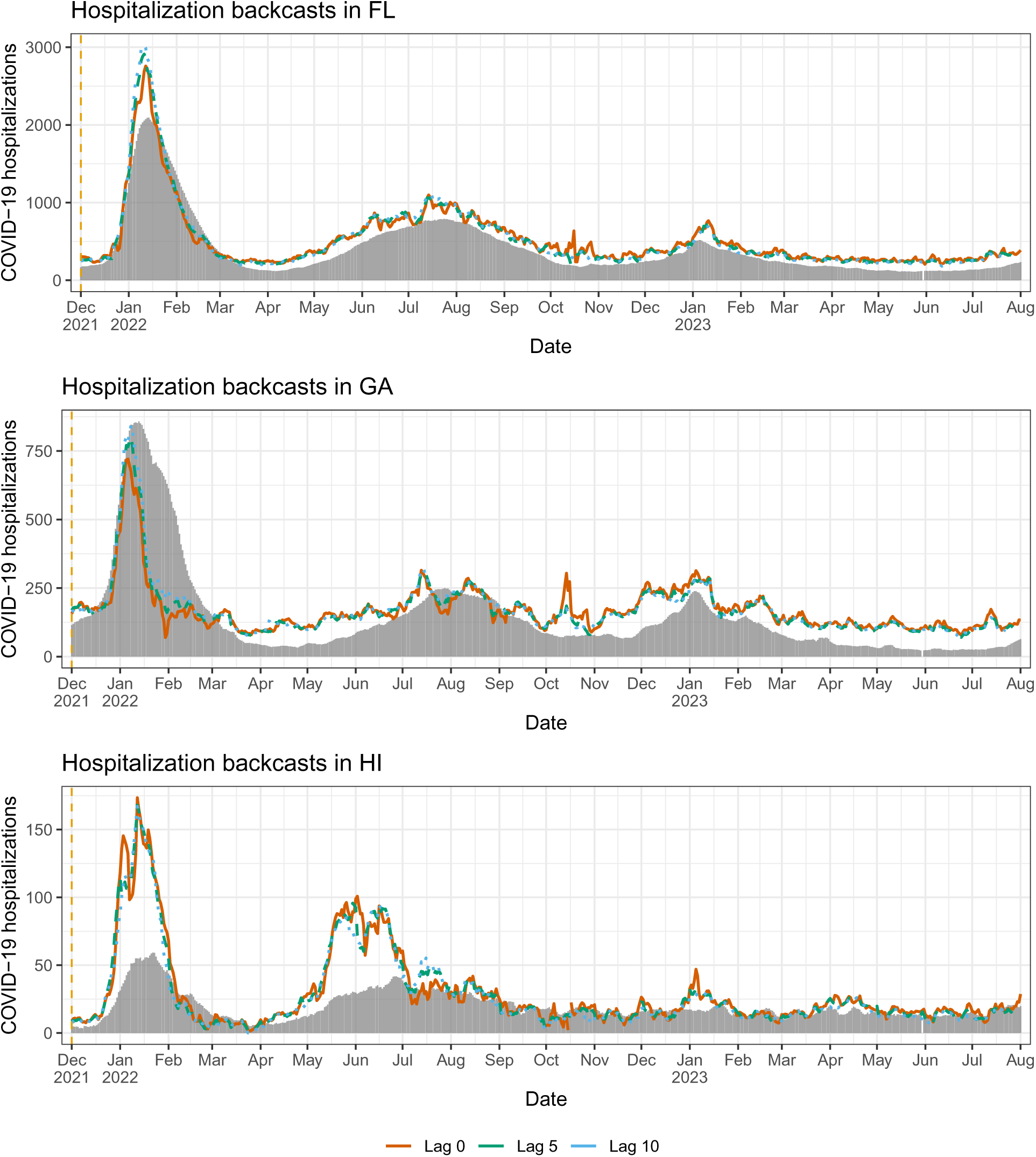
Backcasts from the mixed model in scenario 2, for FL, GA, HI.

**Figure 35:**
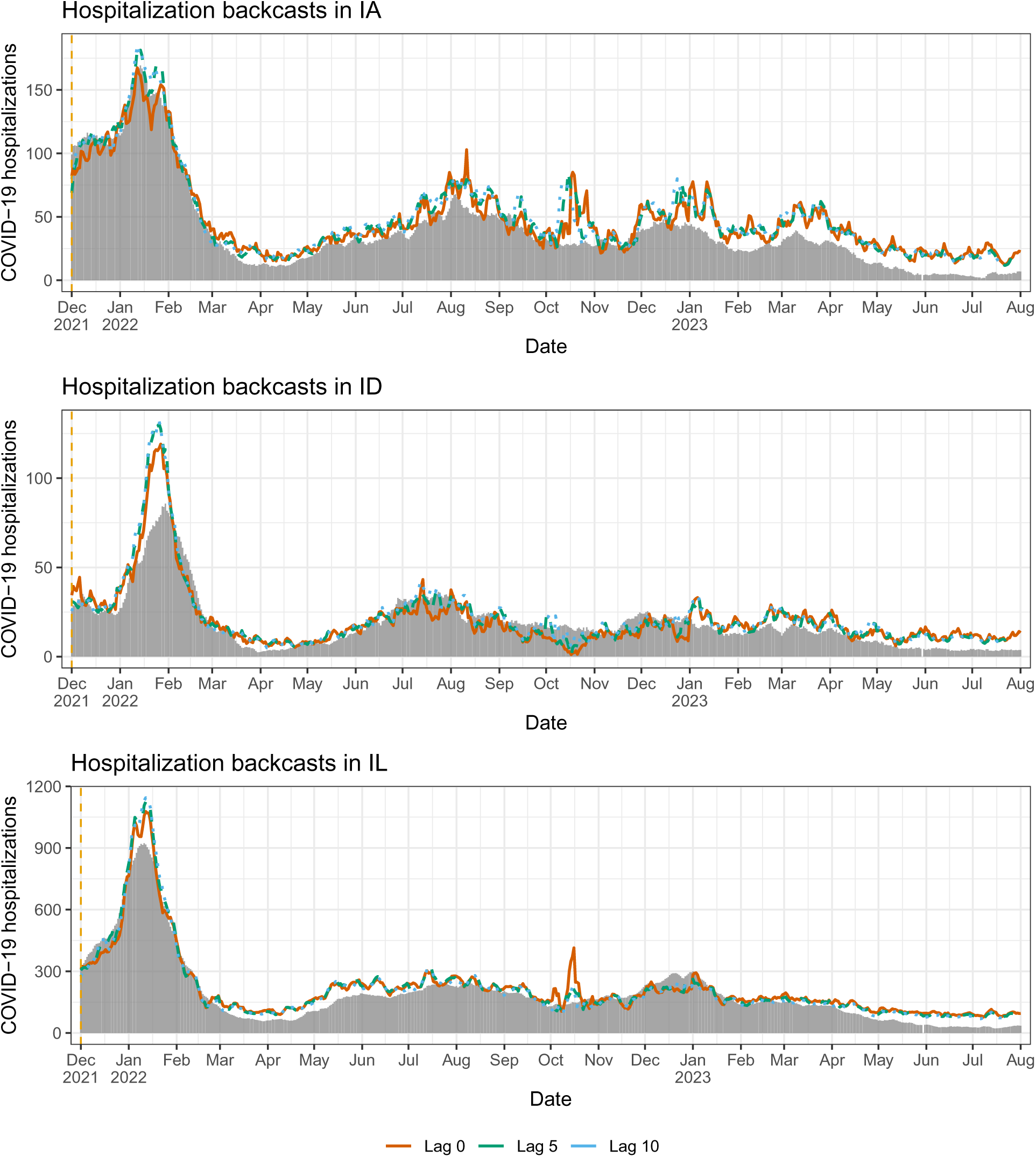
Backcasts from the mixed model in scenario 2, for IA, ID, IL.

**Figure 36:**
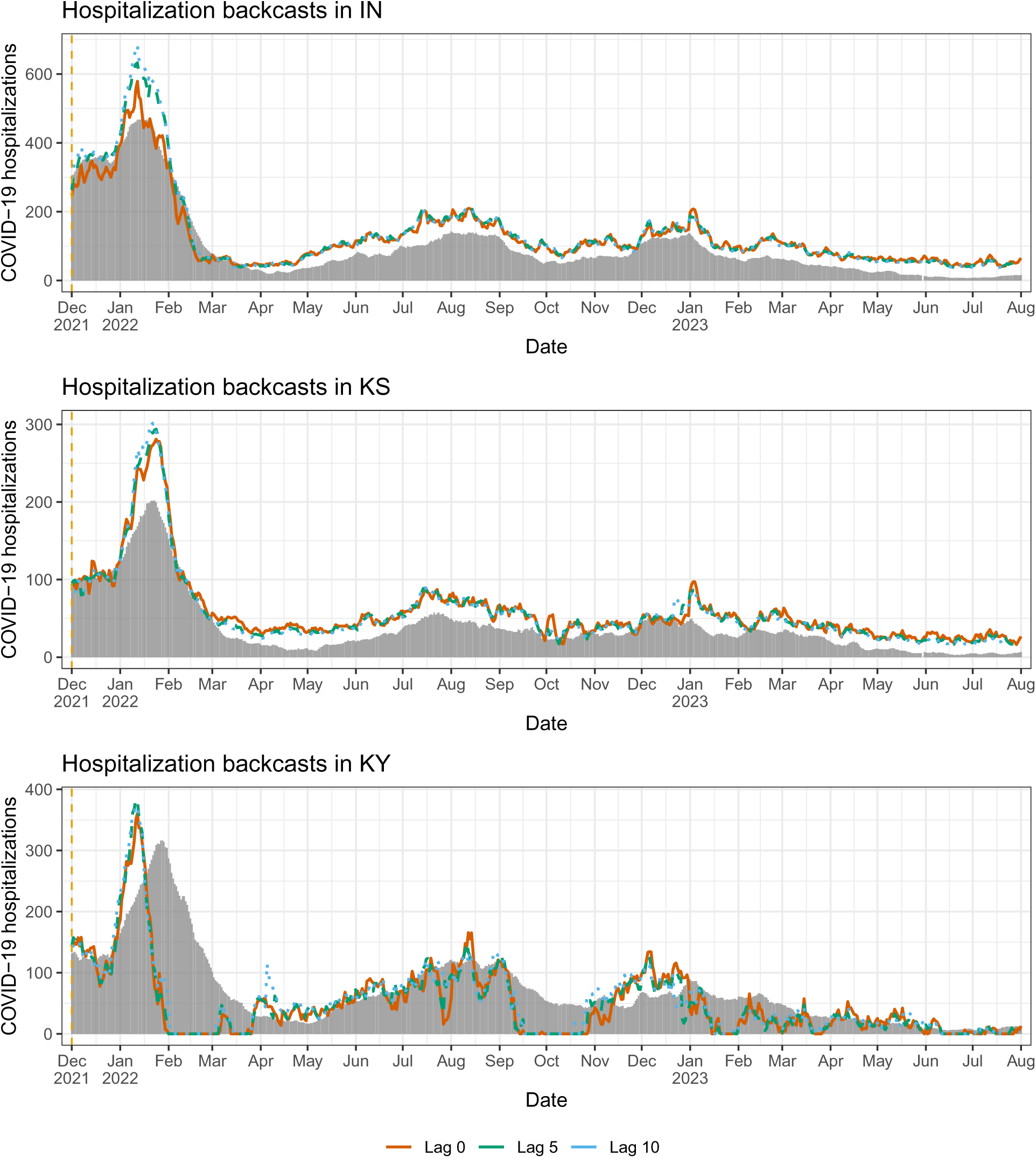
Backcasts from the mixed model in scenario 2, for IN, KS, KY.

**Figure 37:**
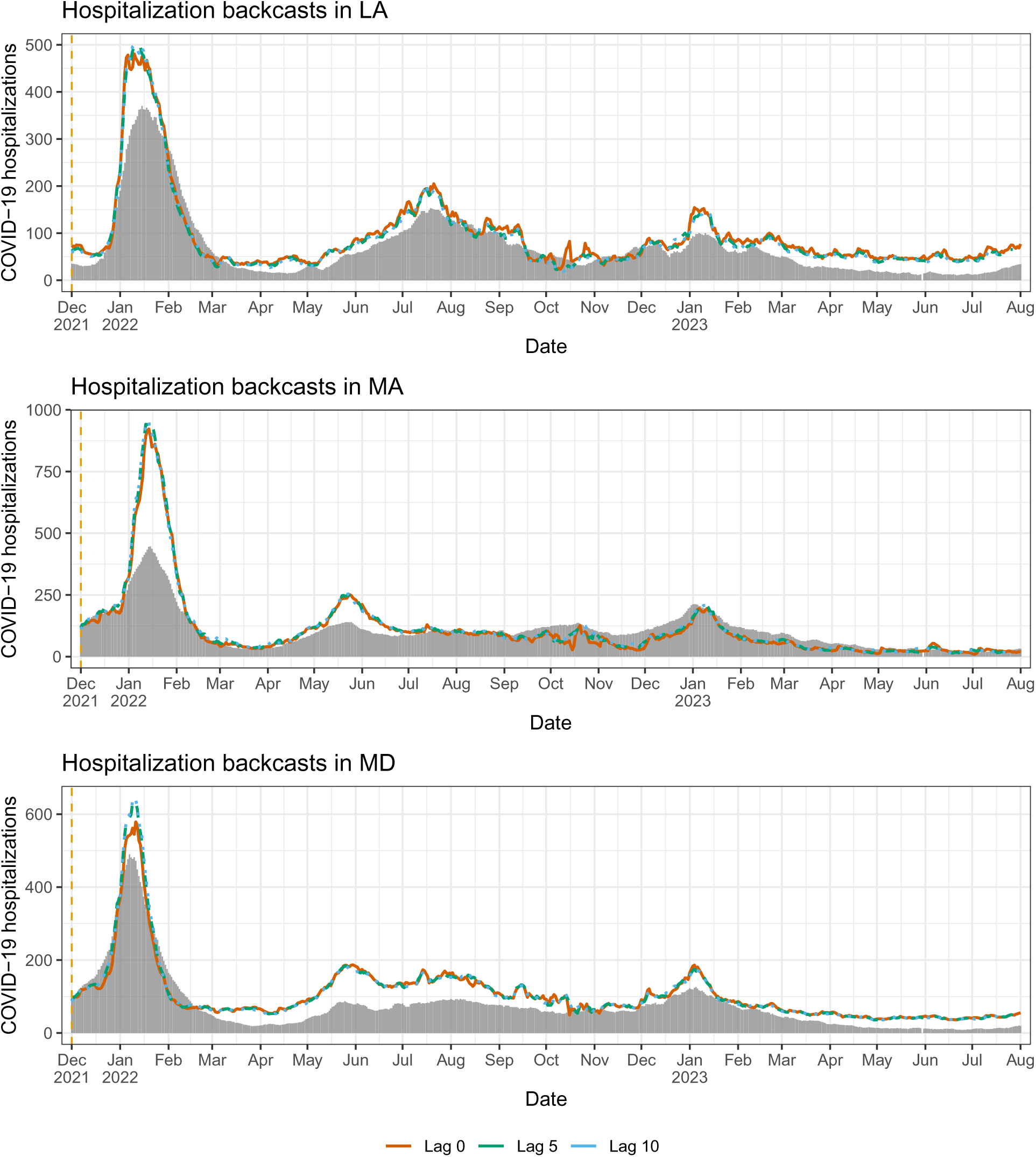
Backcasts from the mixed model in scenario 2, for LA, MA, MD.

**Figure 38:**
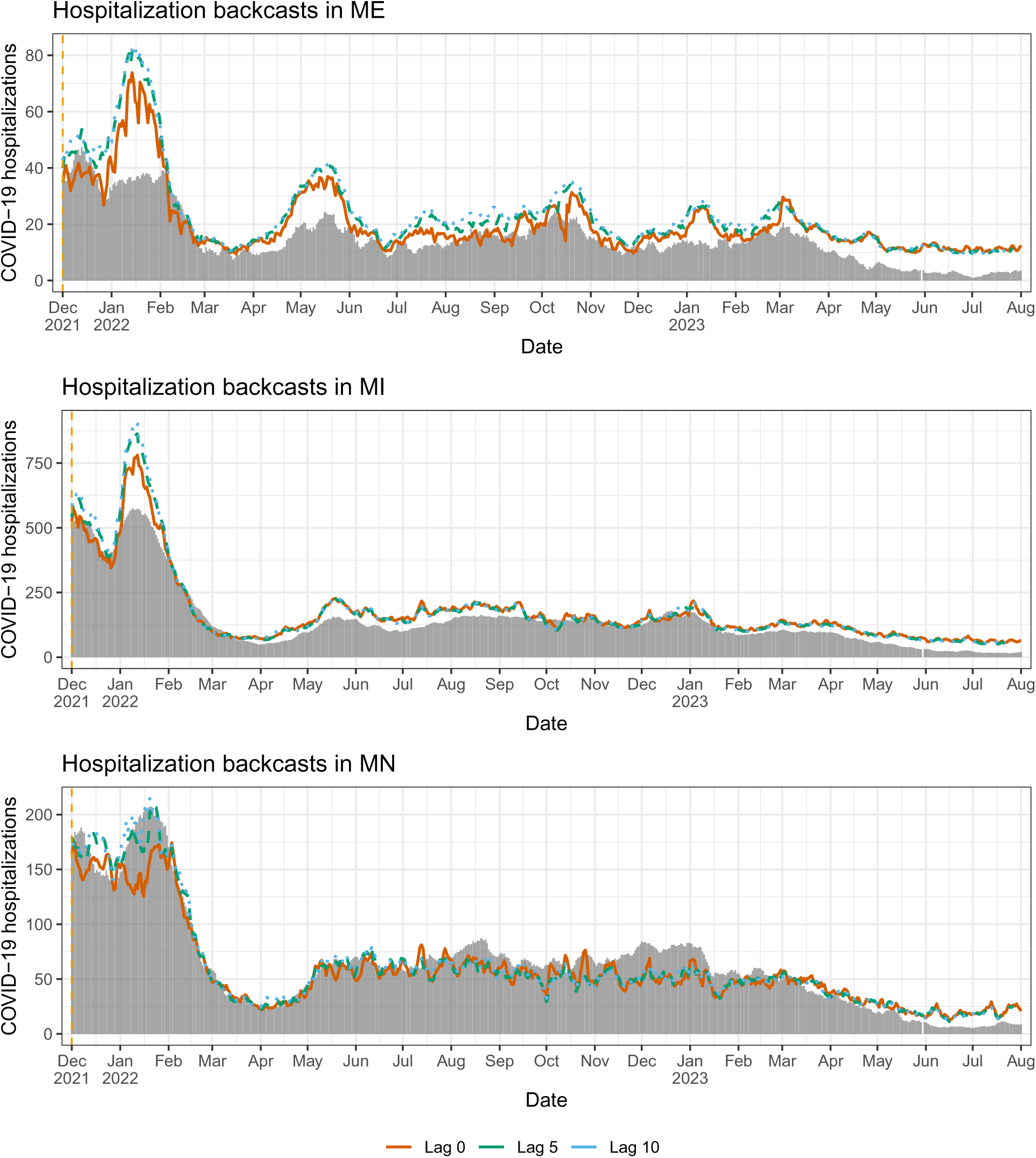
Backcasts from the mixed model in scenario 2, for ME, MI, MN.

**Figure 39:**
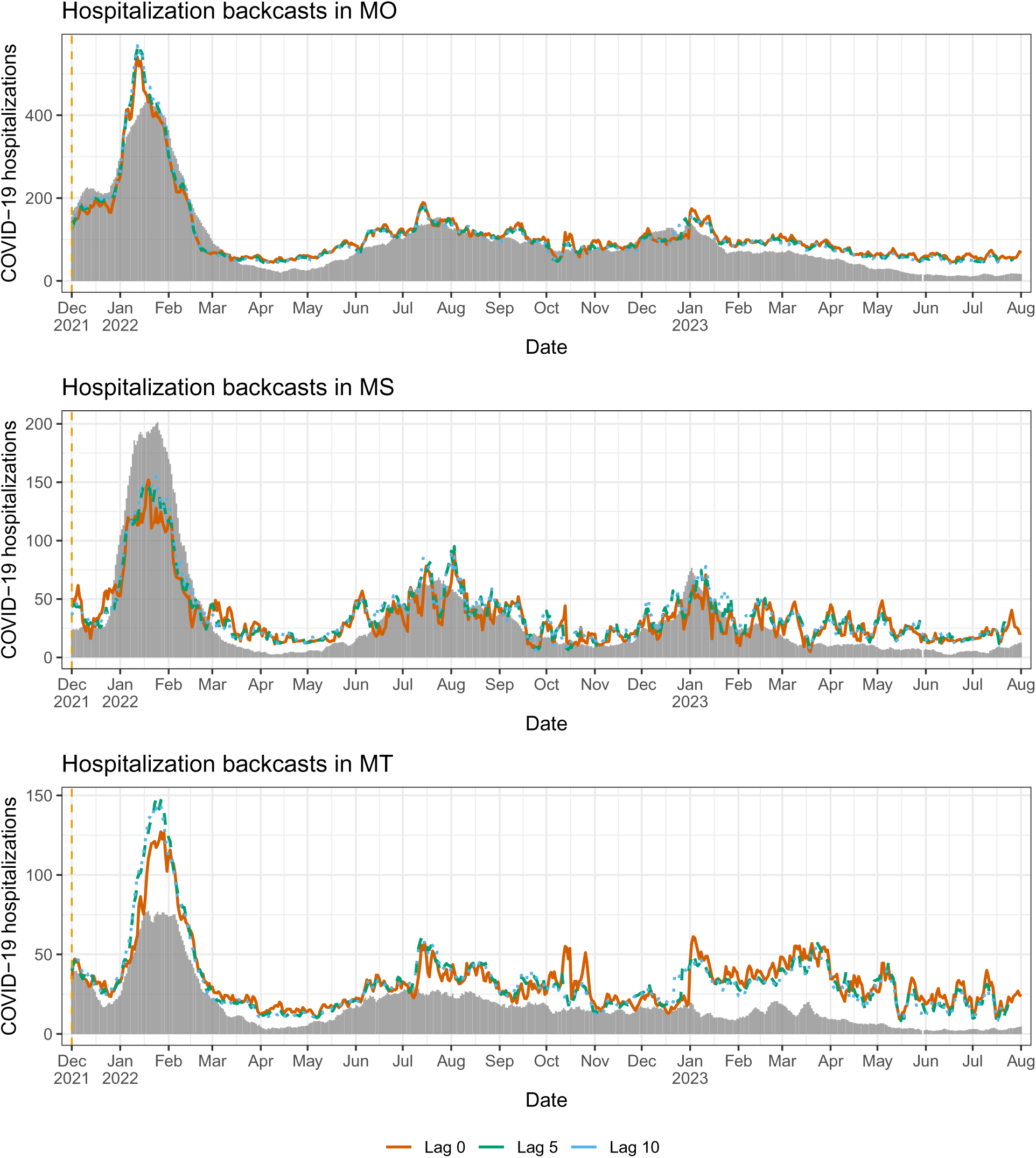
Backcasts from the mixed model in scenario 2, for MO, MS, MT.

**Figure 40:**
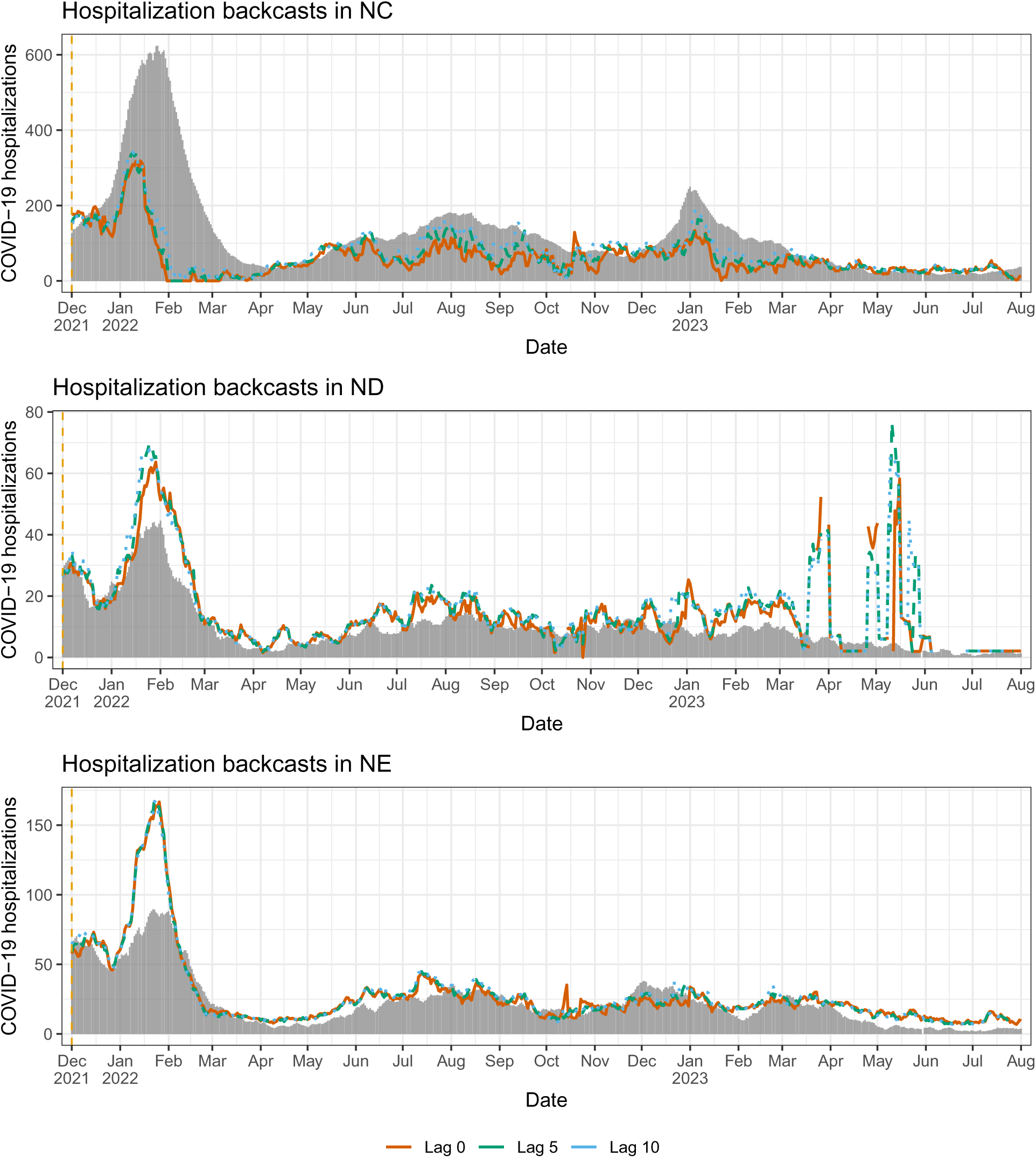
Backcasts from the mixed model in scenario 2, for NC, ND, NE.

**Figure 41:**
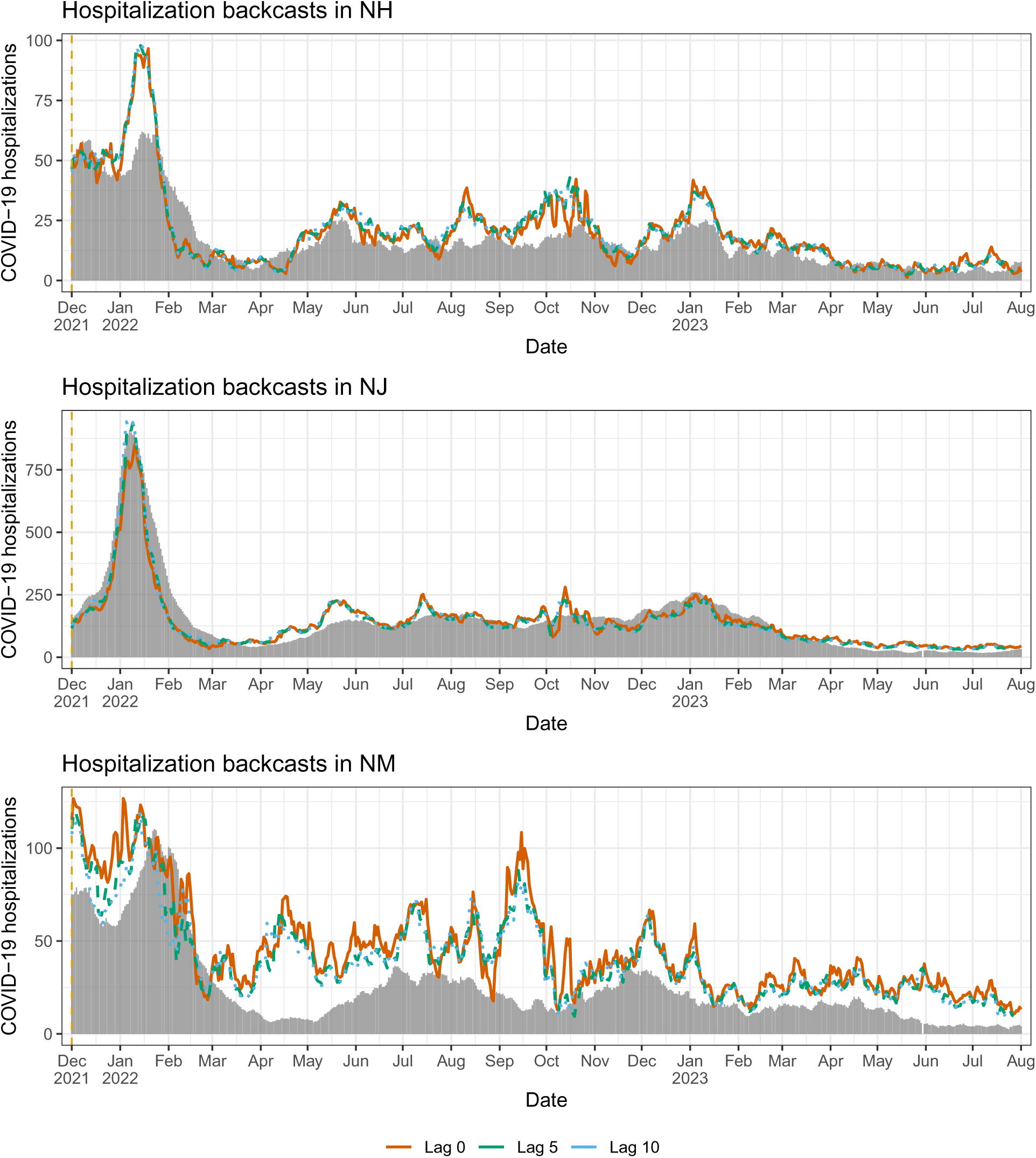
Backcasts from the mixed model in scenario 2, for NH, NJ, NM.

**Figure 42:**
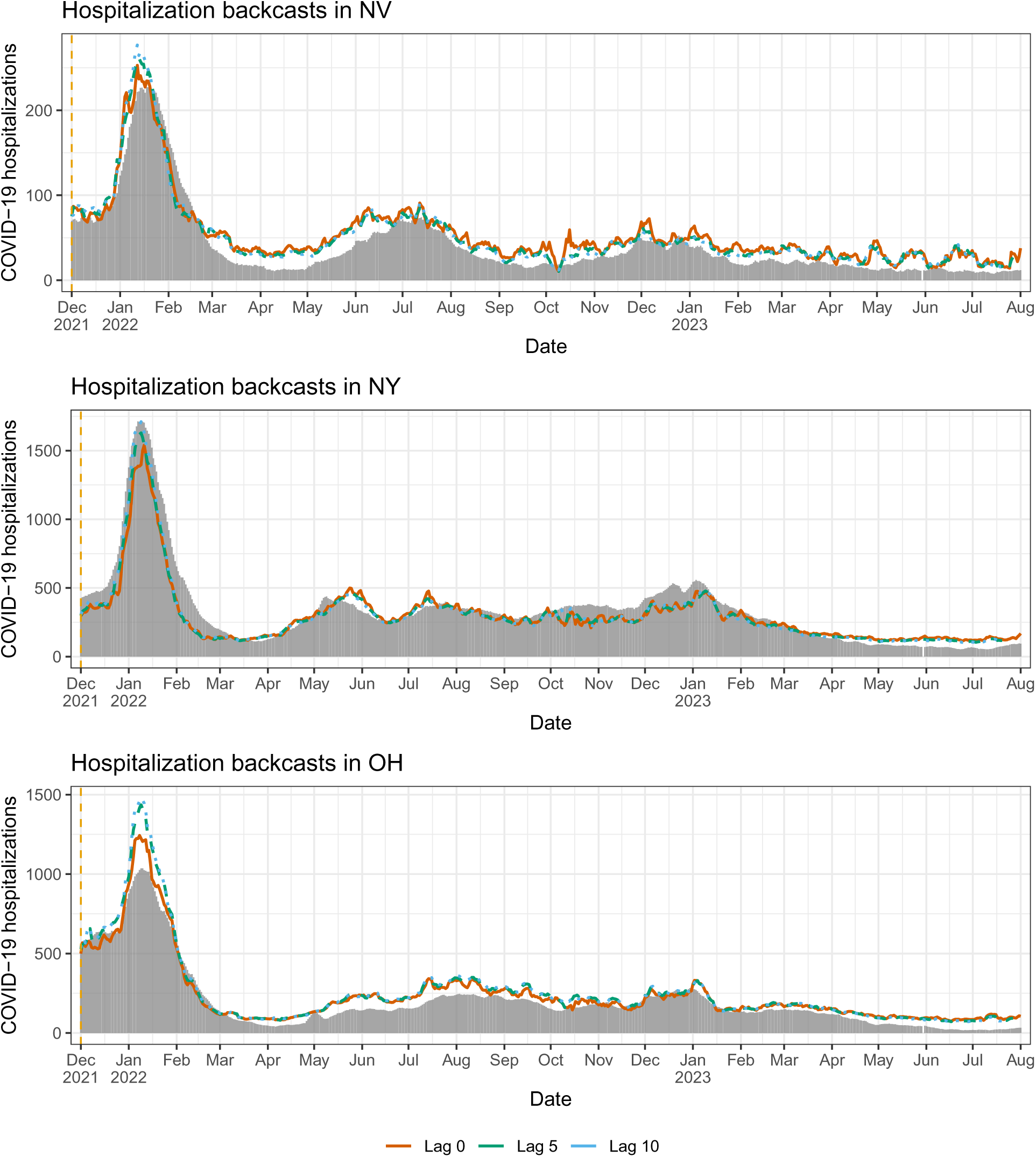
Backcasts from the mixed model in scenario 2, for NV, NY, OH.

**Figure 43:**
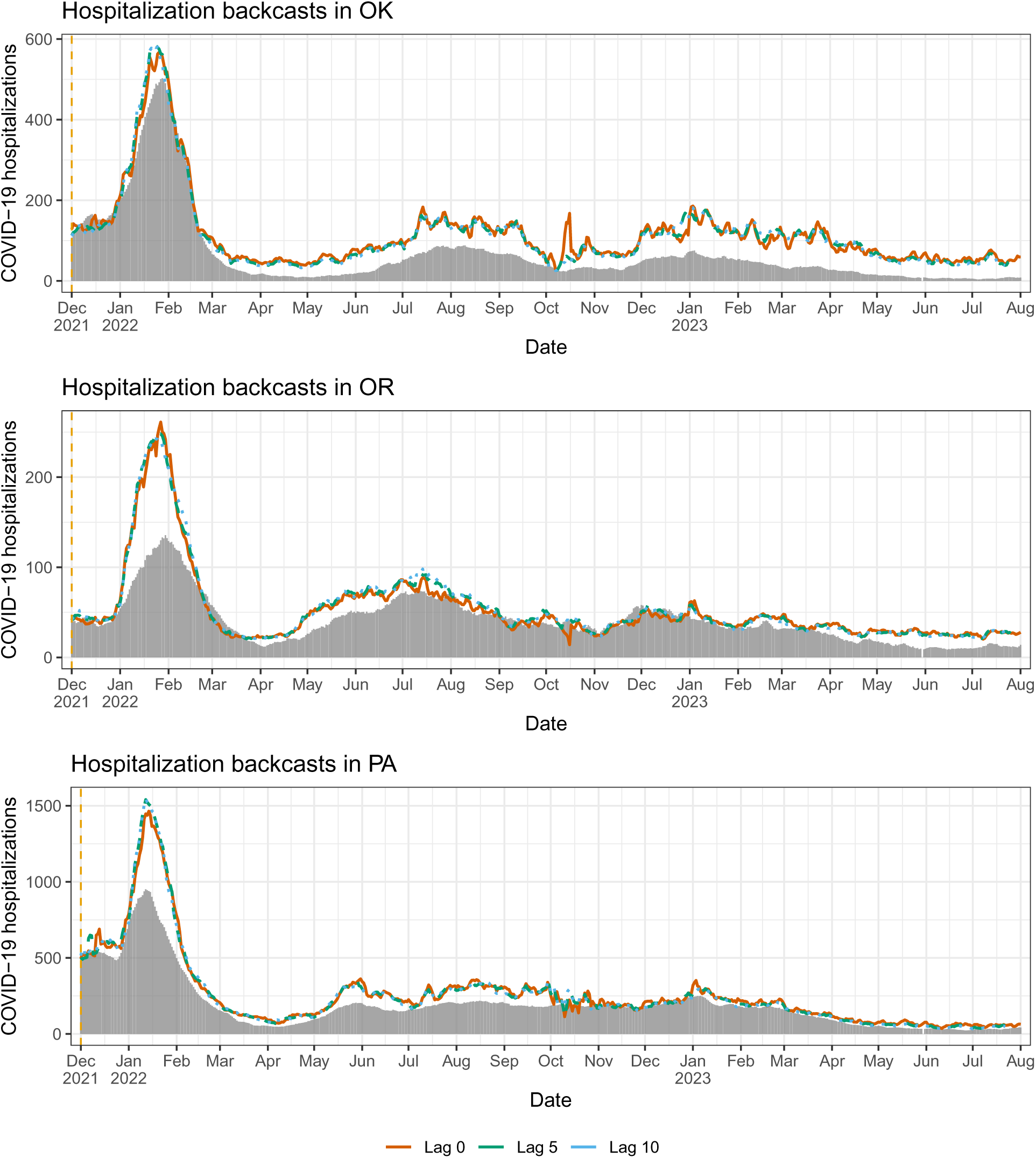
Backcasts from the mixed model in scenario 2, for OK, OR, PA.

**Figure 44:**
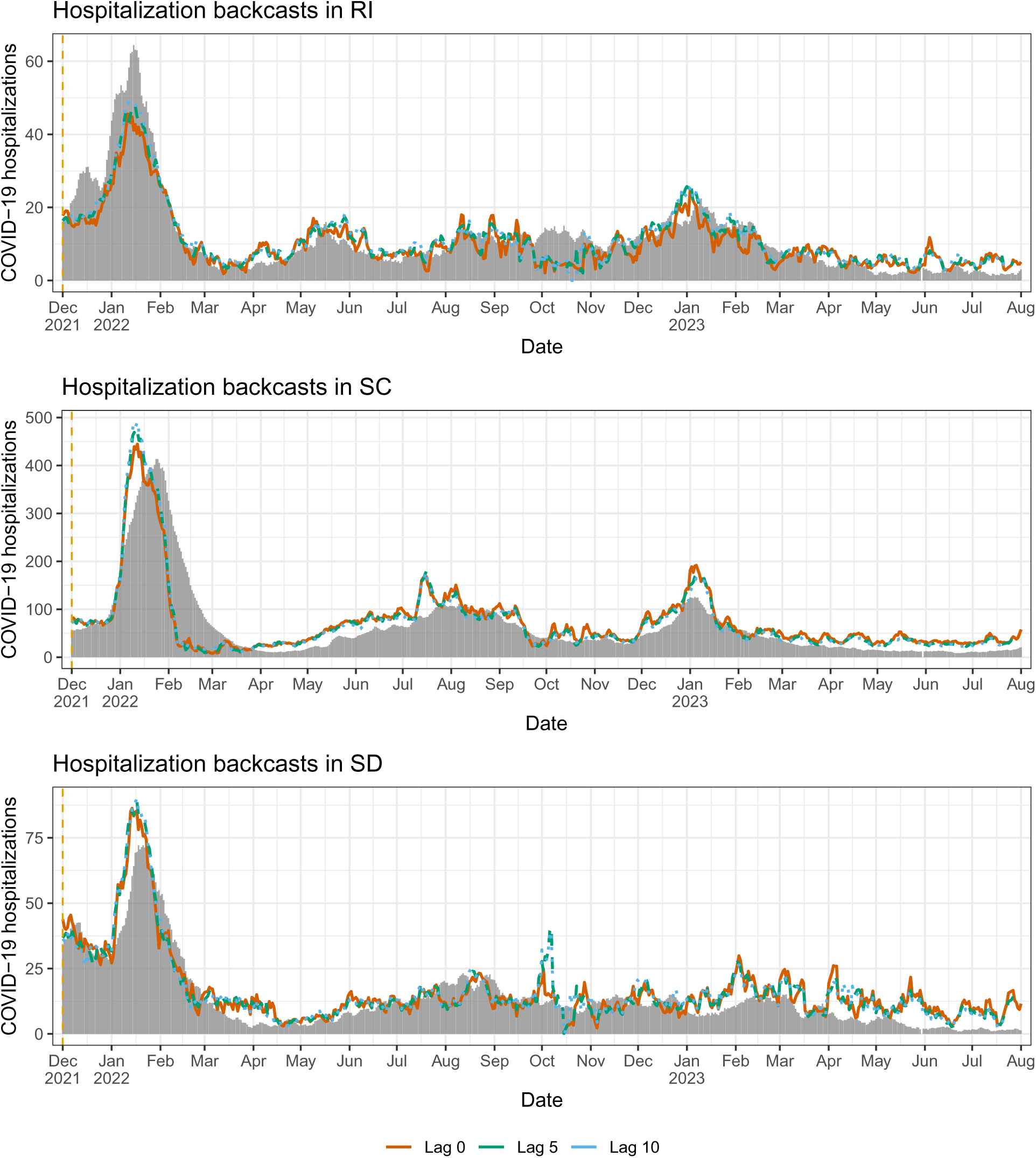
Backcasts from the mixed model in scenario 2, for RI, SC, SD.

**Figure 45:**
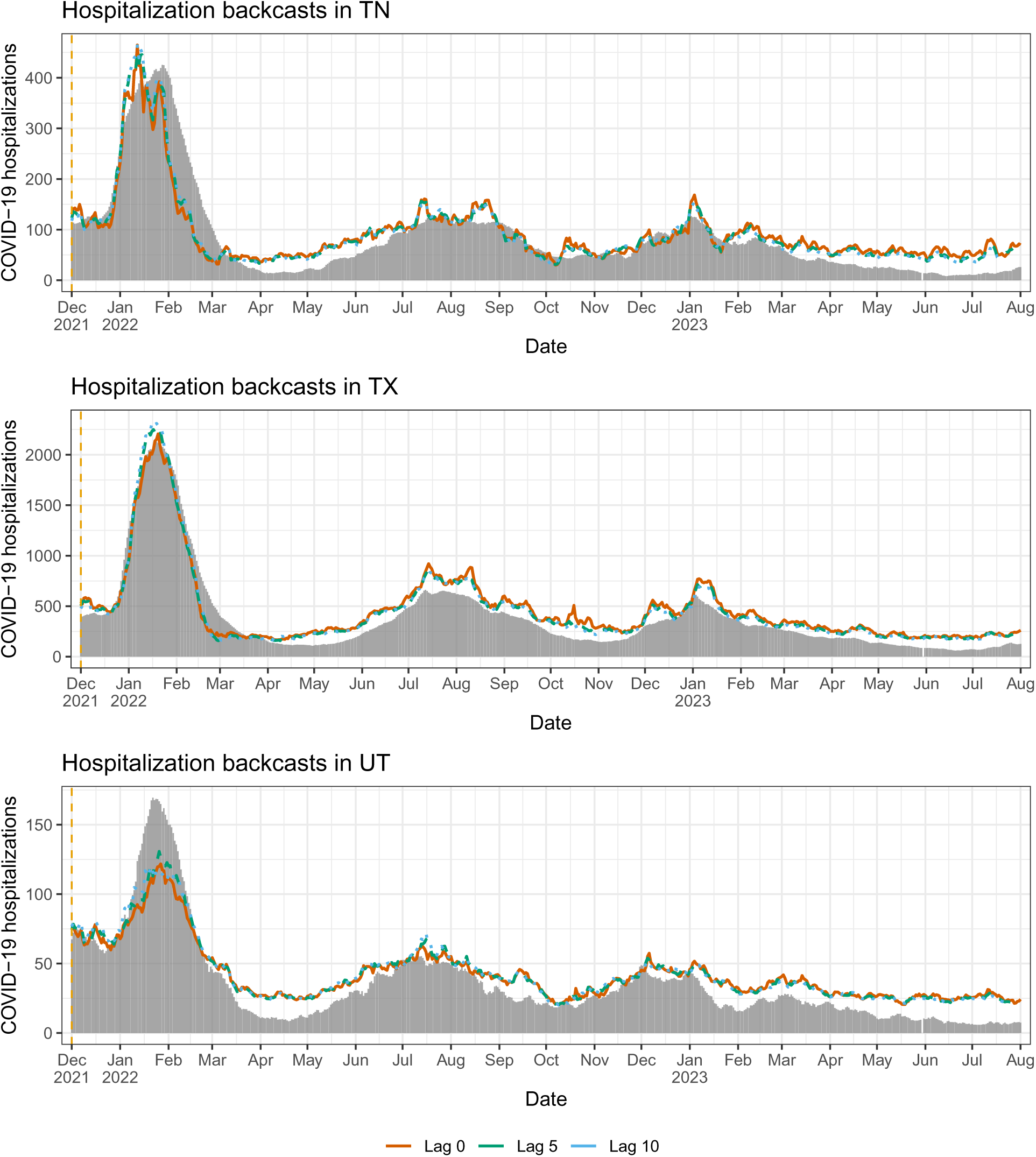
Backcasts from the mixed model in scenario 2, for TN, TX, UT.

**Figure 46:**
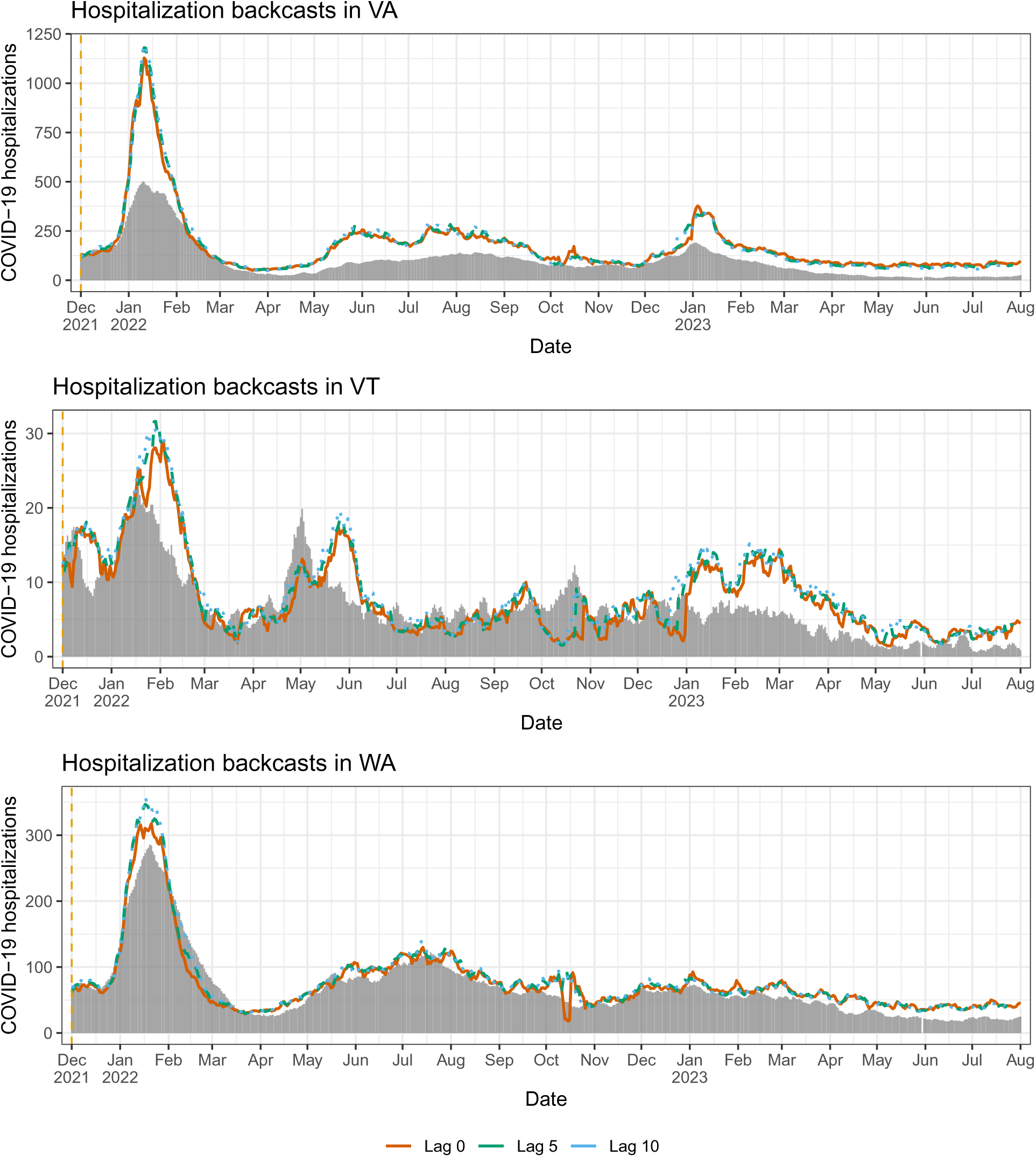
Backcasts from the mixed model in scenario 2, for VA, VT, WA.

**Figure 47:**
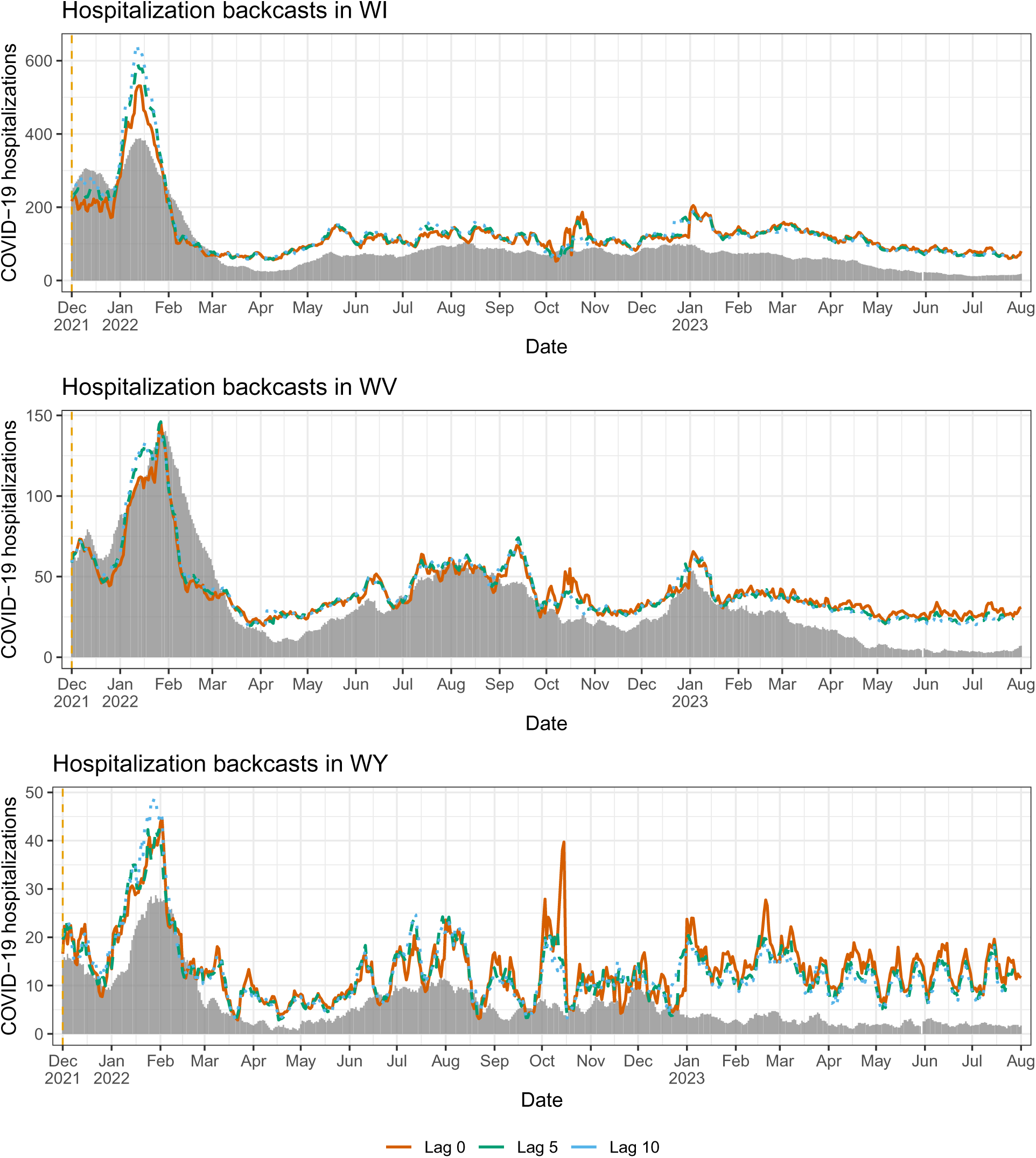
Backcasts from the mixed model in scenario 2, for WI, WV, WY.

